# Use of Baricitinib in Treatment of COVID-19: A Systematic Review

**DOI:** 10.1101/2021.12.26.21268434

**Authors:** A Sampath, A Banerjee, S Atal, R Jhaj

**Affiliations:** All India Institute of Medical Sciences, Bhopal (Madhya Pradesh), India

**Keywords:** Baricitinib, JAK kinase inhibitor, SARS-Cov-2, COVID-19, Clinical outcome, Mortality

## Abstract

**Objectives:** To assess the role of baricitinib alone or in combination with other therapies as a treatment for patients with COVID-19.

**Methods:** Systematic literature search was conducted in the WHO COVID-19 Coronavirus disease database to find clinical studies on use of baricitinib for treatment of COVID-19 between December 1^st^ 2019 and September 30^th^ 2021. Two independent set of reviewers identified the eligible studies fulfilling the inclusion criteria, and relevant data was extracted and a qualitative synthesis of evidence performed. The risk of bias was evaluated with validated tools.

**Results:** A total of 267 articles were found to be eligible after primary screening of title and abstracts. Following assessment of full texts, 19 studies were finally included for this systematic review, out of which 16 are observational, and 3 are interventional studies. Collating the results from these observational and interventional studies, baricitinib used as add on to standard therapy, either alone or in combination with other drugs, was found to have favourable outcomes in moderate to severe hospitalised patients with COVID-19. Furthermore, ongoing trials indicate that drug is being extensively studied across the world for its safety and efficacy in COVID-19.

**Conclusion:** Baricitinib significantly improves clinical outcomes in hospitalized patients with COVID-19 pneumonia and further evidence may establish the drug as a standard treatment among such patients.

## Introduction

In December 2019, a cluster of cases of pneumonia were identified in Wuhan, China which resembled viral pneumonia based on clinical characteristics and chest imaging (1). Analysis revealed a novel corona virus, which was named Severe Acute Respiratory Syndrome Coronavirus- 2 (SARS-CoV-2) which has since then spread globally to become one of the worst pandemics ever seen by humankind (2). At the time of writing, there have been more than 263 million confirmed cases of COVID-19 globally with 52,32,562 reported deaths (3). With multiple ‘waves’ of the Corona Virus Disease – COVID-2019 engulfing the world, causing widespread disruption of normal life and health along with financial strife, the medical and scientific community has focused all its attention on finding successful treatment options for managing this dreaded disease. Therapy approaches, particularly for hospitalised patients, are mainly focused at antiviral and anti-inflammatory activities of new or repurposed drugs.

Treatment guidelines from the WHO, various international authorities like the National Institutes of Health (NIH), U.S., National Institute for Health and Care Excellence (NICE), U.K. and regional authorities focus on the management of complications and providing supportive care with symptomatic treatment as per evidence based recommendations. A multitude of clinical trials and observational studies have been carried out worldwide to improve the survival rates in severe to critical COVID-19 patients, and reduce morbidity in terms of hospitalisation, time to recovery, and symptomatic or virological response in lesser severity of the illness. At present, the established therapeutic options for COVID-19 still remain sparse and include antiviral therapies like remdseivir, immunomodulators like corticosteroids, IL-6 inhibitors and, more recently, specific monoclonal antibodies like REGN-COV2 (casirivimab and imdevimab combination) amongst others (4).

Baricitinib, a novel Janus kinase (JAK) inhibitor, in combination with remdesivir first received emergency use authorization (EUA) by the U.S. Food and Drugs Administration (FDA) in November, 2020 (5), which has subsequently been revised to allow its use as a standalone therapy. At present, European Medicines Agency (EMA) is evaluating applications to include baricitinib as a part of COVID-19 treatment protocol for patients above the age of 10 years of age who require supplemental oxygen therapy (6). In May 2021, the drug regulatory authority of India - Central Drugs Standard Control Organization (CDSCO) has also approved baricitinib in combination with remdesivir, for treatment of confirmed COVID-19 hospitalized adults requiring supplemental oxygen (7). Baricitinib is an oral inhibitor of JAK 1 and 2, which was originally approved for rheumatoid arthritis in 2017 in U.S. and Japan (8). JAKs are a family of enzymes involved in the inflammatory pathway that modulate the signaling pathway, preventing activation of signal transducers and activators of transcription (STATs) for the cytokines like IL-2, 6, 10, 12, IFN- etc. JAK inhibitors (JAK i’s) can prevent the dysregulated production of proinflammatory cytokines primarily involved in cellular survival, proliferation, differentiation, and immigration; proving to be clinically useful in immune, inflammatory, and hematopoietic diseases (9, 10). Baricitinib has thus been evaluated in treatment of COVID-19 with the rationale of suppressing the hyperinflammatory state of cytokine release syndrome (CRS) or hypercytokinemia (cytokine storm) associated with COVID-19 (11).

While previously published systematic reviews (including living reviews) on COVID-19 therapies (12, 13) or on JAK inhibitors (14, 15) have provided updates on the use of baricitinib or other JAK inhibitors, there has been no dedicated systematic review published till date focusing solely on baricitinib. This systematic review, conducted as per the Cochrane methodology, attempts to provide a comprehensive qualitative synthesis of the evidence available regarding use of baricitinib in treatment of COVID-19.

**Figure 1:**
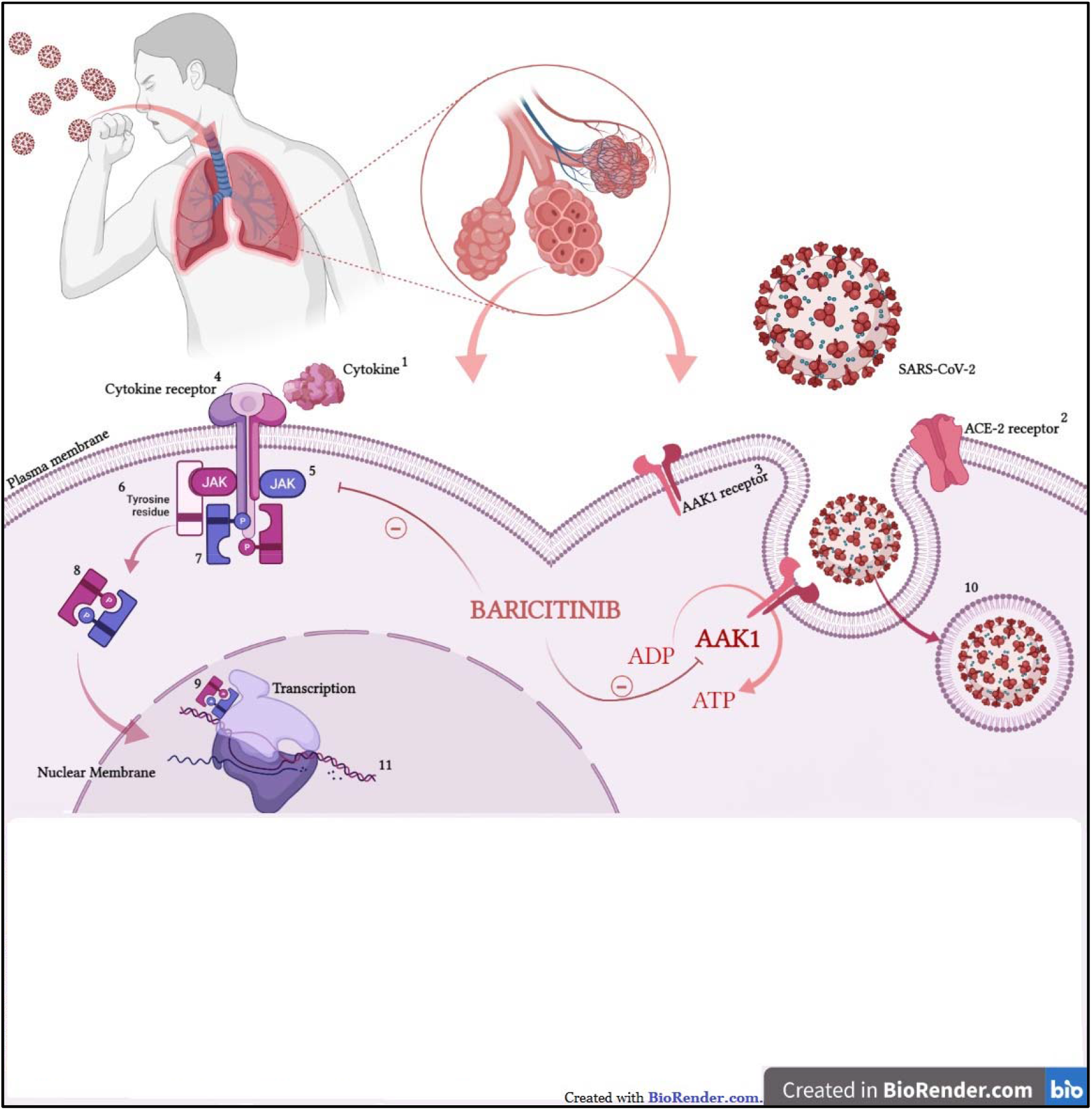
Schematic representation of the mechanism of action of Baricitinib. Mainly functions on three different aspects of Corona Virus Disease, inhibits Viral entry, Inhibits Inflammation by blocking JAK-STAT signal transduction pathways and alters the Immune Status via JAK-STAT inhibition. [1]Interleukins (IL-2, IL-4,IL- 7,IL-g,IL-15,IL-21,IL-6) mainly. IL-2, IL-4.IL- 7,IL-g,IL-15,IL-21 via JAK1/3 STAT1/3/5 to regulate hemostasis, lymphocyte I proliferation where as IL6 acts via Tvrosine kinase JAKl/2 STAT3 to regulate T cell differentiation and inflammation, all of which is inhibited bv Baricitinib. [2]ACE-2 receptor-Angiotensin Converting enzyme-2 acts as a docking receptor for the attachment of SARS C0V-2. [3] AAKl-Adaptor-associated protein I [4]kinase i(alias AP2-associated protein kinase 1, transmembrane enzyme involved in clathrin mediated endocytosis.Baricitinib blocks the enzyme activation hence prevents SARS-CoV 2 Endocytosis.[4] Cytokine, receptors type I and II. [5] Janus Activated Kinase are intracellular non-receptor tyrosine kinases that mediate cytokine signal transduction via the JAK-STAT pathway.Baricitinib acts as a non-selective JAK inhibitor.[6] Tyrosine residues which aid binding of SH2 domain proteins such as STAT. [7] STAT - Signal Transduction Activated Transcription, is a downstream transducer of the JAK enzyme that exists as a inactivated hypophosporylated state in association with JAK intracellularly.f 8] Phosphorylated active form of STAT. [9] Transcriptional Activation of specific genes. [10] Transcription following by translation of proteins leading to the regulated effects specified in [1].

### Methodology

The primary objective of this systematic review is to assess the efficacy and safety of baricitinib therapy in COVID-19, either alone or in combination with other therapies, on the basis of the evidence available through the data published or pre-published from completed clinical studies.

### Eligibility Criteria

The following criteria were used for selecting the studies for the review -

*For inclusion:*

- Interventional studies (RCT’s and non RCT’s) assessing the efficacy and/or safety of baricitinib in COVID-19.
- Observational studies assessing the efficacy and/or safety of baricitinib in COVID-19.
- Published or pre-published studies in the English language from any country satisfying the above criteria.

*For exclusion:*

- Any study published before 2019.
- Studies not conducted in human subjects in a clinical setting (in-vitro/ animal / AI or any other non-clinical studies).
- Case reports, review articles, publications with only abstracts available, or available as conference proceedings, editorial/comments, author response or book chapter, incomplete registered trials.
- Studies with duplicate, or overlapping data or data that cannot be reliably extracted.

### Search Strategy

Information was extracted from the WHO COVID-19 Database (WHO COVID-19 Global literature on Coronavirus Disease) which is a comprehensive database of COVID-19 research articles (16). This database contains global literature from all major standard electronic databases of published, pre published and grey literature sources such as MEDLINE, Scopus, ProQuest Central, ELSEVIER, Web of Science, EMBASE, ScienceDirect, Academic Search Complete, MDPI, PubMed, Wiley, LILACS (Americas), PMC, SSRN, Taylor & Francis, Sage, CINAHL, CAB Abstracts, GIM, APA PsycInfo, Indonesian Research, Centers for Disease Control and Prevention, Africa Wide Information, J-STAGE, ChemRxiv, BioMed Central, Oxford Academic, National Academies Press, Sage, China CDC Weekly, Mary Ann Liebert, WPRIM (Western Pacific), Springer, PAHOIRIS, WHO COVID, MAL, JAMA Network, Relief Web, ICTRP, medRxiv, BMC, bioRxiv, Science Direct.

Additionally search was also conducted separately on PubMed, to cross check and account for any articles that may not have been added to the WHO database. Furthermore, a manual search was performed using the bibliography of selected articles, and any studies that satisfied eligibility criteria were added. We searched all information sources from 1 December 2019 to 30 September 2021.

### Data Collection

Reviewers applied the search criteria in WHO COVID-19 database, using advanced search Boolean Operations. A computer-based literature search was conducted on the databases; on the COVID WHO unified database using the keywords in Titles, Abstracts and Authors: (*Baricitinib* **OR** *Janus Kinase* **OR** *JAK-STAT*) **AND** (*Therapy* **OR** *Treatment*) and on PubMed with the advance search keywords: (*Baricitinib* **OR** *Janus Kinase* **OR** *JAK-STAT*) **AND** (*Therapy* **OR** *Treatment*) **AND** (*2019 novel Coronavirus disease* **OR** *COVID-19* **OR** *SARS-CoV-2* **OR** *novel Coronavirus infection* **OR** *2019-ncov infection* **OR** *Coronavirus disease 2019* **OR** *Coronavirus disease-19* **OR** *2019-ncov disease* **OR** *COV* **OR** *Coronavirus* **OR** *COVID* **OR** *COVID19* **OR** *COVID-19* **OR** *SARS CoV-2*). A variety of COVID synonyms were used only for PubMed as the WHO COVID database is a consolidation of only COVID related results.

Out of 3,68,617 records, a total of 267 results were found eligible and exported onto Microsoft Excel sheet. All exported records were searched for to remove duplicates using the “Author-Title-Year” criteria. All references that had the same title and author, and were published in the same year or the same journal were removed. Essential information (including abstracts and weblinks) for all the remaining articles was exported for screening.

**Figure 2:**
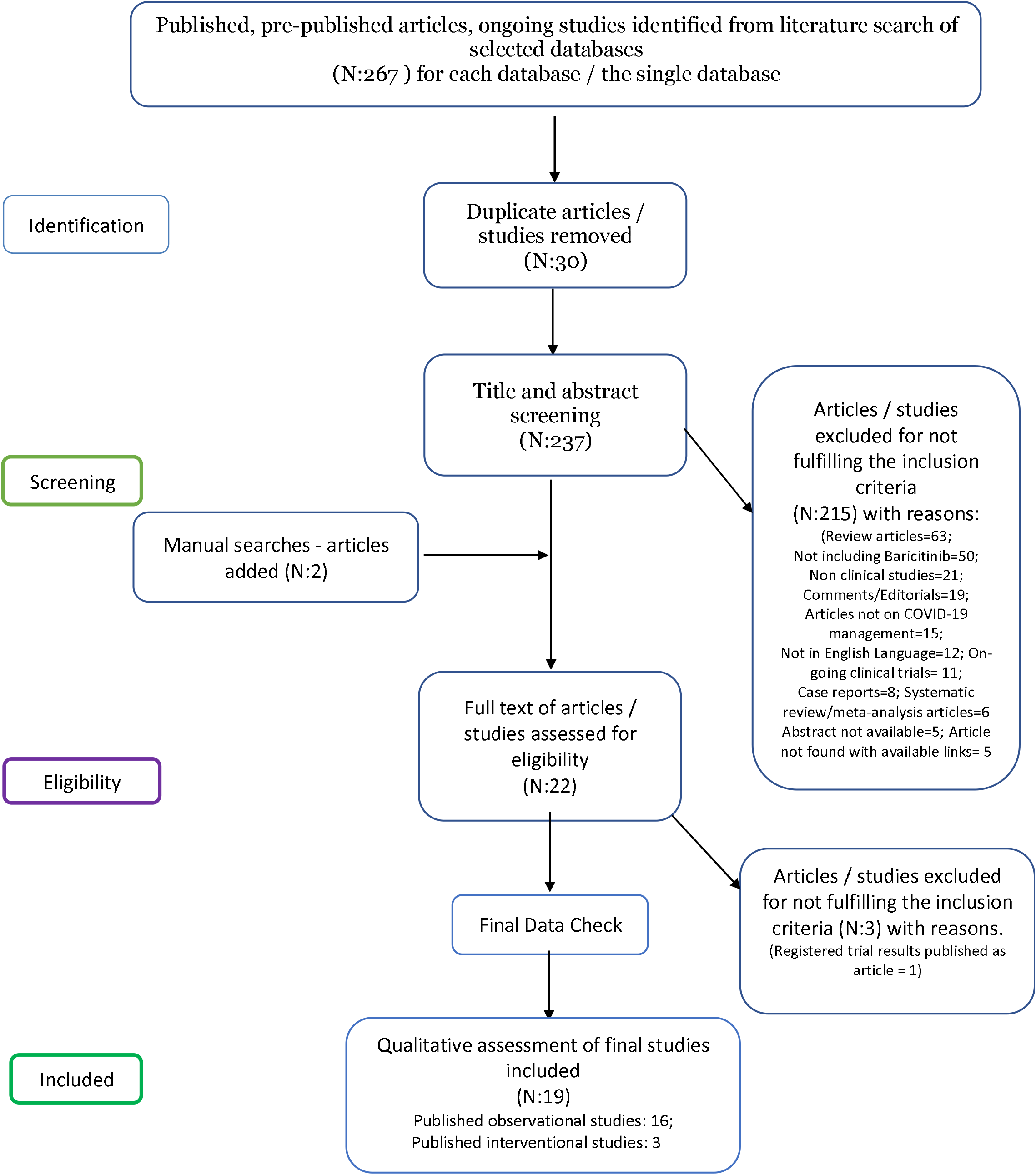
PRISMA flowchart for depicting the methodology

### Study Selection and data extraction

The titles and abstracts were independently screened by two sets of reviewers (AS and SA, AB and RJ) based on the eligibility criteria. Each article was checked by two independent reviewers. Any disagreements regarding study eligibility during screening were settled through consensus decision. All excluded records were given appropriate reasons. Screened articles were then assessed for availability of full texts, which were finally downloaded) and reviewed by two independent reviewers. The decision to include or exclude articles was agreed upon by the two sets of reviewers to arrive at the final set of eligible studies to be included for data extraction for the systematic review.

Data was then extracted from eligible studies related to these pre-decided data items - study characteristics (publication status, study type, design), patient characteristics (country, age, sex, comorbidities, setting and type of care, and severity of COVID-19 symptoms), interventions studied, and outcomes reported (clinical outcomes pertaining to COVID-19 related morbidity or mortality, virologic outcomes, adverse events), apart from the title, authors, source, database, year of publication, country of study. Outcomes of importance were decided upon consensus - based on their relevance by the review team. Standard definitions were used for the severity of disease and various outcome parameters (17). The complete extracted data was then thoroughly checked finally by two reviewers independently. If any disagreement was noticed, the corresponding author took the final decision.

### Risk of bias

The Cochrane tool for assessing the risk of bias in randomised trials (RoB 2.0) (18) was used for the reviewers to segregate the published RCT’s as either at low risk of bias, some concerns—probably low risk of bias, more concerns—probably high risk of bias, or high risk of bias. For the observational studies (Non-Randomised Studies of Intervention, NRSIs), i.e cohort, case control and cross sectionals studies, the “Study Quality Assessment tool” (19) was used for critical appraisal of the internal validity of the studies and bias.

## Results

After thorough assessment, 20 studies were finally included in this systematic review. Out of these, 17 are observational and 3 are interventional studies. Details of these published / pre published studies are summarised in table 1 along with the assessment of study quality / risk of bias.

**Table 1:** Summary of the observational and interventional studies

| *S. No.* | *Title of study* | *Authors* | *Study location* | *Study design* | *Bias assessment* |
| --- | --- | --- | --- | --- | --- |
| *Observational Studies* |  |  |  |  | *(Study Quality Assessment Tool)* |
| 1 | Impact of high dose of baricitinib in severe COVID-19 pneumonia: a prospective cohort study in Bangladesh. | Hasan MJ, Rabbani R, Anam AM, Huq SM, Polish MM, Nessa SS, et al. | Bangladesh | Prospective Cohort Study | *Good* |
| 2 | Use of Baricitinib in Patients With Moderate to Severe Coronavirus Disease 2019 | Titanji BK, Farley MM, Mehta A, Connor-Schuler R, Moanna A Cribbs SK, et al. | United States | Retrospective Cohort Study | *Fair* |
| 3 | Baricitinib restrains the immune dysregulation in patients with severe COVID-19. | Bronte V, Ugel S, Tinazzi E, Vella A, Sanctis FD, Canè S, et al. | Italy | Prospective Cohort Study | *Fair* |
| 4 | Role of Low-Molecular-Weight Heparin in Hospitalized Patients With Severe Acute Respiratory Syndrome Coronavirus 2 Pneumonia: A Prospective Observational Study | Falcone M, Tiseo G, Barbieri G, Russo A, Virdis A, Forfori F, et al. | Italy | Prospective Cohort Study | *Good* |
| 5 | Experience With the Use of Baricitinib and Tocilizumab Monotherapy or Combined, in Patients With Interstitial Pneumonia Secondary to Coronavirus COVID19: A Real-World Study. | Rosas J, Liaño FP, Cantó ML, Barea JM, Beser AR, Rabasa JT, et al. | Spain | Case Control Study | *Fair* |
| 6 | Additional baricitinib loading dose improves clinical outcome in COVID-19 | Hasan MJ, Rabbani R, Anam AM, Huq SM, Polish MM, Nessa SS, et al. | Bangladesh | Case Control Study | *Good* |
| 7 | Clinical Outcomes in a Cohort of Non-Ventilated COVID-19 Patients with Progressive Hypoxemia and Hyper-Inflammatory Response Treated with Baricitinib | Milligan PS, Amsden J, Norris SA, Baker RL, Myers J, Preston F, et al. | United States | Retrospective Cohort Study | *Fair* |
| 8 | Beneficial impact of Baricitinib in COVID-19 moderate pneumonia; multicentre study. | Cantini F, Niccoli L, Nannini C, Matarrese D, Natale M Lotti P, et al. | Italy | Retrospective Cohort Study | *Fair* |
| 9 | Baricitinib therapy in COVID-19: A pilot study on safety and clinical impact. | Cantini F, Niccoli L, Nannini C, Matarese D, Natale M Lotti P, et al. | Italy | Ambivalent Cohort Study | *Fair* |
| 10 | Biological agents for rheumatic diseases in the outbreak of COVID-19: friend or foe? | Santos CS, Fernández XC, Moriano CM, Álvarez ED, Castro CA, Robles AL, et al. | Spain | Retrospective Cohort Study | *Good* |
| 11 | Baricitinib plus dexamethasone compared to dexamethasone for the treatment of severe COVID-19 pneumonia: A retrospective analysis | Pérez-Alba E, Nuzzolo-Shihadeh L, Aguirre-García G, Espinosa-Mora J, Lecona-García J, Flores-Pérez R et al. | Mexico | Retrospective Cohort Study | *Good* |
| 12 | The Effect Of Baricitinib Usage On The Clinical And Biochemical Profiles Of Covid-19 Patients- A Retrospective Observational Study | Amarnath A; Das A | India | Observational Retrospective Cohort Study | *Fair* |
| 13 | Anakinra versus Baricitinib: Different Strategies for Patients Hospitalized with COVID-19 | García-García J, Pérez-Quintana M, Ramos-Giráldez C, Cebrián-González I, Martín-Ponce M, del Valle-Villagrán J, et al | Spain | Retrospective Cohort Study | *Good* |
| 14 | Baricitinib improves respiratory function in patients treated with corticosteroids for SARS-CoV-2 pneumonia: an observational cohort study | Rodríguez-García J, Sanchez-Nievas G, Arevalo-Serrano J, Garcia-Gomez C, Jimenez-Vizuite J, Martinez-Alfaro E. | Spain | Observational Cohort Study | *Good* |
| 15 | Baricitinib against severe COVID-19: effectiveness and safety in hospitalised pretreated patients | Iglesias Gómez R, Méndez R, Palanques-Pastor T, Ballesta-López O, Borrás Almenar C, Megías Vericat J et al. | Spain | Observational Retrospective Cohort Study | *Fair* |
| 16 | Baricitinib reduces 30-day mortality in older adults with moderate-to-severe COVID-19 pneumonia | Abizanda P, Calbo Mayo J, Mas Romero M, Cortés Zamora E, Tabernero Sahuquillo M, Romero Rizos L et al. | Spain | Retrospective Cohort Study | *Good* |
| **Interventional Studies** |  |  |  |  | **(Risk of Bias - RoB.2 tool)** |
| 17 | Efficacy and safety of baricitinib in patients with COVID-19 infection: Results from the randomised, double-blind, placebo- | Marconi VC, Ramanan AV, Bono S, Kartman CE, | Across South and North America, | Double-blinded RCT | *Low* |
|  | controlled, parallel-group COV-BARRIER phase 3 trial. | Krishnan V, Liao R, et al. | Europe, and Asia. |  |  |
| 18 | Baricitinib plus Remdesivir for Hospitalized Adults with Covid-19. | Kalil AC, Patterson TF, Mehta AK, Tomashek KM, Wolfe CR, Ghazaryan V, et al. | Across U.S., Singapore, South Korea, Mexico, Japan, Spain, the United Kingdom, and Denmark | Double-blinded RCT | *Low* |
| 19 | Clinical impact of combination therapy with baricitinib, remdesivir, and dexamethasone in patients with severe COVID-19 | Izumo T, Kuse N, Awano N, Tone M, Sakamoto K, Takada K et al. | Japan | Double-blinded RCT | *Low* |

### Observational Studies

Hasan MJ *et al* (20), in a prospective cohort study conducted in Bangladesh, included 238 patients with severe COVID-19 pneumonia, of whom 122 patients were given baricitinib 8 mg (high dose) and 116 patients given 4 mg (usual dose) orally for 14 days by simple random sampling, along with dexamethasone, remdesivir and enoxaparin/dalteparin (in both groups). The time to reach target oxygen saturation of ≥ 94% (5 vs 8 days; IQR: 4–5 and 6–9 days respectively, *P* < 0.05) and no supplemental oxygen demand were significantly lower in the high dose group. Similarly, the requirement of intensive care unit (ICU) and intubation support was significantly lesser in the patients receiving high dose compared to those receiving usual dose of baricitinib (9% vs 17.2%, *P* = 0.02; 4.1% vs 9%, *P* = 0.001 respectively), as was the 30-day all-cause mortality (3.3% vs 6%, *P* = 0.001). The median length of hospitalization (11 vs 13 days; IQR: 9.5–14 and 10–17.5 days respectively, *P* = 0.072) was also lesser in the high dose group. Thrombocytosis and mouth sores were also significantly more among patients in the 8 mg dose group compared to the 4 mg dose group (9.8% vs 2.6 and 2.4% vs 0.8% respectively). So, it was deduced that high dose baricitinib treatment provided significant survival and other clinical benefits among patients of severe COVID-19 at an increased risk of thrombocytopenia and mouth sores. However, there was no control (standard treatment) group in the study and there were some significant differences in the baseline clinical and biochemical parameters of the patients in the two groups.

A prospective study involving 37 adult patients was conducted by the same investigators, 17 of whom were allocated via simple random sampling to the ‘control’ (No Loading Dose, NLD) group and 20 to the ‘Case’ (Loading dose, LD) group, where they received an additional 8 mg loading dose of baricitinib orally before receiving treatment of 4 mg daily oral baricitinib for 14 days which was common to both groups. Patients of both groups tolerated baricitinib treatment well and showed no significant differences in mortality. However, the median time (in days) to reach SpO_2_ > 95% was quicker in the LD group as compared to NLD [3 (IQR:2-8), vs 4 (IQR:4-5), respectively, *P*=0.180]. The same trend favouring baricitinib LD group was seen for the median time to return to normal breathing [(LD, 5(4–5) vs NLD,8 (7–10)], and the median time of hospitalisation [(LD,12): NLD (15)]. The need for ICU support and mechanical ventilation was higher in the NLD group [29.4% (NLD) vs 10% (LD), *P*<0.05 and 11.8% (NLD) vs 5% (LD), p=0.141 respectively]. The major limitations however were the small sample size, no ‘control group, and no justification for the usage of a loading dose. The authors seem to have used the term ‘case control study’ for a study which was actually conducted as a cohort study (21).

The retrospective cohort study conducted by Titanji BK *et al* (22), in Atlanta, U.S. analysed the effect of combination of oral baricitinib (2-4 mg OD for a maximum of 7 days) and oral hydroxychloroquine (200-400 mg OD) along with standard prophylactic treatment of DVT with unfractionated heparin or LMWH in 15 patients with moderate-severe COVID-19 pneumonia. Of these, 11 patients further received antimicrobials. The patients were followed until recovery or death. The study revealed clinical improvement in 73% patients on baricitinib (with hydroxychloroquine) therapy in terms of fever and other presenting symptoms, oxygen requirement, CRP levels with 80% survival (12/15); three patients died due to underlying pre-existing conditions. The study could not provide a cause-effect relationship of the baricitinib drug therapy due to a small sample size, being retrospective, having no comparator, and concomitant hydroxychloroquine administration.

Bronte V *et al* (23), conducted a prospective observational study in Verona, Italy which included 76 patients with COVID-19 pneumonia, of whom 20 were given an initial loading dose of 4 mg BD baricitinib for two days followed by maintenance dose of 4 mg OD for 5 days, in addition to the standard therapy of hydroxychloroquine and/or antivirals (ritonavir/lopinavir). Supportive therapy of antimicrobials and anticoagulants was also administered to the cohorts based on clinician discretion. The mortality after was significantly lower in the baricitinib cohort compared to the standard therapy cohort [1(5%) vs 25(45%) respectively]. ISimilarly, the need for oxygen flow therapy was also lesser (*P*<0.001), and serum levels of C-Reactive Protein (*P*<0.001), p-STAT3 levels (*P*<0.001), as well as T lymphocytes, NK cells, monocytes, Interleukin (IL)- β, IL-6 and TNF-α plasma concentrations were significantly reduced in the baricitinib cohort.

There was also an increase in IgG antibodies in the baricitinib group, for receptor binding domain of SARS-CoV-2 spike protein, increase in P/F ratio (P=0.02), an absolute increase in circulating lymphocytes, IL-8 serum levels, CD4**^+^** T cells [a drastic increase in an effector memory phenotype, (CD3+CD4+CD45RA–CD27–)]. However, both cohorts faired the same with no significant differences in ARDS incidence, hospitalization duration, fever resolution, lung involvement as on HR-CT/ Chest X ray, CD8**^+^** T cells, the absolute number of NK (Natural Killer) cells, expansion of monocytes. So, it was deduced that baricitinib treatment provided significant survival and other clinical benefits among patients of COVID-19 pneumonia. The presence of missing data and a short follow up duration were some limitations of the study.

Falcone M *et al* (24), in a prospective cohort study (with 1:1 matching) conducted in Pisa, Italy, primarily evaluated the effect of LMWH treatment in 244 patients with COVID-19 pneumonia along with secondarily assessing treatment effects in patients treated with hydroxychloroquine (238), proteases inhibitors (201), doxycycline (150), steroids (141), macrolides (42), baricitinib (40), tocilizumab (13), remdesivir (13), and corticosteroids (141). Although 40 patients received baricitinib along with one or more additional drugs i.e. doxycycline (19), proteases inhibitors [Lopinavir/ritonavir or Darunavir/ritonavir] (16), hydroxychloroquine (19), steroids (15), LMWH (20) etc., only 21 of these had matched controls and were evaluated for outcome. The mortality of patients treated with baricitinib was 9% (n=5, *P*=0.113), the univariate and multivariate regression analysis for the association between interventions and mortality revealed a hazards ratio of 0.14 (*P*=0.006) and an aHR of 0.69 (*P*=0.45), and hazards ratio for intervention and death/severe ARDS was 0.53 (*P*=0.12). Yet, when intervention with baricitinib was matched with mortality and severe ARDS, and regression analysis performed, the HRs were 1.35 (0.32-5.96) and 1.38 (0.48-4.01) with *P* values of 0.678 and 0.549 respectively; both results were not statistically significant. Thus, the study does not show any significant beneficial (or deleterious) effect of baricitinib in the clinical recovery of patients suffering from COVID-19 pneumonia. The study was clearly underpowered to assess effect of baricitinib.

Rosas J *et al* (25), in their retrospective cohort study aimed to investigate the effect of baricitinib and tocilizumab as monotherapy or in combination in patients with interstitial pneumonia due to SARS-CoV-2 in 60 hospitalised patients in Spain. The patients were categorised as baricitinib monotherapy (12), tocilizumab monotherapy (20), a combination of both (11) and neither (17), whereas the other antivirals, azithromycin, interferons and corticosteroids were given based on the clinician’s discretion alone. The study revealed no significant mortality benefits of either of the treatment regimens and no relevant side effects (such as thrombotic symptoms, herpes zoster, leukopenia, thrombopenia, or significant alteration of blood transaminases) of baricitinib treatment. The baricitinib monotherapy group showed a significant (*P*<0.05) reduction in temperature, CRP, D Dimer, oxygen saturation and respiratory rate as compared to standard of care group (neither tocilizumab or baricitinib), and similar but more significant differences (*P*<0.01) were seen between the combination group (tocilizumab + baricitinib) and standard of care group. The comparison of baricitinib vs tocilizumab monotherapy groups indicated greater reductions in CRP, D-Dimer, respiratory rate and lesser lymphocytosis, LDH change and temperature change in the baricitinib group. Baseline comparison revealed that the patients receiving baricitinib/tocilizumab treatment presented with a more serious (worsened) PaO2/FiO2 level in comparison to those who received neither drug, which indicated a selection bias.

The retrospective longitudinal study by Milligan PS *et al* (26), conducted in Indianapolis, U.S., analysed the effect of baricitinib (2 mg, OD, PO for 7 days) along with the standard of care (hydroxychloroquine ± ribavirin ± corticosteroids and anticoagulant treatment) in 22 patients with COVID-19 pneumonia. There was a marked difference in clinical and biochemical baseline characteristics among the participants, possibly due to selection and Berksonian bias. The study revealed a non-universal clinical improvement in patients on baricitinib therapy - 16 patients did not have progression of the disease, while the remaining died (n=3) or progressed to need mechanical ventilation. In the majority of the patients (77.3%), oxygen demands improved by at least one WHO oxygenation criteria prior to completion of the baricitinib regimen. Ferritin and CRP values decreased from median 885 (419–2330) ng/mL to 326.5 ng/mL and 20.35 (8.5-2.75) mg/dL to 17.3mg/dL respectively. Baricitinib also showed side-effects [Thrombocytosis (n=22), Transaminase elevation (n=5), Acute Kidney Injury, AKI (n=2), lymphopenia (n=1), *Staphylococcus aureus* bacteraemia (n=1)] which were mostly mild; only AKI, Lymphopenia and *Staphylococcus aureus* bacteraemia led to treatment discontinuation. However, the study had several limitations including but not limited to its retrospective nature, incomplete data, lack of long term follow-up, and absence of comparators.

Cantini *et al* (12), performed a pilot ambivalent cohort study in Italy where 12 patients suffering from moderate COVID-19 were recruited to receive baricitinib therapy apart from standard therapy of antivirals and hydroxychloroquine. From previous hospital records, another 12 patients were matched to serve as controls as they did not receive baricitinib but received the then-standard of care therapy. Overall, in the baricitinib treated group, clinical characteristics and respiratory function parameters significantly improved consistently compared to baseline. CRP values significantly decreased at 14 days whereas no significant changes were observed in the controls. Fever, SpO_2_, PaO_2_/FiO_2_, CRP, and Modified Early Warning Score (MEWS) significantly improved in the baricitinib-treated group compared with controls (*P*= 0.000; 0.000; 0.017; 0.023; 0.016, respectively). ICU transfer was requested in 33% (4/12) of controls and in none of the baricitinib treated patients (*P*=0.093). Discharge at 14 days occurred in 58% (7/12) of the baricitinib-treated patients vs 8% (1/12) of controls (*P*= 0.027). At discharge, 57% (4/7) of the baricitinib group had a negative viral nasal/oral swab. The study, while indicating a beneficial impact of baricitinib treatment, also has significant limitations such as different treatment timeframes and nature of the cohorts and substantially low sample size of the study.

The same group of investigators also performed a retrospective longitudinal multicentric study in consecutive hospitalised patients with moderate COVID-19 symptoms, to evaluate effectiveness of baricitinib given over 2 weeks (as add on to standard therapy of antivirals lopinavir/ritonavir, LMWH) in 113 patients as compared to standard therapy in 78 patients. Comparison of baseline characteristics revealed no significant differences. The two-week case-fatality rate was found to be significantly lower in the baricitinib group patients as compared to the control group [0% vs 6.4% in baricitinib vs control, *P*= 0.01]. Similarly, lesser ICU admission was also seen [0.88% vs 17.9%]. When analysed at the end of each week, the SpO_2_, oxygen partial pressure/FiO_2_, transaminase levels, lymphocyte counts were significantly increased while CRP and IL-6 levels were significantly reduced in the baricitinib group. As compared to the control group, at discharge, the baricitinib group showed a significant reduction in the proportion of patients who tested positive for pharyngeal swabs for COVID-19 [12.5% vs 40%] (13).

Santol CS *et al* (27), in their observational study retrospectively analysed the incidence of COVID-19 pneumonia in patients being treated with disease-modifying anti-rheumatic drugs (DMARDs). Out of 820 patients with rheumatic diseases, 40 contracted COVID. Although 3 out of the 820 patients were being treated with baricitinib, none of them contracted COVID, indicating a possible protective benefit of the drug. However, the very low numbers of COVID-19 cases and patients receiving baricitinib were major limitations of the study.

In a retrospective cohort analysis done by Alba EP *et al*, in Mexico (28), baricitinib in combination with dexamethasone (n = 123) was compared to dexamethasone alone (n = 74) among hospitalised patients with severe COVID-19; having SpO2 < 93% and bilateral pulmonary infiltrates. The primary outcomes was overall 30 day mortality which was found to be significantly lower in the combination therapy group (20.3%) as compared to the dexamethasone monotherapy group (40.5 %); p < 0.05. The risk of hospital acquired infections was not increased by combination therapy. Another pre published small retrospective analysis from India (29) described the clinical and biochemical parameters of 31 patients with moderate – severe receiving a combination of barcitinib and remdesivir and showed that there was a significant reduction (p < 0.05) in the oxygen requirement, and levels of CRP, IL-6 over 7 days among patients post initiation of therapy. The mortality rate in the study was 9.6% (3/31); patients who died were elderly and had co- morbidities.

Another retrospective cohort study conducted by Garcia-Garcia JA *et al* (30), in Spain evaluated the effect of treatment with IL-1 inhibitor anakinra (n = 125) versus baricitinib (n = 217) as add on to background corticosteroid therapy and standard care on the survival of hospitalised patients with COVID-19 pneumonia, and found that there were no differences in mortality between the two groups (17.6% vs 16.6%). However, significantly lesser number of patients receiving baricitinib required invasive mechanical ventilation (4.6%) compared to those on anakinra (10.4%); p = 0.039.

Garcia-Rodriguez JL *et al* (31), recently published a prospective cohort study comparing improvement in pulmonary function among patients with moderate – severe COVID-19 pneumonia treated with corticosteroids (n = 50) versus corticosteroids and baricitinib (n = 62); both in addition to lopinavir / ritonavir and hydroxychloroquine. Results showed a significantly larger improvement (p < 0.001) in SpO2 / FiO2 in the combination group compared to corticosteroids alone (mean difference 49; 95% CI 22, 77). Similarly, a significantly lower proportion of patients required supplemental oxygen in the combination group at discharge and 1 month (25.8%, 12.9%) compared to those in the corticosteroids alone group (62%, 28%); with OR’s of 0.18 and 0.31 respectively at the two time points.

Gómez *et al* (*32*), conducted an observational retrospective study in Spain involving 43 patients hospitalized for severe COVID-19 (SpO2 < 92%) who received 4 mg baricitinib orally for 5-7 days; out of these 84% (36/43) also received corticosteroids. A significant median clinical improvement of 3 points (IQR 1–4) was observed on an 8-category ordinal scale from day 1 to day 15 (p < 0.01). The median time of recovery was 12 days, all patients survived by day 30 and day 60; all participants were discharged from hospital due to clinical improvement. There was a significant improvement (p < 0.05) in all the clinical parameters related to poor prognosis of COVID-19 while no adverse events of interest like respiratory tract infections, reactivation of latent infections or creatinine changes were reported.

Abizanda *et al* (33), in their retrospective cohort study recruited 328 older adults with moderate to severe COVID-19 pneumonia (n =164 each on baricitinib and propensity score matched controls). They were categorized into two age brackets: <70 years (n = 86) and ≥ 70 years (n = 78) old, and mean total dose of baricitinib received was 17.6 mg ± 10.2 for a mean 5.9 days of treatment. Few of the participants also received tocilizumab, anakinra, and corticosteroids. Patients in the ≥ 70 age group showed significantly lower 30-day fatality rate with baricitinib when analysed using Cox proportional hazard models adjusted for various important covariates (HR 0.21; 95% CI 0.09–0.47; p < 0.001). Similar results were also found in those younger than 70 (HR 0.14; 95% CI 0.03–0.64; p = 0.011) despite a higher disease severity in the baricitinib group. This study demonstrated the clinical utility and safety of baricitinib in elderly patients for whom trial results or separate analyses have not been published.

### Interventional Studies

Marconi *et al* (34), have reported the results of COV BARRIER, a phase 3 randomized, double blinded, placebo-controlled trial, which enrolled 1525 adult hospitalized symptomatic patients with COVID-19 requiring low flow oxygen and having ≥1 elevated inflammatory marker (C reactive protein, D-dimer, lactate dehydrogenase, ferritin). This study was conducted at 101 centres across 12 countries in South and North America, Europe, and Asia. Patients were randomized in a ratio of 1:1 to receive oral baricitinib 4 mg (n=764) or placebo once daily for 14 days, in addition to standard treatment (SOC – including low dose systemic corticosteroids – 79.3%, mainly dexamethasone, remdesivir – 18.9%, and anticoagulants). All patients were followed for progression of disease and mortality till day 28 and additionally for mortality up to day 60.

On day 28, no statistically significant difference was seen in the proportion of patients who progressed (requiring high flow oxygen, non-invasive / invasive ventilation, or death) upon treatment with baricitinib versus placebo, in addition to SOC [27.8% vs 30.5%; odds ratio (OR) 0.85, 95% confidence interval (CI) 0.67- 1.08; p=0.18). However, it was noted that baricitinib reduced the rate of all-cause mortality significantly compared to placebo in a prespecified secondary endpoint analysis. On day 28, 8.1% mortality was seen in patients who received baricitinib plus standard treatment compared to 13.1% seen in those who received placebo plus standard treatment (Hazard ration, HR 0.57, 95% CI 0.41 – 0.78, p = 0.0018), corresponding to a reduction in mortality rate by 38.2%. Similar significant reduction was seen in long term mortality on day 60 (HR 0.62, 95% CI 0.47 – 0.83, p = 0.005). The mortality benefits were more pronounced among patient subgroups with higher baseline severity. In terms of safety analysis, there were no significant differences between the baricitinib and placebo groups for treatment emergent adverse events (44.5% vs 44.4%), serious adverse events (14.7% vs 18%) or serious infections or venous thromboembolic events. The study showed synergistic effect of baricitinib with corticosteroid therapy to reduce mortality among hospitalized COVID-19 patients.

In May 2020, results for first stage of Adaptive COVID-19 Treatment Trial (ACTT-1) showing the effectiveness of remdesivir treatment in hospitalized COVID-19 patients with pneumonia were released (35). Kalil *et al* (36) designed ACTT-2 aiming to evaluate the efficacy of remdesivir in combination with baricitinib compared to remdesivir alone plus placebo (control) as treatment in 1033 hospitalized adults with COVID-19; 515 were assigned to the combination group. The patients received a loading dose of 200 mg remdesivir on day 1 intravenously followed by a maintenance dose of 100 mg from day 2 - 10 or until hospital discharge/death, and baricitinib was administered at a daily dose of 4 mg for 14 days or until hospital discharge/death.

Patients in the remdesivir and baricitinib combination group recovered a day faster compared to the ones who received remdesivir alone (median 7 days vs 8 days; rate ratio, RR for recovery, 1.16; 95% CI 1.01 to 1.32; P=0.03). Median time to recovery for patients receiving high flow oxygen or noninvasive ventilation (baseline score of 6) in the combination group compared to control was 10 days vs 18 days (RR for recovery 1.51; 95% CI 1.10 to 2.08). For the patients who received no oxygen (baseline score 4), supplemental oxygen (baseline score 5) and mechanical ventilation (baseline score 7), the rate ratio of recovery was 0.88 (95% CI, 0.63 to 1.23), 1.17 (95% CI, 0.98 to 1.39) and 1.08 (95% CI, 0.59 to 1.97) respectively. The overall mortality rate after 28 days of randomization in combination group was 5.1% compared to 7.8% in the control group (HR 0.65; 95% CI, 0.39 to 1.09). Significant difference in mortality was observed in patients who received supplemental oxygen (1.9% vs. 4.7%; hazard ratio, 0.40; 95% CI, 0.14 to 1.14) and those who received high flow oxygen or noninvasive ventilation (7.5% vs. 12.9%; hazard ratio, 0.55; 95% CI, 0.22 to 1.38) respectively. The time taken for recovery in combination group patients was 6 days vs 8 days in control group patients (rate ratio, 1.21; 95% CI, 1.06 to 1.39). Similar trend was seen in discharged patients; 6 days vs 7 days in respective groups (rate ratio, 1.24; 95% CI, 1.07 to 1.44). A total of 207 patients (40.7%) in combination group exhibited grade 3 or 4 adverse events (anemia, hyperglycemia, decreased lymphocyte count, and acute kidney injury) compared to 238 patients (46.8%) in the control group. Out of these, 81 patients (16.0%) had episodes of serious adverse events compared to 107 patients (21.0%) in combination and control group respectively with a difference of -5.0 percentage point between the two groups (95% CI, −9.8 to −0.3; P = 0.03). This study shows that baricitinib in combination with remdesivir is not only superior and safe with fewer serious adverse events compared to remdesivir alone, but it also helps reducing the time taken by the patients to recover and accelerating further improvements in their clinical status especially for those who received high flow oxygen or noninvasive ventilation.

Izumo *et al* (37), conducted a small interventional study on 44 severely or critically affected COVID-19 patients without severe renal dysfunction at the Japanese Red Cross Medical Center, Tokyo, who were administered a triple combination therapy of oral baricitinib 4 mg (upto 14 days), IV remdesivir (in standard dosing regimen upto 10 days) and IV dexamethasone 6 mg (upto 10 days). Overall results showed effectiveness of this combination therapy (BRD) in terms of low mortality rate (2.3%) and not requiring invasive mechanical ventilation (90%). The median duration of hospitalization and ICU stay was 11 days and 6 days respectively whereas the median time of recovery was 9 days The incidence of adverse events was 34% (15/44). This was the first published study demonstrating the safety and efficacy of the triple combination therapy of BRD but the lack of any comparator group is the major limitation.

## Discussion

For the past 20 months, the COVID-19 pandemic has presented the most challenging conditions seen in a century for the clinicians across the globe due to its infectivity, morbidity and mortality, characterized by the spread of lethal pneumonia in up to 15–20% of the cases (8). Despite over an year’s intensive research efforts, there are still no exact courses of treatment for hospitalized or non-hospitalized patients. The current therapeutic strategies are focused on the use of anti-inflammatory agents like corticosteroids, IL-6 inhibitors and antivirals agents like remdesivir in combination with standard supportive care. The need for more therapeutic options, proven as effective therapies still remains, particularly for the hospitalized subset of patients suffering from moderate – severe COVID-19 who are at risk of progressing to life endangering complications like ARDS and multiorgan involvement. Amidst these conditions, baricitinib, an oral janus kinase inhibitor that inhibits cytokine release thereby facilitating cellular differentiation, proliferation and survival, was emergency approval (EUA) by the US FDA in November 2020 to be used for hospitalised patients with moderate to severe COVID- 19 pneumonia as part of a combination regimen. A daily dose of 4 mg in combination with standard dose of remdesivir plus supporting therapy was added to the treatment guidelines. Subsequently, in July 2021, the FDA has revised this EUA to allow use of baricitinib as monotherapy (without the need for combination with remdesivir) for treatment of COVID-19 in hospitalized adults and pediatric patients 2 years of age or older requiring supplemental oxygen, non-invasive or invasive mechanical ventilation, or ECMO (5).

In the present systematic review, we attempted to unearth evidence and possible benefits – both available and upcoming, of baricitinib as a potential anti-inflammatory agent for treatment of COVID-19. Data extracted from published observational and interventional studies showed promising immunomodulatory and anti-inflammatory results of baricitinib as a standalone treatment along with standard care or in combination with other drugs, in improving clinical, biochemical and other related disease parameters in patients with moderate – severe COVID-19. Due to the continued immense global need for clinical care and treatment of the disease particularly in those hospitalised COVID-19, a lot of the evidence has actually come from observational studies on different drugs being tried in patients admitted in the hospitals due to COVID-19, rather than the hierarchical gold standard - randomised, blinded controlled trials. We could find a similar evidence base for baricitinib also where published observational studies of different types outnumbered the published interventional trials. Though the quality of evidence may be lower via observational studies (or NRSIs), the sheer number of parallel conclusions, supported by the evidence from RCTs, albeit limited at present, point to a substantial possibility of benefits which may not merely be due to chance. This could prove to be further significant, as evidence from more ongoing RCTs gets published.

Multiple studies have been attempted to investigate the same question – whether baricitinib provide any clinical benefit to hospitalised patients diagnosed with COVID-19? Some of these studies compare the efficacy of baricitinib alone to the standard of care for COVID-19 patients (antivirals, immunomodulators, protease inhibitors etc.), with different dosing regimens or loading dosages, or as a part of a multi-drug pharmacological treatment regimens to analyse the comparative efficacy and safety between baricitinib and different comparators. Despite the considerable heterogeneity in terms of study populations, sites, study designs, treatment regimens, significant individual limitations, as well as the differences in summary measures, the evidence available does largely provide a common pattern in favour of utility of the drug in COVID-19, supported by satisfactory safety of treatment. Further studies, particularly the multiple ongoing RCTs, should be able to provide more definitive evidence in all regards.

Among the eleven observational studies, including 9 cohort/cross-sectional and two case control studies, most of the results appear to point towards a similar pattern favouring beneficial improvements of key clinical outcomes and/or biochemical or laboratory parameters with baricitinib therapy in moderate to severe COVID-19 hospitalised patients, generally used in addition to standard of care. Observational studies also compared baricitinib to other drugs such as tocilizumab (25), used various doses of baricitinib (20, 31) assessed therapeutic effects of loading dose changes h or combination with other drugs such as tocilizumab (21), LMWHs like enoxaparin (22–26,31) etc., to investigate outcome parameters such as mortality, improvement in oxygen saturation, hospital stay, need for intensification of treatment like ventilatory requirement, virological clearance and inflammatory biochemical markers like CRP. Most of the above permutations of baricitinib interventions and outcomes depicted results favour of baricitinib, through benefits in alleviating the clinical, biochemical and other disease parameters of COVID-19.However, in some studies, reduction in mortality was not shown to be significant due to baricitinib treatment (21,24,25). It is also important to note that some studies also show increased incidence of adverse drug reactions as mouth sores (20), infections (26), thrombocytopenia (20, 25) during the treatment or at the end of treatment with baricitinib. But no study showed an inferiority of baricitinib in the treatment or clinical aid, and a non-inferiority was also depicted in studies between the varying doses of baricitinib when compared (25). This may inherently be due to the short duration of treatment and/or follow up period along with limited sample sizes to making any definite conclusions. A major drawback of these studies is the inherent nature of the studies, being observational, which introduces numerous methodological limitations and flaws along with biases. This inherently gives rise to two sets of bias, namely the selection bias and confounding errors due to the lack of randomisation, which was managed to a certain level in some studies, yet many failed to do the same. When graded by the study quality assessment tool for observational cohort and cross-sectional studies (19), a validated tool for assessing quality of observational studies, eight studies ranked “fair” and nine of the them ranked “good”. The details of assessment of each study is available in the supplementary data 1.

Results of two clinical trials could be found published at the time of conduct of search for this review. Among these, the ACTT-1 results led to the grant of EUA for baricitinib to be used in combination with remdesivir in hospitalised COVID-19 patients. The trial showed that participants who received baricitinib plus remdesivir (along with standard care of treatment) showed significant improvement in median time of recovery compared to control group subjects (6 days vs 8 days, rate ratio 1.21; 95% CI, 1.06 to 1.39), with a 5.1% mortality rate in combination group compared to 7.8% in control (hazard ratio 0.65; 95% CI, 0.39 to 1.09) (36). Similar trend in reduction in mortality by 38.2% was observed by Marconi *et al* in the experimental arm of oral baricitinib 4 mg plus standard treatment compared to control who received placebo in addition to standard treatment (8.1% vs 13.1%, HR 0.57, 95% CI 0.41 – 0.78, p = 0.0018) (34). There was also improvement in participants’ requirement for oxygen therapy especially in those who received high flow oxygen or for non-invasive ventilation (7.5% vs. 12.9%; HR, 0.55; 95% CI, 0.22 to 1.38). Encouragingly, fewer episodes of severe adverse events were seen in participants belonging to the baricitinib group compared to the standard of care. The risk of bias was assessed using the risk of bias (ROB 2.0) tool (38). Both the studies were graded having overall ‘low risk of bias’. Detailed assessment is available in the supplementary data 2.

### Ongoing Clinical Trials

In addition to the published or pre published studies, it is also important to review the key ongoing studies (registered clinical trials) on the use of baricitinib in COVID-19 as well to understand the kind of evidence that is expected to be generated in coming times. Currently, there are 11 ongoing clinical trials for assessing the safety and efficacy of oral baricitinib alone or in combination therapy for treating hospitalized patients with COVID-19.

Table 2 enlists such clinical trials, mostly multicentric RCTs, found registered on various global clinical trial registries whose findings are yet to be published. Out of the 11 trials, one trial is in phase IV, two each are in phase III and phase II/III, five are in phase II and one is in phase Ib/II respectively.. One trial was terminated since similar results from another trial were released and one trial was withdrawn as it failed to make changes requested by the FDA.

**Table 2:** Registered on-going clinical trials to evaluate the efficacy and safety of baricitinib in COVID-19 patients

| **S. No** | **Trial identifier/**<br>**registry ID and phase** | **Trial setting -**<br>**Patient**<br>**population** | **Last updated status** | **Official title** | **Sponsor/lead**<br>**institution,**<br>**Country** | **Starting date –**<br>**Completion date (Primary**<br>**/ Study)** |
| --- | --- | --- | --- | --- | --- | --- |
| 1 | NCT04390464 (39)<br>Phase- IV | Hospitalised | Recruiting<br>(Updated on: May 20,<br>2020) | mulTi-Arm Therapeutic study in pre-ICu patients admitted<br>with Covid-19 – Repurposed Drugs (TACTIC-R) | Cambridge University<br>Hospitals NHS<br>Foundation Trust<br>United Kingdom | May 8, 2020<br>May 7, 2021/<br>May 1, 2022 |
| 2 | NCT04640168 (40)<br>Phase- III | Hospitalised | Completed<br>(Results not yet<br>published)<br>(Updated on: November<br>17, 2021) | A Multicenter, Adaptive, Randomized Blinded Controlled<br>Trial of the Safety and Efficacy of Investigational<br>Therapeutics for the Treatment of COVID-19 in<br>Hospitalized Adults (ACTT-4) | National Institute of<br>Allergy and Infectious<br>Diseases (NIAID)<br>U.S. | November 23, 2020<br>May 18, 2021/<br>June 18, 2021 |
| 3 | NCT04832880 (41,42)<br>Phase- III | Hospitalised | Not yet recruiting<br>(Updated on: April 6,<br>2021) | Factorial, Multicentric, Randomized Clinical Trial of<br>Remdesivir and Immunotherapy in Combination With<br>Dexamethasone for Moderate COVID-19 (the<br>AMMURAVID Trial) | ASST Fatebenefratelli<br>Sacco<br>Italy | April 6, 2021<br>March 2022/ December<br>2022 |
| 4 | NCT04320277 (43)<br>Phase- II/III | Hospitalised | Not yet recruiting<br>(Updated on: April 22,<br>2020) | Baricitinib Combined with Antiviral Therapy in<br>Symptomatic Patients Infected by COVID-19: an Open-label, Pilot Study | Hospital of Prato<br>Italy | May 16, 2020<br>June 30, 2020/ July 30, 2020 |
| 5 | NCT04340232 (44)<br>Phase- II/III | Hospitalised | Withdrawn (Could not<br>make FDA mandated<br>changes)<br>(Updated on: March 8,<br>2021) | Safety and Efficacy of Baricitinib for COVID-19 | University of Colorado,<br>Denver<br>U.S. | March 2021<br>August 2021/ October 2021 |
| 6 | NCT04399798 (45) | Hospitalised | Not yet recruiting | A proof-of Concept Study of the Use of Janus Kinase 1 and<br>2 Inhibitor, Baricitinib, in the Treatment of COVID-19- | IRCCS Policlinico S.<br>Matteo | May 15, 2020 |
|  | Phase- II |  | (Updated on: May 22, 2020) | related Pneumonia | Italy | September 15, 2020/<br>November 15, 2020 |
| 7 | NCT04393051 (46)<br>Phase- II | Hospitalised | Not yet recruiting<br>(Updated on: May 20, 2020) | BARICIVID-19 STUDY: Multicentre, Randomised, Phase IIa Clinical Trial Evaluating Efficacy and Tolerability of Baricitinib as add-on Treatment of In-patients With COVID-19 Compared to Standard Therapy | Azienda Ospedaliero, Universitaria Pisana<br>Italy | May 20, 2020<br>June 30, 2020/<br>July 30, 2020 |
| 8 | NCT04373044 (47)<br>Phase- II | Hospitalised | Terminated (The study was terminated after the release of results of ACTT-2 (NCT04401579).)<br>(Updated on: May 26, 2021) | A Phase II Randomized Double-Blind Trial of Baricitinib or Placebo Combined With Antiviral Therapy in Patients With Moderate and Severe COVID-19 | University of Southern California<br>U.S. | May 1, 2020<br>May 12, 2021 |
| 9 | NCT04346147 (48)<br>Phase- II | Hospitalised | Active, not recruiting<br>(Updated on: February 10, 2021) | Clinical Trial to Evaluate Efficacy of 3 Types of Treatment in Patients With Pneumonia by COVID-19 (Covid19COVINIB) | Hospital Universitario de Fuenlabrada<br>Spain | May 7, 2020<br>June 2021/ September 2021 |
| 10 | NCT04321993 (49)<br>Phase- II | Hospitalised | Recruiting<br>(Updated on: July 24, 2020) | Treatment of Moderate to Severe Coronavirus Disease (COVID-19) in Hospitalized Patients | Lisa Barrett, Nova Scotia Health Authority<br>Canada | April 17, 2020<br>February 2022/ March 2022 |
| 11 | 20-0356 (50)<br>Phase- Ib/II | Hospitalised | Recruiting<br>(Updated on: October 14, 2020) | A Phase I/II Clinical Trial to evaluate the efficacy of baricitinib to prevent respiratory insufficiency progression in onco-hematological patients affected with COVID19: A structured summary of a study protocol for a randomised controlled trial | Institutional Crowd Funding<br>Spain | December 16, 2020<br>June 2021 |

The latest trial to get underway is a phase IV multi arm platform trial - Therapeutic study in pre-ICU patients admitted with COVID-19 – Repurposed Drugs (TACTIC-R) being conducted at multiple centers across U.K. (39, 51). In this study, 687 to 1167 participants are estimated to be enrolled by the end. It is a randomized, parallel 3-arm (1:1:1 ratio) trial aiming to evaluate the efficacy of baricitinib and ravulizumab as a potential treatment for patients hospitalized due to COVID-19 compared to standard treatment alone over a treatment study period of 14 days with a follow-up at day 28 and day 90 post discharge from hospital. Participants enrolled in the first arm will receive standard of care for COVID-19 patients. The baricitinib arm will receive the dose of 4 mg once a day plus standard of care for 14 days or until discharge. The safety and efficacy of different treatment arms will be assessed on a 7-point pulmonary scale; death, invasive mechanical ventilation, non-invasive ventilation or high flow oxygen, low flow oxygen, no oxygen, discharged but normal activities not resumed and discharged with normal activity resumed.

The NIAID trial – ACTT-4 (40), which has stopped further recruitment in April 2021 (52) was a phase III trial on hospitalized participants with moderate to severe COVID-19 randomized to receive remdesivir plus baricitinib versus remdesivir plus dexamethasone (n= 1500; recruited= 1010). The primary endpoint of this trial was the proportion of subjects not dying or not getting hospitalized on mechanical ventilation i.e. mechanical ventilation free survival by day 29 and there were multiple secondary outcome measures. The interim efficacy analysis showed that neither treatment regimen was likely to be found to be significantly better than the other, and no safety issues were found with either regimen.

The phase III AMMURAVID trial is being conducted across 21 study locations in Italy (41, 42) since April, 2021 and is expected to enrol approximately 4000 participants by the end of December 2022. These participants will be randomly divided into four arms aiming to test and evaluate the efficacy of remdesivir, baricitinib, remdesivir + baricitinib against the control arm (dexamethasone 6 mg IV x 10 days). The main objective of this trial is to reduce progression of severe respiratory failure in hospitalized COVID-19 patients with PaO_2_/FiO_2_ ratio <200 mmHg (ARDS-range), and additionally to assess the effects of these interventional drugs on the immune response markers. In the baricitinib arm, the participants will receive dexamethasone plus 4 mg baricitinib once a day orally for 10 days. The primary end point is the proportion of participants with PaO_2_/FiO_2_ ratio <200 mmHg at day 7,14,21 and 28 in each experimental arm compared to control.

A phase II/III study (BARI-COVID) (43), is an open-label, non-randomized, 2-week trial registered to be conducted at the Hospital of Prato, Italy, estimated to recruit 200 participants to test the efficacy of baricitinib in combination with antiviral therapies in hospitalized patients with mild to moderate COVID-19. All participants in the experimental arm will receive 4 mg baricitinib orally in combination with antiviral therapy - 250 mg of lopinavir/ritonavir once a day for two weeks. The control arm participants will receive the antiviral therapy or hydroxychloroquine for two weeks. The primary endpoint of this study is to compare the differences in proportion of participants requiring ICU admission and to evaluate the values of CRP, IL-6 and TNFα to estimate the worsening of the disease. However, no participants have been recruited so far under this trial.

Another therapeutic trial (BREATH) (45), piloted in Italy, is a single arm open label study to assess the safety and efficacy of baricitinib 4 mg baricitinib orally for 7 days in hospitalized patients with COVID-19 pneumonia. On 8^th^ day, the participants will be assessed based on their need of moderate or severe oxygenation impairment according to Berlin criteria (measured as PaO_2_/FiO_2_) or death. On 15^th^ day, assessment of the participants will be done for median SpO_2_, number, types and severity of adverse events, biological assessments (level of various interleukins, TNF alpha, interferon gamma, viral load etc.) and rate of mortality will be done.

A phase II study, (Baricivid-19) (46), is a multicentre RCT conducted in Pisa, Italy, which recruited 126 patients (not recruiting now) to test the efficacy, safety and tolerability of baricitinib (4 mg orally for 14 days plus standard care) in comparison to the standard of care therapy in hospitalised patients who had SARS-CoV2 pneumonia. The primary endpoint was reduction in invasive ventilation after a week and fortnight of treatment, and other endpoint were parameters such as mortality (at 14, 28 days), time to invasive mechanical ventilation, length of hospital / ICU stays, toxicity etc.

Another phase II study, (Covid19COVINIB) is an open label RCT being conducted in Madrid, Spain (48) which aims to recruit 165 patients to study the efficacy of baricitinib and imatinib in comparison to supportive care therapy in patients who have early SARS-CoV2 pneumonia. Participants will receive 400 mg OD of imatinib and baricitinib 4mg OD orally in the experimental arms whereas control arm will receive therapeutic intervention aiming towards controlling any clinical deterioration. The primary end point is to assess clinical improvements post treatment as per a 7-point ordinal scale by two points at least, and the secondary end point is to measures safety (number of serious adverse effects and premature discontinuation of the treatment) and tolerability (number of participants with treatment related ADRs by CTCAE v5 through the study completion) after 30 days.

The other phase II non-randomized trial is being conducted by Dalhousie University (49) in Canada, anticipating to recruit ∼800 adult patients hospitalized with moderate-severe COVID-19. In the experimental arm of the trial, all the participants will be given 2 mg baricitinib once a day for 10 days whereas the participants in the control arm will receive standard of care. The clinical status of the participants will be evaluated on day 15 on a 7-point ordinal scale. The main objective of this trial is to determine the time taken by the participants to show clinical improvements in terms of their respiratory rate, fever, normal pulmonary function, and oxygen saturation.

A phase Ib/II trial, BARCOVID19 (50), in Barcelona, Spain is expected to recruit 136 participants with the main objective of determining the tolerability of baricitinib in oncohematological patients with COVID-19 in phase I. The phase II of this trial aims to determine the reduction of inflammatory response due to COVID-19, and prevention of development of severe ARDS in oncohematological patients compared to the other treatment received at the discretion of the investigator. The dose of 4 mg of oral baricitinib is being used for 5 – 7 days which may be reduced to 2 mg depending on the observed toxicity. Apart from baricitinib, participants will receive antibiotics as per their clinical requirements, and prophylaxis to manage their thromboembolic disease. Administration of remdesivir or dexamethasone will be considered for specific indications among participants.

Out of these 11 trials, one was withdrawn, and another was terminated. A single arm, open label phase II/III trial to evaluate the effect of baricitinib on the severity and progression of COVID-19 (44) was withdrawn as the responsible party failed to make the necessary changes as instructed by the U.S. FDA. The participants were proposed to be given 2 mg baricitinib once a day for 14 days. Similarly, a phase II trial proposed by University of Southern California in collaboration with National Cancer Institute (47) was terminated as results from another similar study were published (ACTT-2; trial identifier number- NCT04401579).

These ongoing clinical trials are expected to be completed in coming months (Table 2) and they comprise of different phases, both randomized and non-randomized, single and multicentric studies being conducted in different countries, employing different dosages of experimental drugs, as well as using varying timepoints or intervals related to onset of symptoms, disease severities of COVID-19, and different outcome measures. This may lead to conflicting results in the future. The table of comparison provides a clearer view of all important specified summary measures. However, the strength of such evidence is likely to be strong considering the well-defined methodologies of such studies. Interestingly, a phase IV trial has been launched in the UK (on the TACTIC-R platform) which includes baricitinib as one of the treatment arms, but baricitinib is still under review for approval by the EMA for its use in COVID-19 (6) and its approval for COVID-19 in UK could not be found. The drug has however been recently approved in March 2021 for moderate to severe atopic dermatitis by NICE (53). Grave concerns are being raised worldwide currently regarding the new ‘Omicron’ variant of SARS CoV- 2 which has been declared as a variant of concern (VOC) by the WHO. Current knowledge about this variant is limited in terms of its transmissibility, severity of disease as well as effectiveness of currently available vaccines and treatments including the newer treatments like molnupiravir (54). It has raised fresh question marks over the adequacy of treatment options available for management of COVID-19, for all grades of disease – mild to severe, and the world is gripped with the fear of new waves of the pandemic again.

In terms of the current status of baricitinib in the recommendations for COVID-19 treatment, the US NIH guidelines include baricitinib as an add on treatment option to corticosteroids and/or remdesivir for hospitalised patients requiring high flow oxygen or noninvasive / invasive ventilation or ECMO (55). However, baricitinib does not find a place as of now in the treatment recommendations / guidelines for COVID-19 from the WHO, NICE or in countries like India where the drug has actually received an emergency approval like the U.S. (56, 57).

## Limitations

Despite using the comprehensive WHO database of COVID research for the literature search, which is a collection of most of the recognised electronic databases globally, there may still have been omissions especially from areas like grey or non-published literature. Publications in only English language were considered for eligibility which could have led to a language or geographical bias. Majority of the studies finally selected for the review were observational - Cohort/Cross Sectional (14) and Case Controls (2), whereas only three published interventional studies could be found in the designated period of search. In the hierarchy of evidence, this systematic review thus provides relatively lower level of evidence. The results of multiple clinical trials are not yet available as these are still ongoing.

We could not perform a meta-analysis as most of the evidence is in the form of non-randomised studies of interventions (NRSI) with varying outcome measures, intervention dosing and comparators. Such data could not be combined with the published RCT results. Accounting for the heterogeneity with a funnel plot and a meta-analysis based on such data would not be a fruitful exercise. Furthermore, standard definitions of disease severity and items of outcomes are not specified globally, and had to be considered as were used by the authors of the respective studies. Non-uniformity of studies in terms of participants, outcomes, confounders etc. affects the external validity in the final analyses. As most of these studies are hospital based, a Berksonian bias is also likely to be present. We did not measure heterogeneity statistically, or the publication bias formally.

## Conclusion

Baricitinib used alone as add on to standard of care or in combination with other treatments has been seen to reduce the risk of mortality and provide significant clinical improvements in hospitalized participants with moderate to severe COVID-19, particularly those requiring high flow oxygen or mechanical ventilation as evidenced from multiple observational and interventional studies published till date. However, the quality of evidence available still needs to be bolstered further to draw definitive conclusions.

## Data Availability

All data produced in the present work are contained in the manuscript

## Declarations

### Ethics approval

Not applicable. *Consent to participate*: Not applicable. *Consent for publication*: Not applicable.

### Availability of data and material

Supplementary data provided.

### Code availability

Not applicable.

### Author’s contributions

SA and AS contributed to the study design and concept. AS and AB carried out the data extraction. SA and RJ reviewed the data. AS and AB prepared the manuscript. SA and RJ edited the manuscript. All authors approved the final manuscript.

## Acknowledgement

None.

---

**Supplementary 1.**
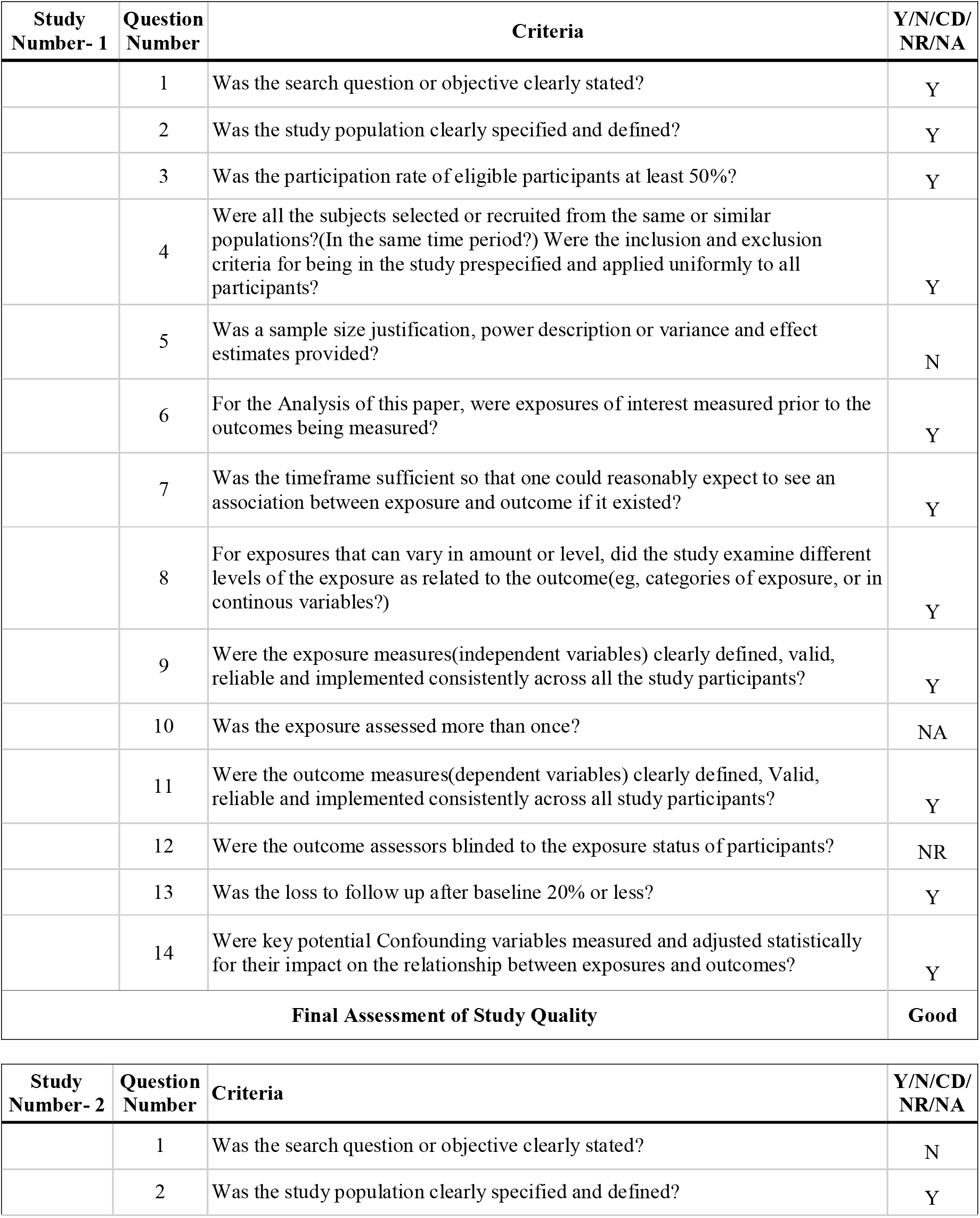
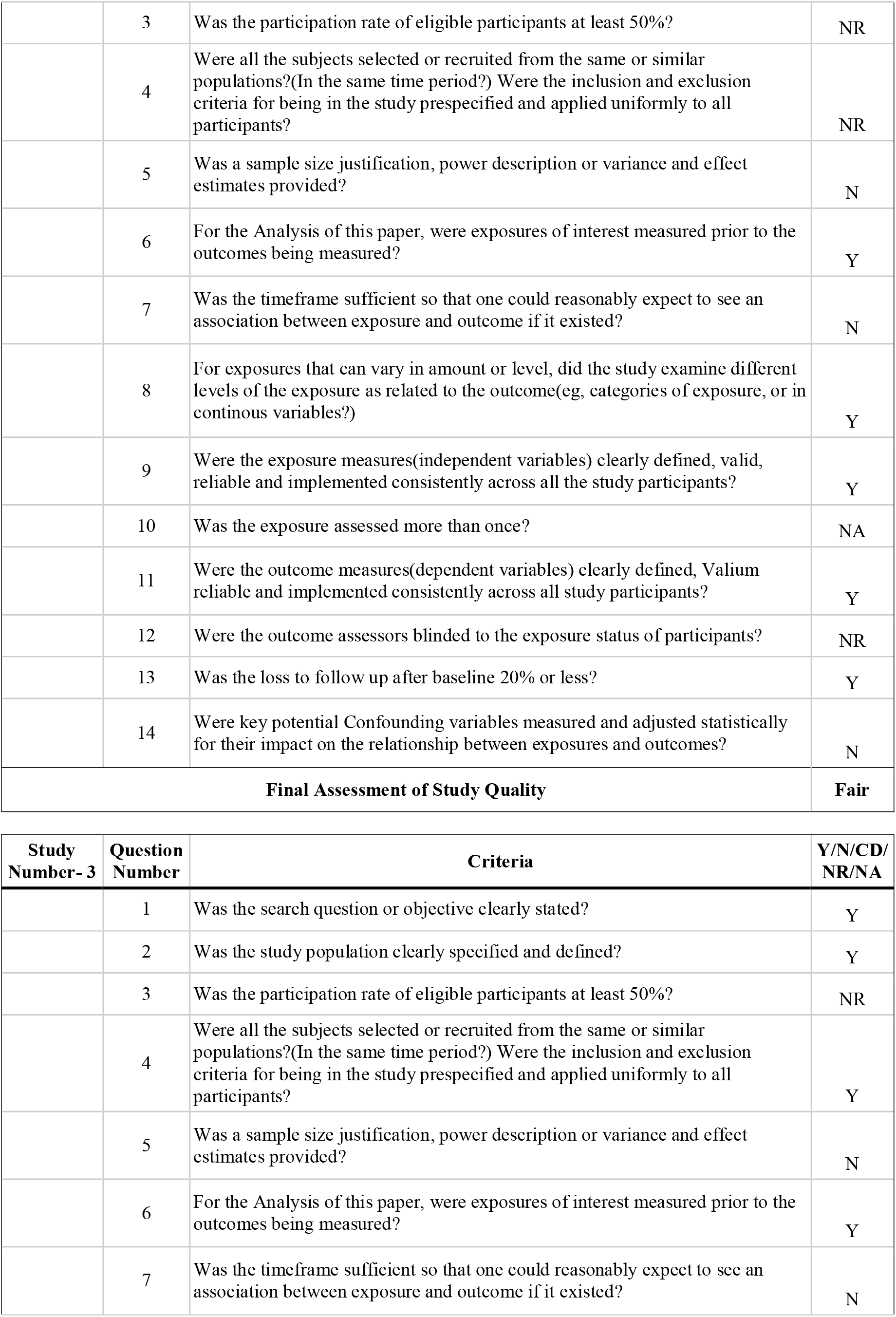
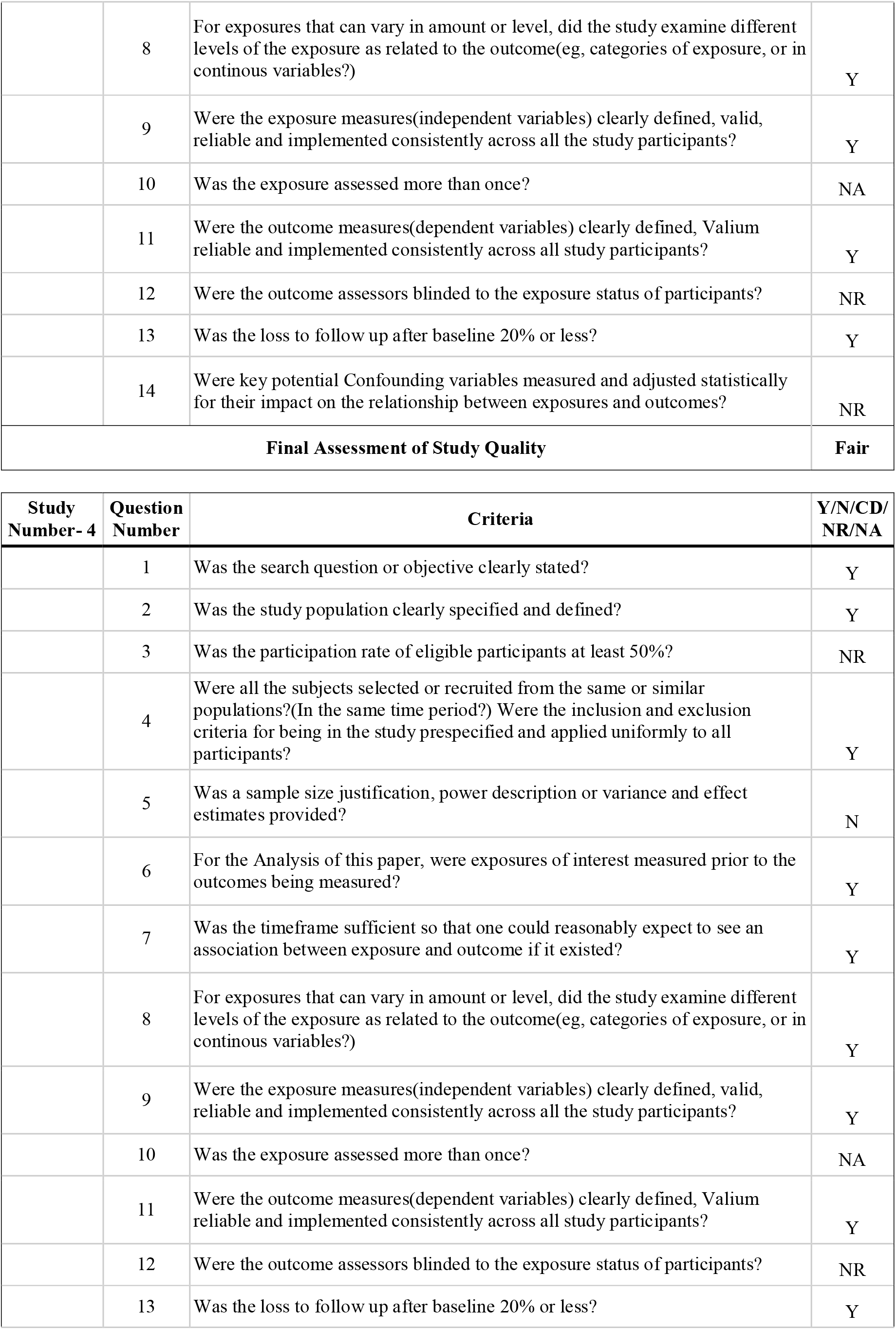
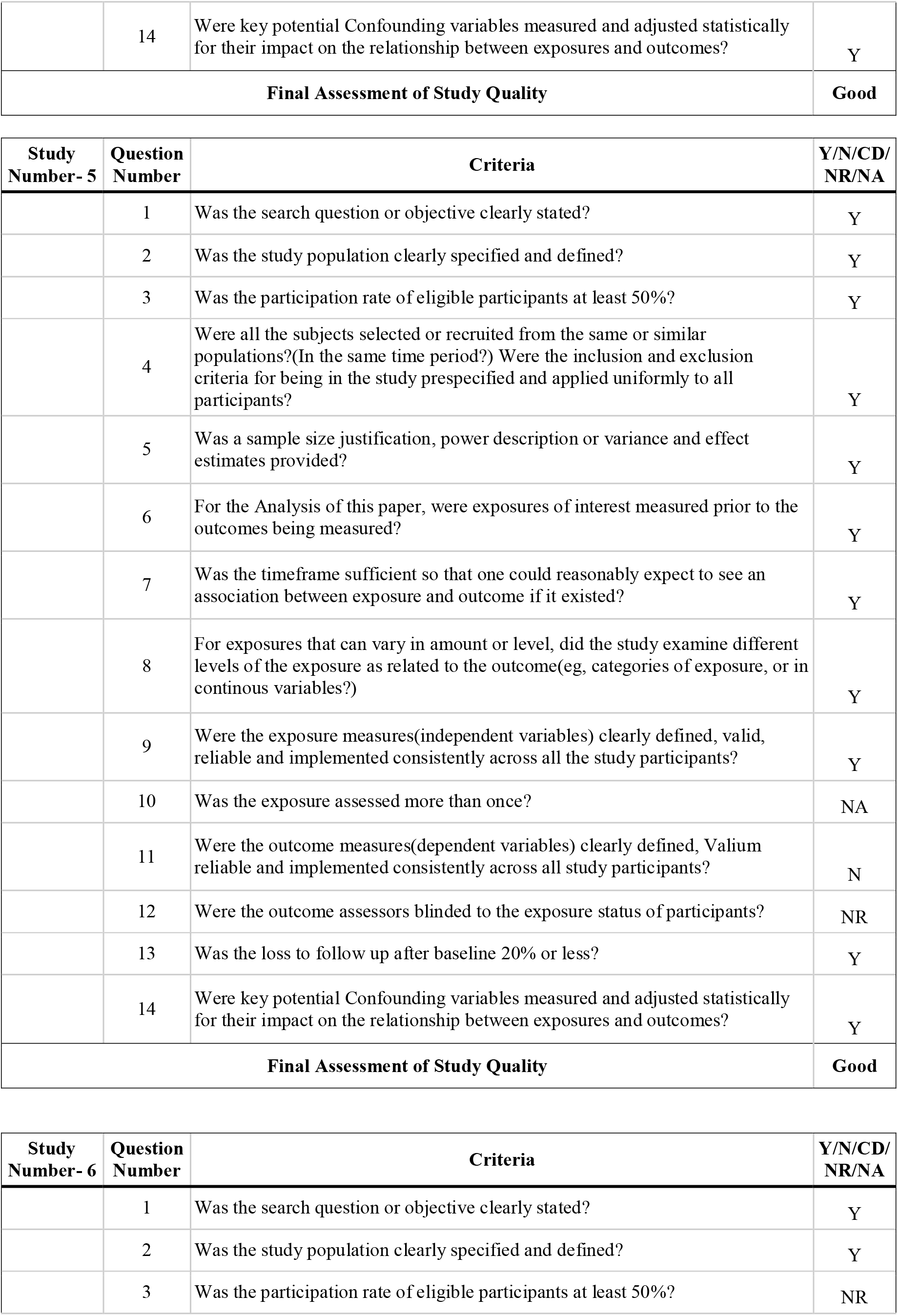
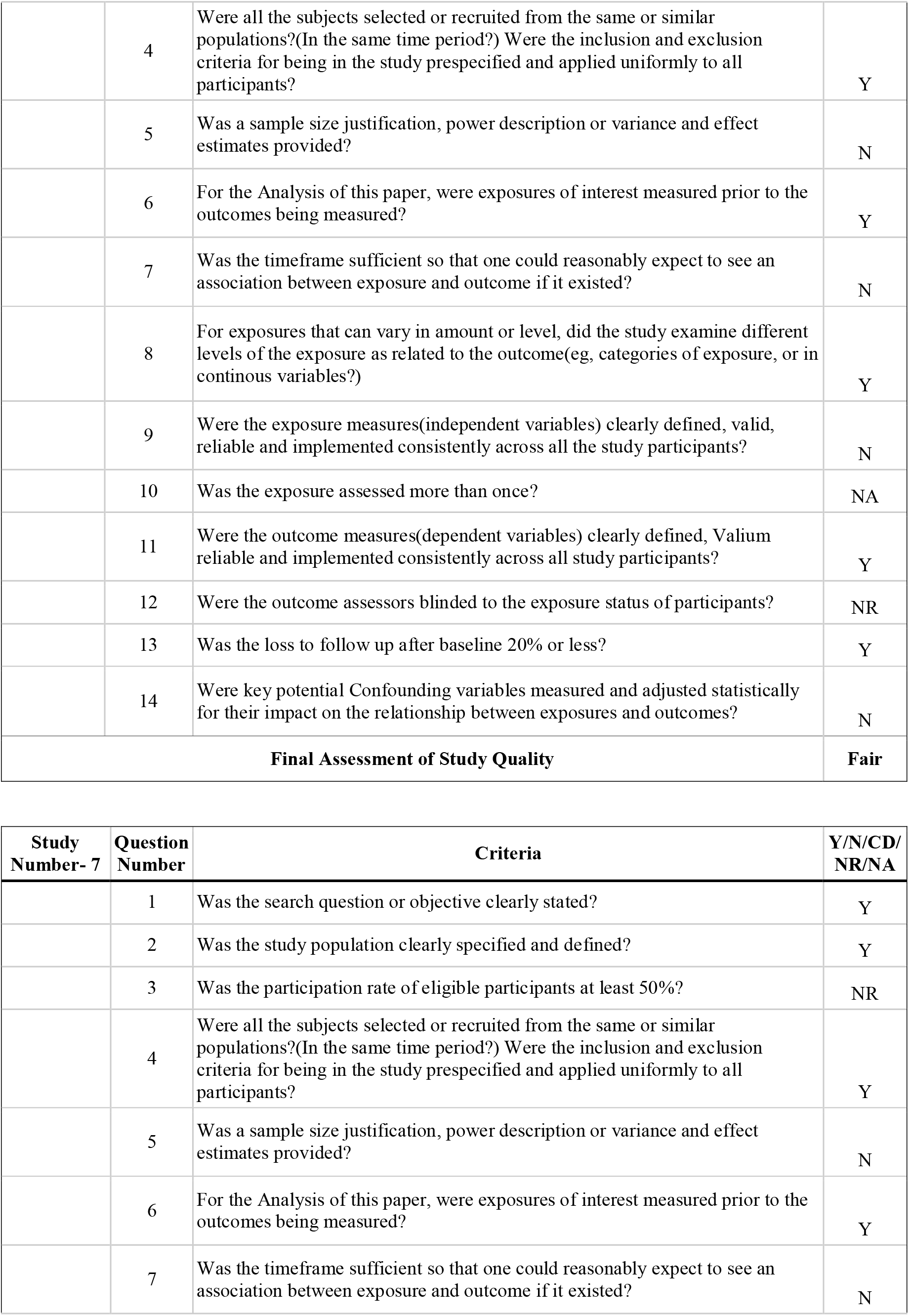
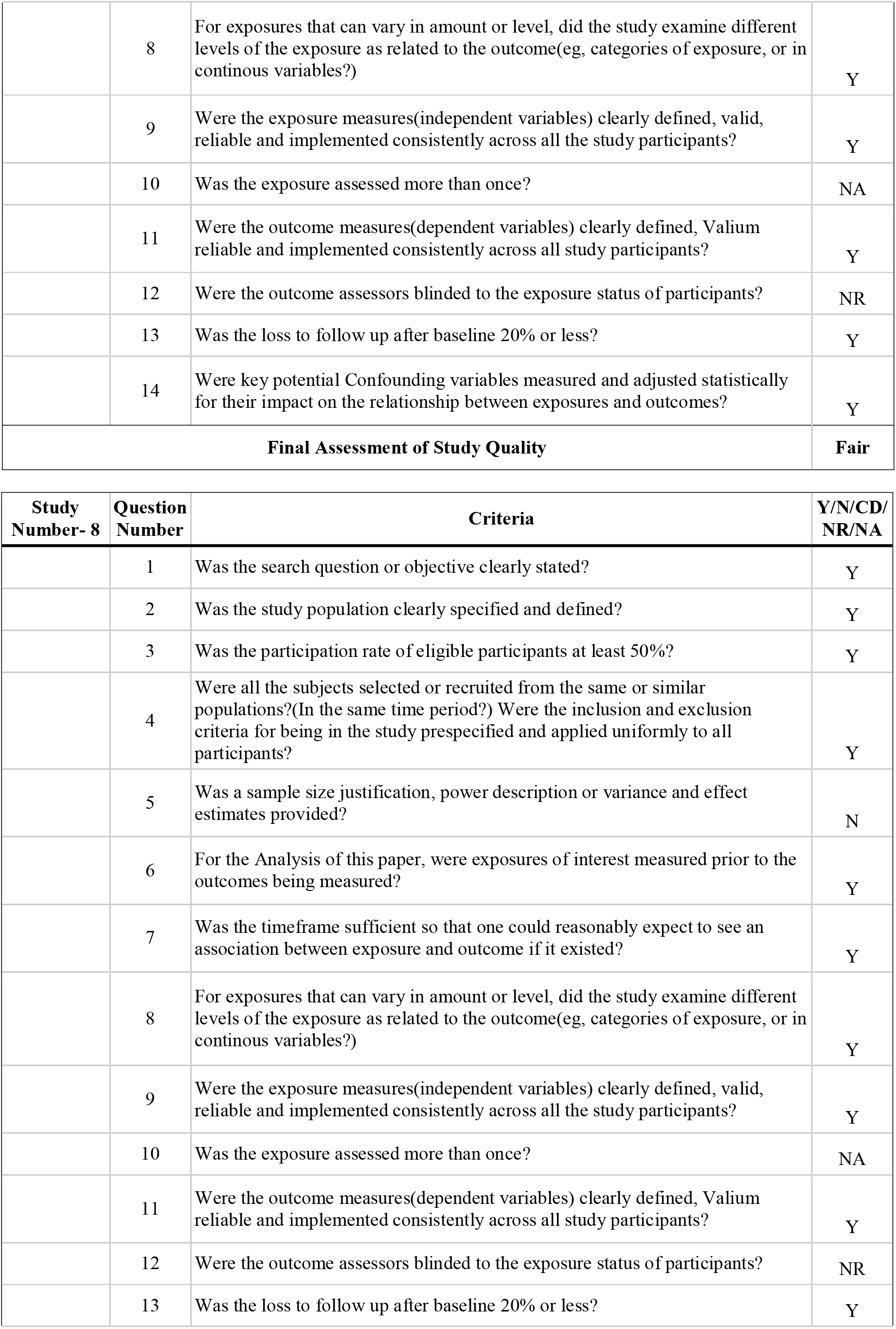
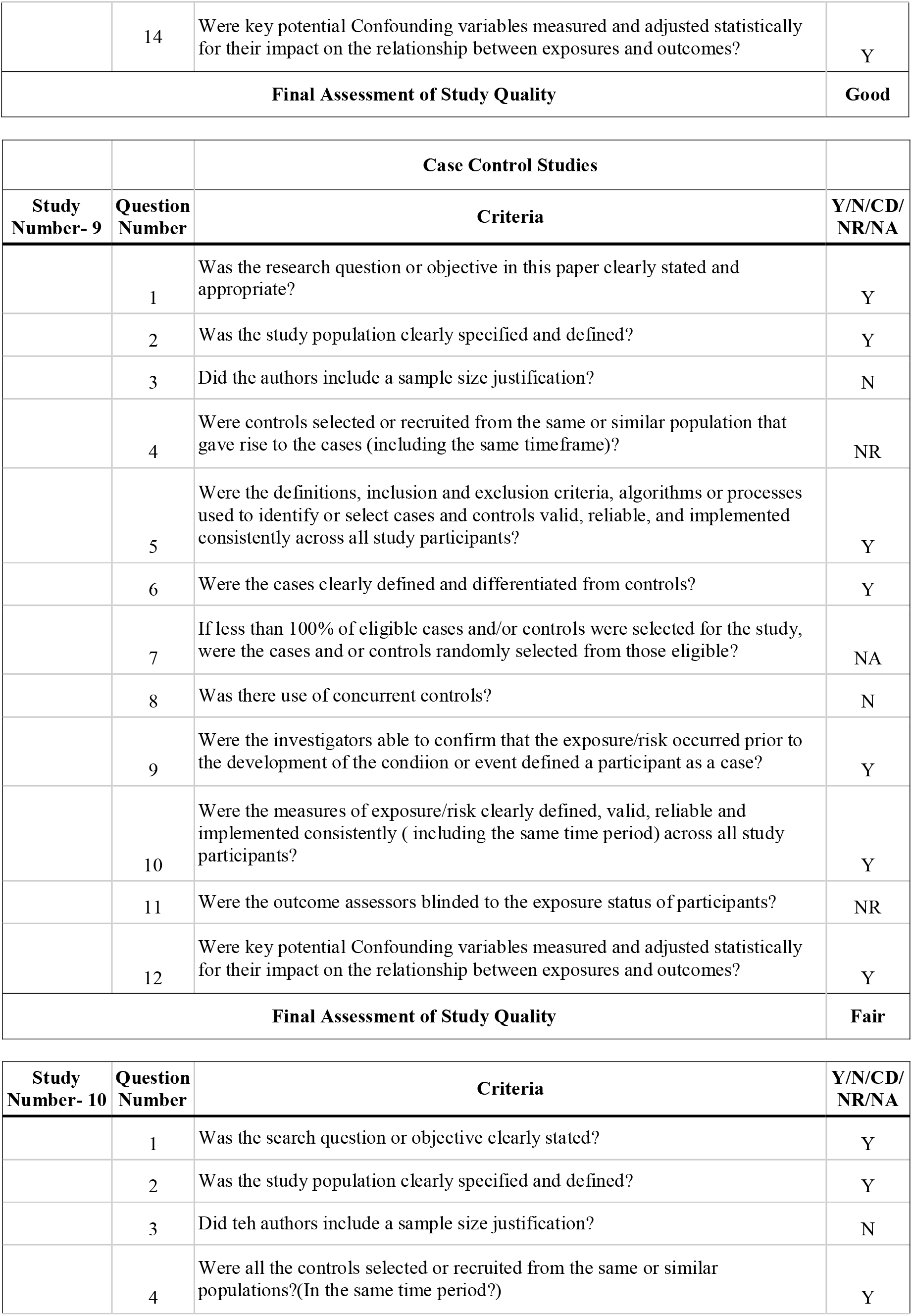
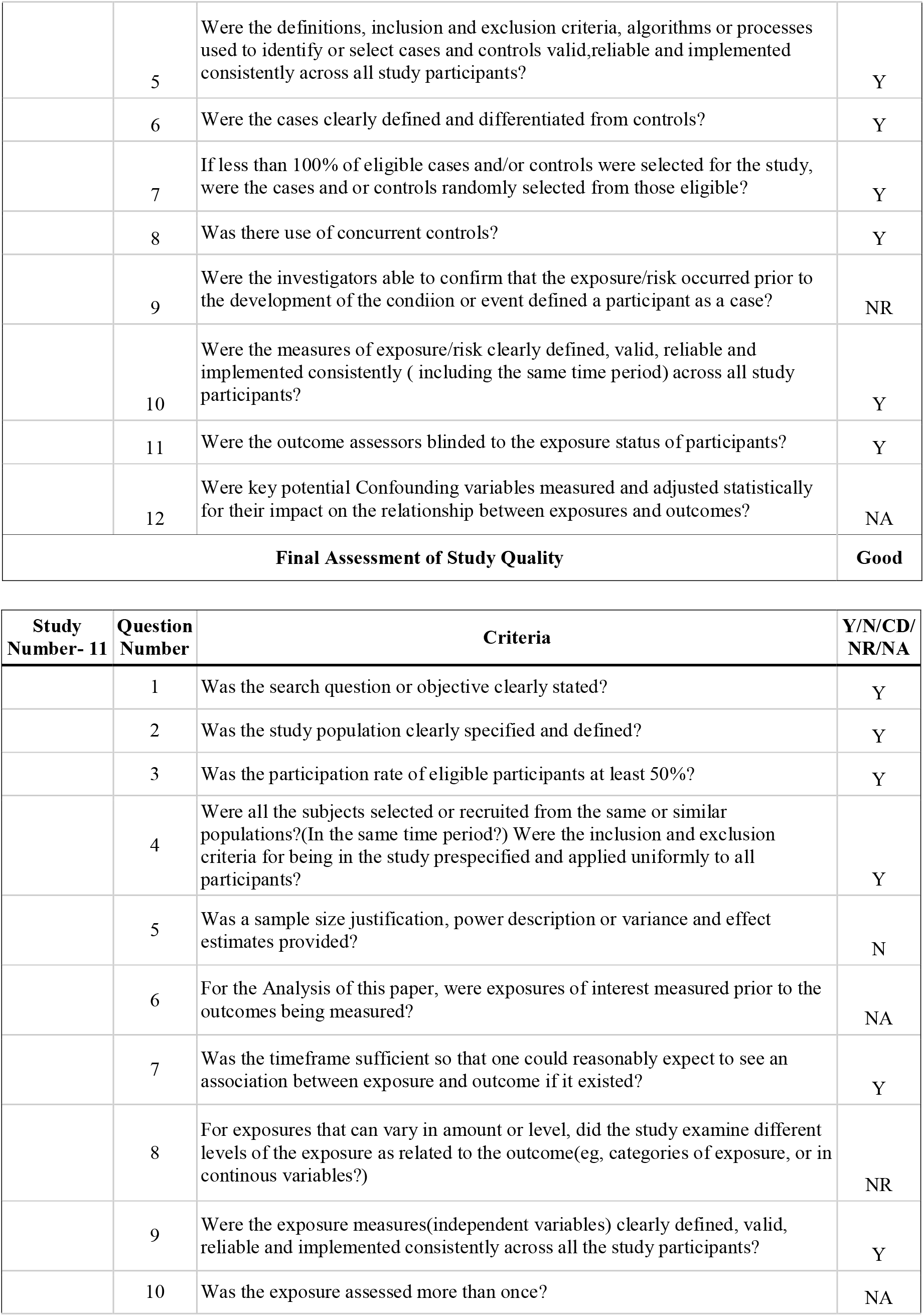
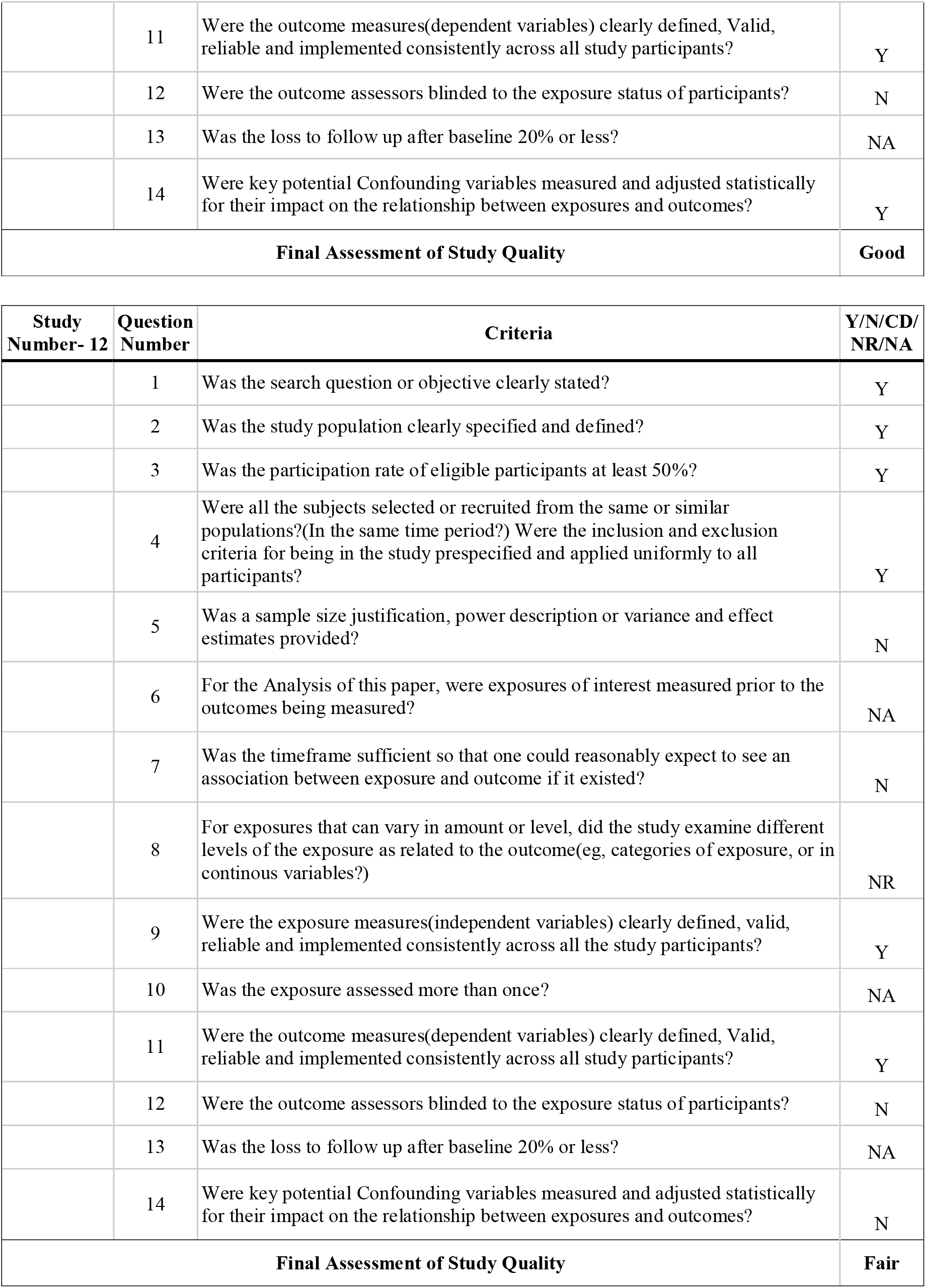
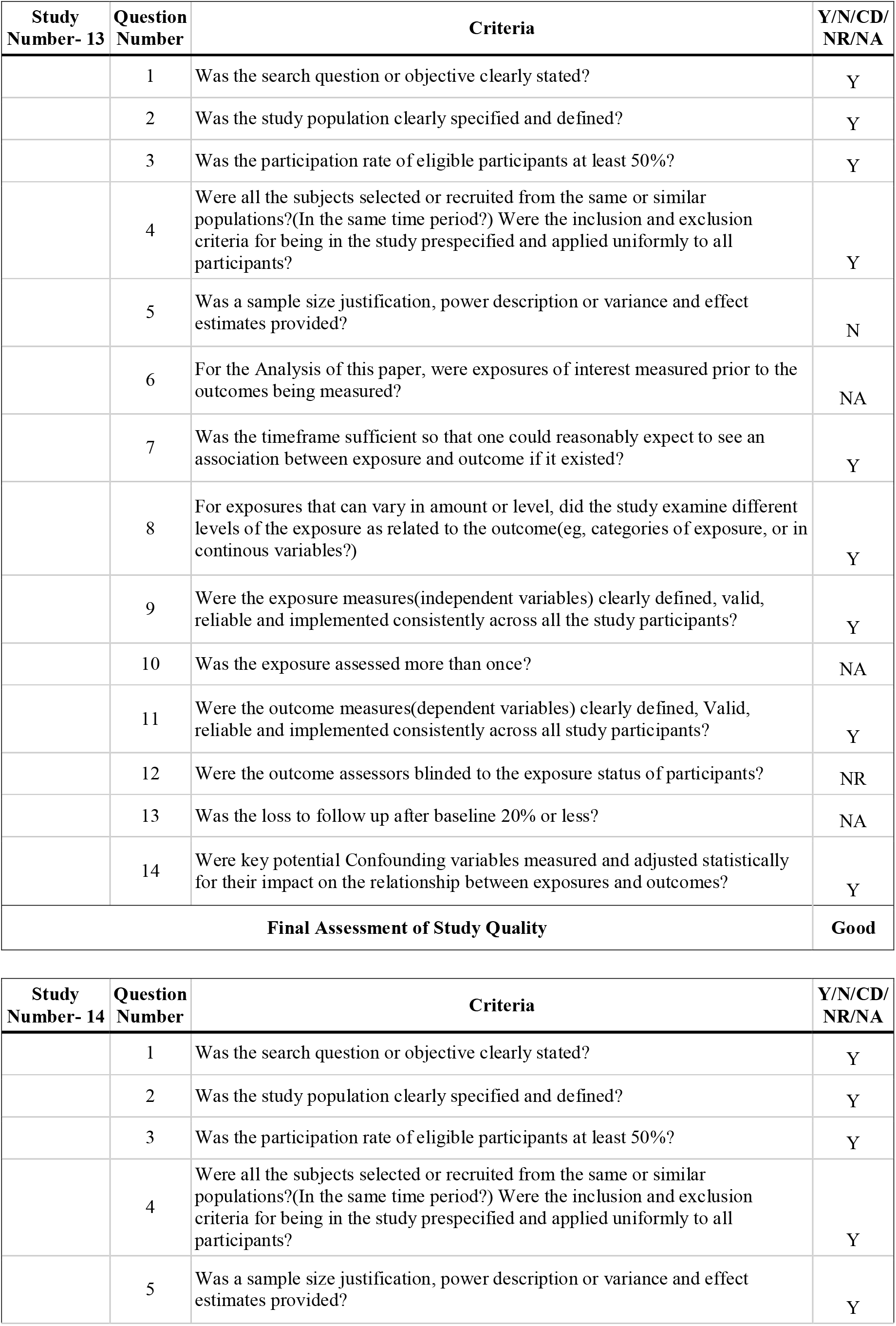
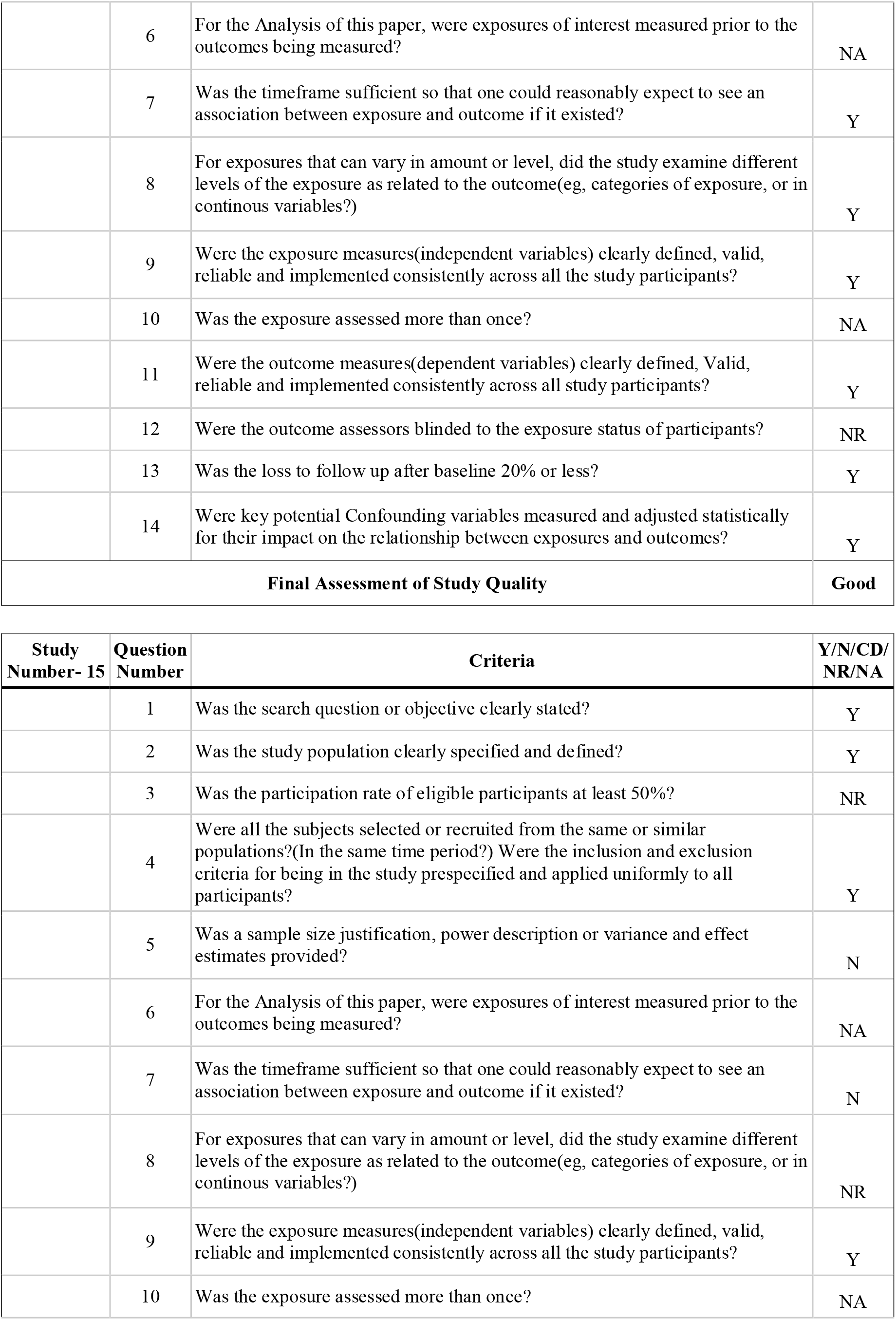
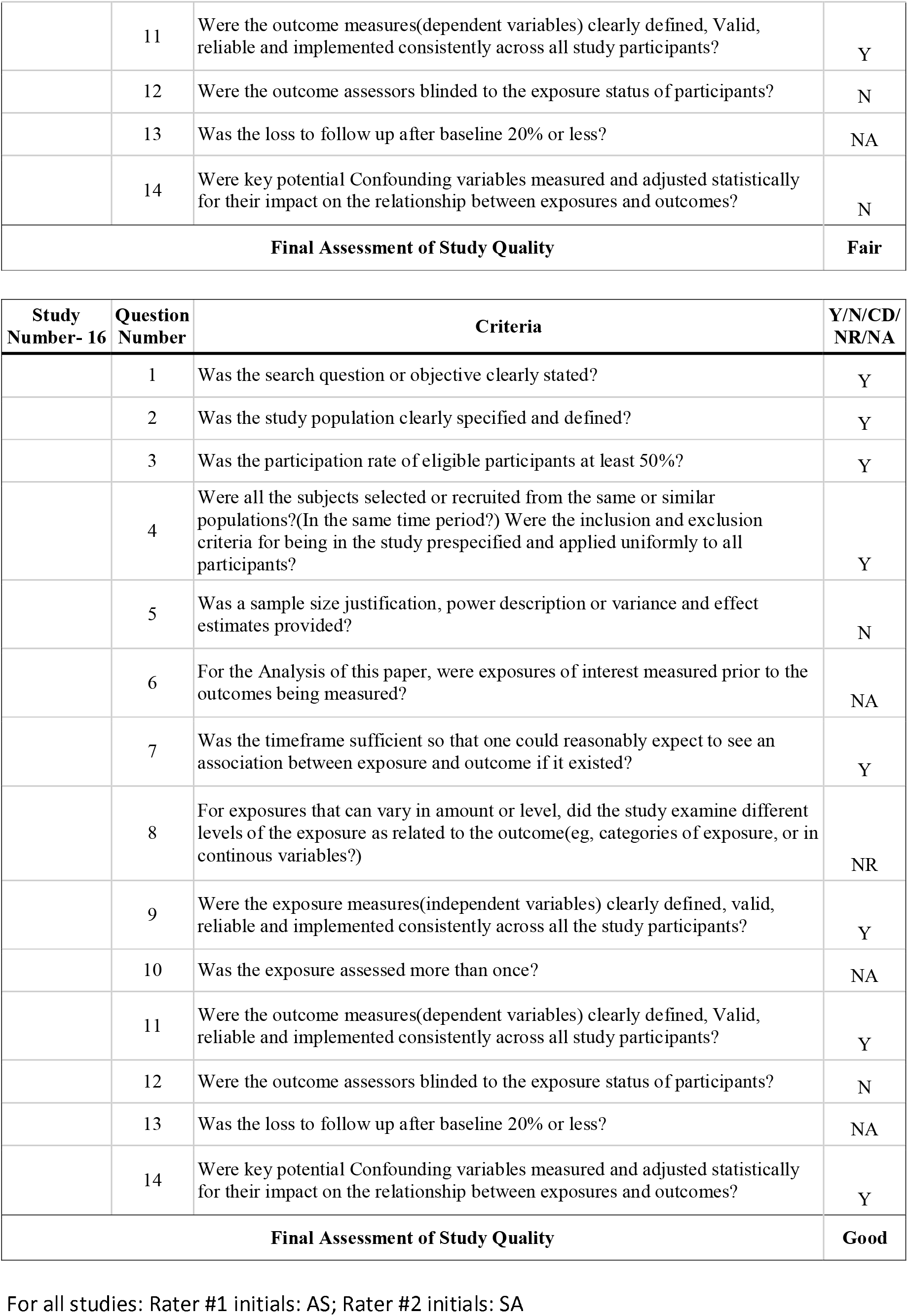
Detailed assessment of the published observational studies using study quality assessment tools observational cohort and cross-sectional studies (To ensure the rubric is more objective, all studies that satisfied by answering more than 70% of the questions with “yes” were graded “good” or low probability of bias”, studies with 45% or above but lesser than 70 were graded as “fair” or “intermediate level of bias” and all others as “low” or high risk of Bias.)

| Study Number- 1 | Question Number | Criteria | Y/N/CD/NR/NA |
| --- | --- | --- | --- |
|  | 1 | Was the search question or objective clearly stated? | Y |
|  | 2 | Was the study population clearly specified and defined? | Y |
|  | 3 | Was the participation rate of eligible participants at least 50%? | Y |
|  | 4 | Were all the subjects selected or recruited from the same or similar populations?(In the same time period?) Were the inclusion and exclusion criteria for being in the study prespecified and applied uniformly to all participants? | Y |
|  | 5 | Was a sample size justification, power description or variance and effect estimates provided? | N |
|  | 6 | For the Analysis of this paper, were exposures of interest measured prior to the outcomes being measured? | Y |
|  | 7 | Was the timeframe sufficient so that one could reasonably expect to see an association between exposure and outcome if it existed? | Y |
|  | 8 | For exposures that can vary in amount or level, did the study examine different levels of the exposure as related to the outcome(eg, categories of exposure, or in continuous variables?) | Y |
|  | 9 | Were the exposure measures(independent variables) clearly defined, valid, reliable and implemented consistently across all the study participants? | Y |
|  | 10 | Was the exposure assessed more than once? | NA |
|  | 11 | Were the outcome measures(dependent variables) clearly defined, Valid, reliable and implemented consistently across all study participants? | Y |
|  | 12 | Were the outcome assessors blinded to the exposure status of participants? | NR |
|  | 13 | Was the loss to follow up after baseline 20% or less? | Y |
|  | 14 | Were key potential Confounding variables measured and adjusted statistically for their impact on the relationship between exposures and outcomes? | Y |
| **Final Assessment of Study Quality** |  |  | **Good** |

| Study Number- 2 | Question Number | Criteria | Y/N/CD/NR/NA |
| --- | --- | --- | --- |
|  | 1 | Was the search question or objective clearly stated? | N |
|  | 2 | Was the study population clearly specified and defined? | Y |
|  | 3 | Was the participation rate of eligible participants at least 50%? | NR |
|  | 4 | Were all the subjects selected or recruited from the same or similar populations?(In the same time period?) Were the inclusion and exclusion criteria for being in the study prespecified and applied uniformly to all participants? | NR |
|  | 5 | Was a sample size justification, power description or variance and effect estimates provided? | N |
|  | 6 | For the Analysis of this paper, were exposures of interest measured prior to the outcomes being measured? | Y |
|  | 7 | Was the timeframe sufficient so that one could reasonably expect to see an association between exposure and outcome if it existed? | N |
|  | 8 | For exposures that can vary in amount or level, did the study examine different levels of the exposure as related to the outcome(eg, categories of exposure, or in continuous variables?) | Y |
|  | 9 | Were the exposure measures(independent variables) clearly defined, valid, reliable and implemented consistently across all the study participants? | Y |
|  | 10 | Was the exposure assessed more than once? | NA |
|  | 11 | Were the outcome measures(dependent variables) clearly defined, Valium reliable and implemented consistently across all study participants? | Y |
|  | 12 | Were the outcome assessors blinded to the exposure status of participants? | NR |
|  | 13 | Was the loss to follow up after baseline 20% or less? | Y |
|  | 14 | Were key potential Confounding variables measured and adjusted statistically for their impact on the relationship between exposures and outcomes? | N |
| **Final Assessment of Study Quality** |  |  | **Fair** |

| **Study Number- 3** | **Question Number** | **Criteria** | **Y/N/CD/NR/NA** |
| --- | --- | --- | --- |
|  | 1 | Was the search question or objective clearly stated? | Y |
|  | 2 | Was the study population clearly specified and defined? | Y |
|  | 3 | Was the participation rate of eligible participants at least 50%? | NR |
|  | 4 | Were all the subjects selected or recruited from the same or similar populations?(In the same time period?) Were the inclusion and exclusion criteria for being in the study prespecified and applied uniformly to all participants? | Y |
|  | 5 | Was a sample size justification, power description or variance and effect estimates provided? | N |
|  | 6 | For the Analysis of this paper, were exposures of interest measured prior to the outcomes being measured? | Y |
|  | 7 | Was the timeframe sufficient so that one could reasonably expect to see an association between exposure and outcome if it existed? | N |
|  | 8 | For exposures that can vary in amount or level, did the study examine different levels of the exposure as related to the outcome(eg, categories of exposure, or in continuous variables?) | Y |
|  | 9 | Were the exposure measures(independent variables) clearly defined, valid, reliable and implemented consistently across all the study participants? | Y |
|  | 10 | Was the exposure assessed more than once? | NA |
|  | 11 | Were the outcome measures(dependent variables) clearly defined, Valium reliable and implemented consistently across all study participants? | Y |
|  | 12 | Were the outcome assessors blinded to the exposure status of participants? | NR |
|  | 13 | Was the loss to follow up after baseline 20% or less? | Y |
|  | 14 | Were key potential Confounding variables measured and adjusted statistically for their impact on the relationship between exposures and outcomes? | NR |
| **Final Assessment of Study Quality** |  |  | **Fair** |

| **Study Number- 4** | **Question Number** | **Criteria** | **Y/N/CD/NR/NA** |
| --- | --- | --- | --- |
|  | 1 | Was the search question or objective clearly stated? | Y |
|  | 2 | Was the study population clearly specified and defined? | Y |
|  | 3 | Was the participation rate of eligible participants at least 50%? | NR |
|  | 4 | Were all the subjects selected or recruited from the same or similar populations?(In the same time period?) Were the inclusion and exclusion criteria for being in the study prespecified and applied uniformly to all participants? | Y |
|  | 5 | Was a sample size justification, power description or variance and effect estimates provided? | N |
|  | 6 | For the Analysis of this paper, were exposures of interest measured prior to the outcomes being measured? | Y |
|  | 7 | Was the timeframe sufficient so that one could reasonably expect to see an association between exposure and outcome if it existed? | Y |
|  | 8 | For exposures that can vary in amount or level, did the study examine different levels of the exposure as related to the outcome(eg, categories of exposure, or in continuous variables?) | Y |
|  | 9 | Were the exposure measures(independent variables) clearly defined, valid, reliable and implemented consistently across all the study participants? | Y |
|  | 10 | Was the exposure assessed more than once? | NA |
|  | 11 | Were the outcome measures(dependent variables) clearly defined, Valium reliable and implemented consistently across all study participants? | Y |
|  | 12 | Were the outcome assessors blinded to the exposure status of participants? | NR |
|  | 13 | Was the loss to follow up after baseline 20% or less? | Y |
|  | 14 | Were key potential Confounding variables measured and adjusted statistically for their impact on the relationship between exposures and outcomes? | Y |
| **Final Assessment of Study Quality** |  |  | **Good** |

| **Study Number- 5** | **Question Number** | **Criteria** | **Y/N/CD/NR/NA** |
| --- | --- | --- | --- |
|  | 1 | Was the search question or objective clearly stated? | Y |
|  | 2 | Was the study population clearly specified and defined? | Y |
|  | 3 | Was the participation rate of eligible participants at least 50%? | Y |
|  | 4 | Were all the subjects selected or recruited from the same or similar populations?(In the same time period?) Were the inclusion and exclusion criteria for being in the study prespecified and applied uniformly to all participants? | Y |
|  | 5 | Was a sample size justification, power description or variance and effect estimates provided? | Y |
|  | 6 | For the Analysis of this paper, were exposures of interest measured prior to the outcomes being measured? | Y |
|  | 7 | Was the timeframe sufficient so that one could reasonably expect to see an association between exposure and outcome if it existed? | Y |
|  | 8 | For exposures that can vary in amount or level, did the study examine different levels of the exposure as related to the outcome(eg, categories of exposure, or in continuous variables?) | Y |
|  | 9 | Were the exposure measures(independent variables) clearly defined, valid, reliable and implemented consistently across all the study participants? | Y |
|  | 10 | Was the exposure assessed more than once? | NA |
|  | 11 | Were the outcome measures(dependent variables) clearly defined, Valid, reliable and implemented consistently across all study participants? | N |
|  | 12 | Were the outcome assessors blinded to the exposure status of participants? | NR |
|  | 13 | Was the loss to follow up after baseline 20% or less? | Y |
|  | 14 | Were key potential Confounding variables measured and adjusted statistically for their impact on the relationship between exposures and outcomes? | Y |
| **Final Assessment of Study Quality** |  |  | **Good** |

| **Study Number- 6** | **Question Number** | **Criteria** | **Y/N/CD/NR/NA** |
| --- | --- | --- | --- |
|  | 1 | Was the search question or objective clearly stated? | Y |
|  | 2 | Was the study population clearly specified and defined? | Y |
|  | 3 | Was the participation rate of eligible participants at least 50%? | NR |
|  | 4 | Were all the subjects selected or recruited from the same or similar populations?(In the same time period?) Were the inclusion and exclusion criteria for being in the study prespecified and applied uniformly to all participants? | Y |
|  | 5 | Was a sample size justification, power description or variance and effect estimates provided? | N |
|  | 6 | For the Analysis of this paper, were exposures of interest measured prior to the outcomes being measured? | Y |
|  | 7 | Was the timeframe sufficient so that one could reasonably expect to see an association between exposure and outcome if it existed? | N |
|  | 8 | For exposures that can vary in amount or level, did the study examine different levels of the exposure as related to the outcome(eg, categories of exposure, or in continuous variables?) | Y |
|  | 9 | Were the exposure measures(independent variables) clearly defined, valid, reliable and implemented consistently across all the study participants? | N |
|  | 10 | Was the exposure assessed more than once? | NA |
|  | 11 | Were the outcome measures(dependent variables) clearly defined, Valid, reliable and implemented consistently across all study participants? | Y |
|  | 12 | Were the outcome assessors blinded to the exposure status of participants? | NR |
|  | 13 | Was the loss to follow up after baseline 20% or less? | Y |
|  | 14 | Were key potential Confounding variables measured and adjusted statistically for their impact on the relationship between exposures and outcomes? | N |
| **Final Assessment of Study Quality** |  |  | **Fair** |

| **Study Number- 7** | **Question Number** | **Criteria** | **Y/N/CD/NR/NA** |
| --- | --- | --- | --- |
|  | 1 | Was the search question or objective clearly stated? | Y |
|  | 2 | Was the study population clearly specified and defined? | Y |
|  | 3 | Was the participation rate of eligible participants at least 50% ? | NR |
|  | 4 | Were all the subjects selected or recruited from the same or similar populations?(In the same time period?) Were the inclusion and exclusion criteria for being in the study prespecified and applied uniformly to all participants? | Y |
|  | 5 | Was a sample size justification, power description or variance and effect estimates provided? | N |
|  | 6 | For the Analysis of this paper, were exposures of interest measured prior to the outcomes being measured? | Y |
|  | 7 | Was the timeframe sufficient so that one could reasonably expect to see an association between exposure and outcome if it existed? | N |
|  | 8 | For exposures that can vary in amount or level, did the study examine different levels of the exposure as related to the outcome(eg, categories of exposure, or in continuous variables?) | Y |
|  | 9 | Were the exposure measures(independent variables) clearly defined, valid, reliable and implemented consistently across all the study participants? | Y |
|  | 10 | Was the exposure assessed more than once? | NA |
|  | 11 | Were the outcome measures(dependent variables) clearly defined, Valium reliable and implemented consistently across all study participants? | Y |
|  | 12 | Were the outcome assessors blinded to the exposure status of participants? | NR |
|  | 13 | Was the loss to follow up after baseline 20% or less? | Y |
|  | 14 | Were key potential Confounding variables measured and adjusted statistically for their impact on the relationship between exposures and outcomes? | Y |
| **Final Assessment of Study Quality** |  |  | **Fair** |

| **Study Number- 8** | **Question Number** | **Criteria** | **Y/N/CD/NR/NA** |
| --- | --- | --- | --- |
|  | 1 | Was the search question or objective clearly stated? | Y |
|  | 2 | Was the study population clearly specified and defined? | Y |
|  | 3 | Was the participation rate of eligible participants at least 50% ? | Y |
|  | 4 | Were all the subjects selected or recruited from the same or similar populations?(In the same time period?) Were the inclusion and exclusion criteria for being in the study prespecified and applied uniformly to all participants? | Y |
|  | 5 | Was a sample size justification, power description or variance and effect estimates provided? | N |
|  | 6 | For the Analysis of this paper, were exposures of interest measured prior to the outcomes being measured? | Y |
|  | 7 | Was the timeframe sufficient so that one could reasonably expect to see an association between exposure and outcome if it existed? | Y |
|  | 8 | For exposures that can vary in amount or level, did the study examine different levels of the exposure as related to the outcome(eg, categories of exposure, or in continuous variables?) | Y |
|  | 9 | Were the exposure measures(independent variables) clearly defined, valid, reliable and implemented consistently across all the study participants? | Y |
|  | 10 | Was the exposure assessed more than once? | NA |
|  | 11 | Were the outcome measures(dependent variables) clearly defined, Valium reliable and implemented consistently across all study participants? | Y |
|  | 12 | Were the outcome assessors blinded to the exposure status of participants? | NR |
|  | 13 | Was the loss to follow up after baseline 20% or less? | Y |
|  | 14 | Were key potential Confounding variables measured and adjusted statistically for their impact on the relationship between exposures and outcomes? | Y |
| **Final Assessment of Study Quality** |  |  | **Good** |

|  |  | **Case Control Studies** |  |
| --- | --- | --- | --- |
| **Study Number- 9** | **Question Number** | **Criteria** | **Y/N/CD/NR/NA** |
|  | 1 | Was the research question or objective in this paper clearly stated and appropriate? | Y |
|  | 2 | Was the study population clearly specified and defined? | Y |
|  | 3 | Did the authors include a sample size justification? | N |
|  | 4 | Were controls selected or recruited from the same or similar population that gave rise to the cases (including the same timeframe)? | NR |
|  | 5 | Were the definitions, inclusion and exclusion criteria, algorithms or processes used to identify or select cases and controls valid, reliable, and implemented consistently across all study participants? | Y |
|  | 6 | Were the cases clearly defined and differentiated from controls? | Y |
|  | 7 | If less than 100% of eligible cases and/or controls were selected for the study, were the cases and or controls randomly selected from those eligible? | NA |
|  | 8 | Was there use of concurrent controls? | N |
|  | 9 | Were the investigators able to confirm that the exposure/risk occurred prior to the development of the condition or event defined a participant as a case? | Y |
|  | 10 | Were the measures of exposure/risk clearly defined, valid, reliable and implemented consistently ( including the same time period) across all study participants? | Y |
|  | 11 | Were the outcome assessors blinded to the exposure status of participants? | NR |
|  | 12 | Were key potential Confounding variables measured and adjusted statistically for their impact on the relationship between exposures and outcomes? | Y |
| **Final Assessment of Study Quality** |  |  | **Fair** |

| **Study Number- 10** | **Question Number** | **Criteria** | **Y/N/CD/NR/NA** |
| --- | --- | --- | --- |
|  | 1 | Was the search question or objective clearly stated? | Y |
|  | 2 | Was the study population clearly specified and defined? | Y |
|  | 3 | Did the authors include a sample size justification? | N |
|  | 4 | Were all the controls selected or recruited from the same or similar populations?(In the same time period?) | Y |
|  | 5 | Were the definitions, inclusion and exclusion criteria, algorithms or processes used to identify or select cases and controls valid, reliable and implemented consistently across all study participants? | Y |
|  | 6 | Were the cases clearly defined and differentiated from controls? | Y |
|  | 7 | If less than 100% of eligible cases and/or controls were selected for the study, were the cases and or controls randomly selected from those eligible? | Y |
|  | 8 | Was there use of concurrent controls? | Y |
|  | 9 | Were the investigators able to confirm that the exposure/risk occurred prior to the development of the condition or event defined a participant as a case? | NR |
|  | 10 | Were the measures of exposure/risk clearly defined, valid, reliable and implemented consistently (including the same time period) across all study participants? | Y |
|  | 11 | Were the outcome assessors blinded to the exposure status of participants? | Y |
|  | 12 | Were key potential Confounding variables measured and adjusted statistically for their impact on the relationship between exposures and outcomes? | NA |
| **Final Assessment of Study Quality** |  |  | **Good** |

| **Study Number- 11** | **Question Number** | **Criteria** | **Y/N/CD/NR/NA** |
| --- | --- | --- | --- |
|  | 1 | Was the search question or objective clearly stated? | Y |
|  | 2 | Was the study population clearly specified and defined? | Y |
|  | 3 | Was the participation rate of eligible participants at least 50%? | Y |
|  | 4 | Were all the subjects selected or recruited from the same or similar populations?(In the same time period?) Were the inclusion and exclusion criteria for being in the study prespecified and applied uniformly to all participants? | Y |
|  | 5 | Was a sample size justification, power description or variance and effect estimates provided? | N |
|  | 6 | For the Analysis of this paper, were exposures of interest measured prior to the outcomes being measured? | NA |
|  | 7 | Was the timeframe sufficient so that one could reasonably expect to see an association between exposure and outcome if it existed? | Y |
|  | 8 | For exposures that can vary in amount or level, did the study examine different levels of the exposure as related to the outcome(eg, categories of exposure, or in continuous variables?) | NR |
|  | 9 | Were the exposure measures(independent variables) clearly defined, valid, reliable and implemented consistently across all the study participants? | Y |
|  | 10 | Was the exposure assessed more than once? | NA |
|  | 11 | Were the outcome measures(dependent variables) clearly defined, Valid, reliable and implemented consistently across all study participants? | Y |
|  | 12 | Were the outcome assessors blinded to the exposure status of participants? | N |
|  | 13 | Was the loss to follow up after baseline 20% or less? | NA |
|  | 14 | Were key potential Confounding variables measured and adjusted statistically for their impact on the relationship between exposures and outcomes? | Y |
| **Final Assessment of Study Quality** |  |  | **Good** |

| **Study Number- 12** | **Question Number** | **Criteria** | **Y/N/CD/NR/NA** |
| --- | --- | --- | --- |
|  | 1 | Was the search question or objective clearly stated? | Y |
|  | 2 | Was the study population clearly specified and defined? | Y |
|  | 3 | Was the participation rate of eligible participants at least 50%? | Y |
|  | 4 | Were all the subjects selected or recruited from the same or similar populations?(In the same time period?) Were the inclusion and exclusion criteria for being in the study prespecified and applied uniformly to all participants? | Y |
|  | 5 | Was a sample size justification, power description or variance and effect estimates provided? | N |
|  | 6 | For the Analysis of this paper, were exposures of interest measured prior to the outcomes being measured? | NA |
|  | 7 | Was the timeframe sufficient so that one could reasonably expect to see an association between exposure and outcome if it existed? | N |
|  | 8 | For exposures that can vary in amount or level, did the study examine different levels of the exposure as related to the outcome(eg, categories of exposure, or in continuous variables?) | NR |
|  | 9 | Were the exposure measures(independent variables) clearly defined, valid, reliable and implemented consistently across all the study participants? | Y |
|  | 10 | Was the exposure assessed more than once? | NA |
|  | 11 | Were the outcome measures(dependent variables) clearly defined, Valid, reliable and implemented consistently across all study participants? | Y |
|  | 12 | Were the outcome assessors blinded to the exposure status of participants? | N |
|  | 13 | Was the loss to follow up after baseline 20% or less? | NA |
|  | 14 | Were key potential Confounding variables measured and adjusted statistically for their impact on the relationship between exposures and outcomes? | N |
| **Final Assessment of Study Quality** |  |  | **Fair** |

| Study Number- 13 | Question Number | Criteria | Y/N/CD/NR/NA |
| --- | --- | --- | --- |
|  | 1 | Was the search question or objective clearly stated? | Y |
|  | 2 | Was the study population clearly specified and defined? | Y |
|  | 3 | Was the participation rate of eligible participants at least 50%? | Y |
|  | 4 | Were all the subjects selected or recruited from the same or similar populations?(In the same time period?) Were the inclusion and exclusion criteria for being in the study prespecified and applied uniformly to all participants? | Y |
|  | 5 | Was a sample size justification, power description or variance and effect estimates provided? | N |
|  | 6 | For the Analysis of this paper, were exposures of interest measured prior to the outcomes being measured? | NA |
|  | 7 | Was the timeframe sufficient so that one could reasonably expect to see an association between exposure and outcome if it existed? | Y |
|  | 8 | For exposures that can vary in amount or level, did the study examine different levels of the exposure as related to the outcome(eg, categories of exposure, or in continuous variables?) | Y |
|  | 9 | Were the exposure measures(independent variables) clearly defined, valid, reliable and implemented consistently across all the study participants? | Y |
|  | 10 | Was the exposure assessed more than once? | NA |
|  | 11 | Were the outcome measures(dependent variables) clearly defined, Valid, reliable and implemented consistently across all study participants? | Y |
|  | 12 | Were the outcome assessors blinded to the exposure status of participants? | NR |
|  | 13 | Was the loss to follow up after baseline 20% or less? | NA |
|  | 14 | Were key potential Confounding variables measured and adjusted statistically for their impact on the relationship between exposures and outcomes? | Y |
| **Final Assessment of Study Quality** |  |  | **Good** |

| Study Number- 14 | Question Number | Criteria | Y/N/CD/NR/NA |
| --- | --- | --- | --- |
|  | 1 | Was the search question or objective clearly stated? | Y |
|  | 2 | Was the study population clearly specified and defined? | Y |
|  | 3 | Was the participation rate of eligible participants at least 50%? | Y |
|  | 4 | Were all the subjects selected or recruited from the same or similar populations?(In the same time period?) Were the inclusion and exclusion criteria for being in the study prespecified and applied uniformly to all participants? | Y |
|  | 5 | Was a sample size justification, power description or variance and effect estimates provided? | Y |
|  | 6 | For the Analysis of this paper, were exposures of interest measured prior to the outcomes being measured? | NA |
|  | 7 | Was the timeframe sufficient so that one could reasonably expect to see an association between exposure and outcome if it existed? | Y |
|  | 8 | For exposures that can vary in amount or level, did the study examine different levels of the exposure as related to the outcome(eg, categories of exposure, or in continuous variables?) | Y |
|  | 9 | Were the exposure measures(independent variables) clearly defined, valid, reliable and implemented consistently across all the study participants? | Y |
|  | 10 | Was the exposure assessed more than once? | NA |
|  | 11 | Were the outcome measures(dependent variables) clearly defined, Valid, reliable and implemented consistently across all study participants? | Y |
|  | 12 | Were the outcome assessors blinded to the exposure status of participants? | NR |
|  | 13 | Was the loss to follow up after baseline 20% or less? | Y |
|  | 14 | Were key potential Confounding variables measured and adjusted statistically for their impact on the relationship between exposures and outcomes? | Y |
| **Final Assessment of Study Quality** |  |  | **Good** |

| **Study Number- 15** | **Question Number** | **Criteria** | **Y/N/CD/NR/NA** |
| --- | --- | --- | --- |
|  | 1 | Was the search question or objective clearly stated? | Y |
|  | 2 | Was the study population clearly specified and defined? | Y |
|  | 3 | Was the participation rate of eligible participants at least 50%? | NR |
|  | 4 | Were all the subjects selected or recruited from the same or similar populations?(In the same time period?) Were the inclusion and exclusion criteria for being in the study prespecified and applied uniformly to all participants? | Y |
|  | 5 | Was a sample size justification, power description or variance and effect estimates provided? | N |
|  | 6 | For the Analysis of this paper, were exposures of interest measured prior to the outcomes being measured? | NA |
|  | 7 | Was the timeframe sufficient so that one could reasonably expect to see an association between exposure and outcome if it existed? | N |
|  | 8 | For exposures that can vary in amount or level, did the study examine different levels of the exposure as related to the outcome(eg, categories of exposure, or in continuous variables?) | NR |
|  | 9 | Were the exposure measures(independent variables) clearly defined, valid, reliable and implemented consistently across all the study participants? | Y |
|  | 10 | Was the exposure assessed more than once? | NA |
|  | 11 | Were the outcome measures(dependent variables) clearly defined, Valid, reliable and implemented consistently across all study participants? | Y |
|  | 12 | Were the outcome assessors blinded to the exposure status of participants? | N |
|  | 13 | Was the loss to follow up after baseline 20% or less? | NA |
|  | 14 | Were key potential Confounding variables measured and adjusted statistically for their impact on the relationship between exposures and outcomes? | N |
| **Final Assessment of Study Quality** |  |  | **Fair** |

| **Study Number- 16** | **Question Number** | **Criteria** | **Y/N/CD/NR/NA** |
| --- | --- | --- | --- |
|  | 1 | Was the search question or objective clearly stated? | Y |
|  | 2 | Was the study population clearly specified and defined? | Y |
|  | 3 | Was the participation rate of eligible participants at least 50%? | Y |
|  | 4 | Were all the subjects selected or recruited from the same or similar populations?(In the same time period?) Were the inclusion and exclusion criteria for being in the study prespecified and applied uniformly to all participants? | Y |
|  | 5 | Was a sample size justification, power description or variance and effect estimates provided? | N |
|  | 6 | For the Analysis of this paper, were exposures of interest measured prior to the outcomes being measured? | NA |
|  | 7 | Was the timeframe sufficient so that one could reasonably expect to see an association between exposure and outcome if it existed? | Y |
|  | 8 | For exposures that can vary in amount or level, did the study examine different levels of the exposure as related to the outcome(eg, categories of exposure, or in continuous variables?) | NR |
|  | 9 | Were the exposure measures(independent variables) clearly defined, valid, reliable and implemented consistently across all the study participants? | Y |
|  | 10 | Was the exposure assessed more than once? | NA |
|  | 11 | Were the outcome measures(dependent variables) clearly defined, Valid, reliable and implemented consistently across all study participants? | Y |
|  | 12 | Were the outcome assessors blinded to the exposure status of participants? | N |
|  | 13 | Was the loss to follow up after baseline 20% or less? | NA |
|  | 14 | Were key potential Confounding variables measured and adjusted statistically for their impact on the relationship between exposures and outcomes? | Y |
| **Final Assessment of Study Quality** |  |  | **Good** |

**Supplementary 2.**
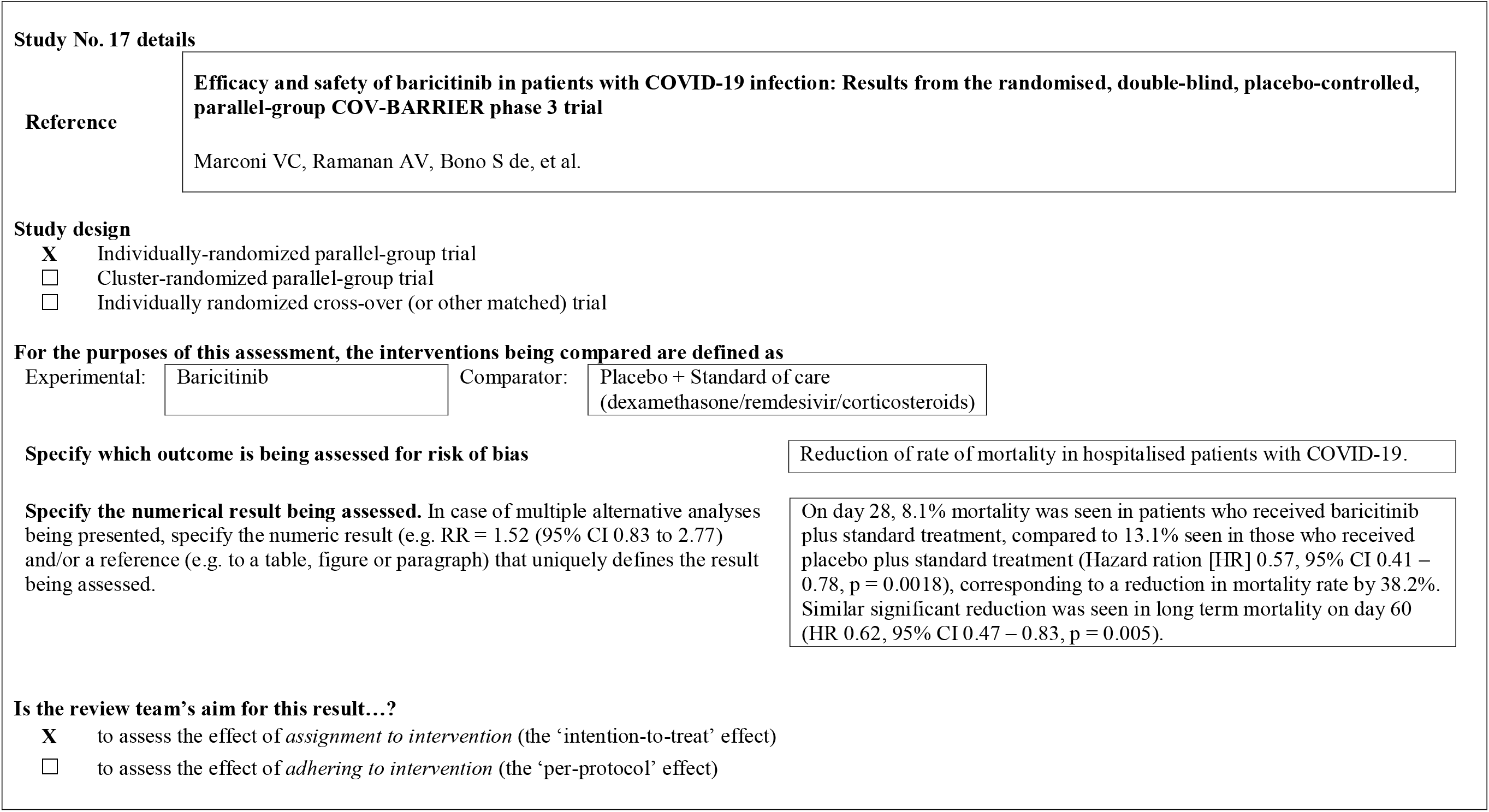
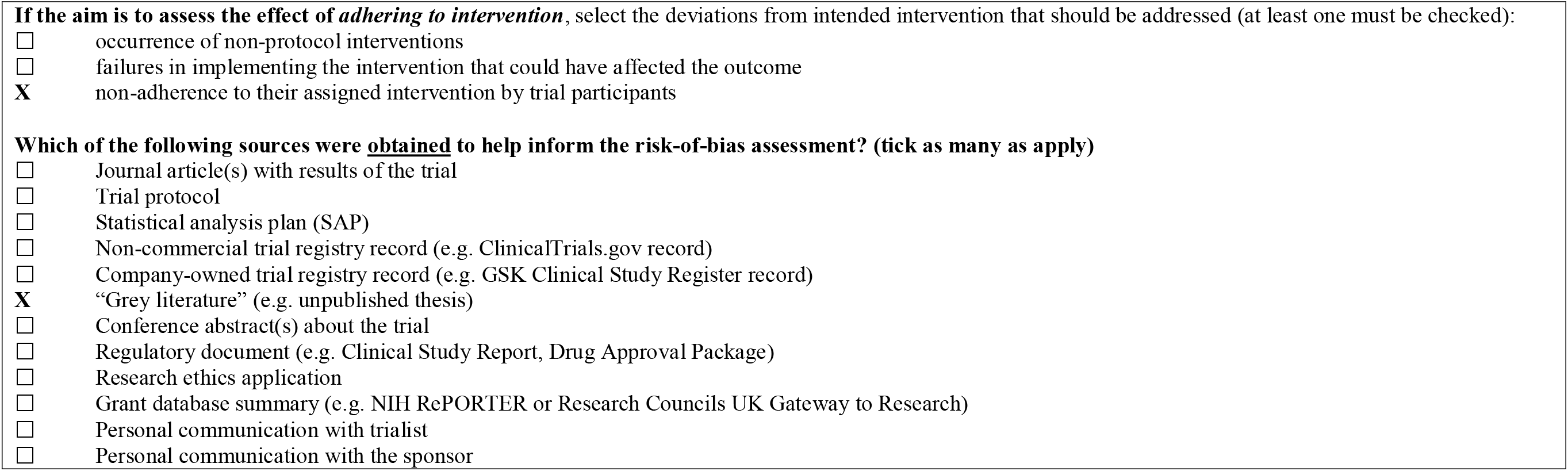
Detailed assessment of the published interventional studies using study quality assessment tools for case-control studies

## Risk of bias assessment

Responses <u>underlined in green</u> are potential markers for low risk of bias, and responses in red are potential markers for a risk of bias. Where questions relate only to sign posts to other questions, no formatting is used.

### Domain 1: Risk of bias arising from the randomization process

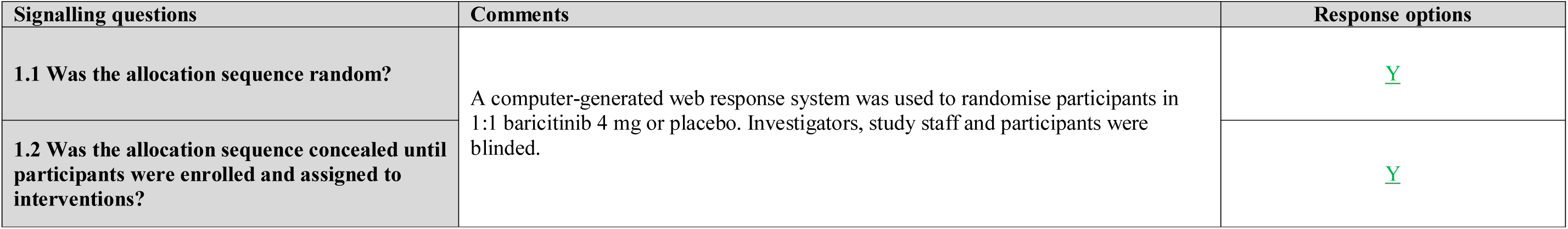
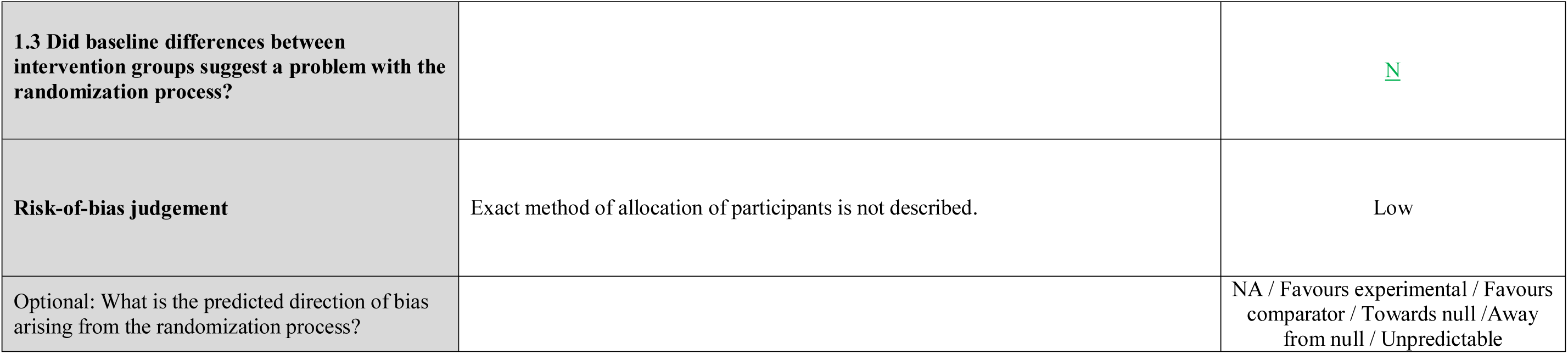

| Signalling questions | Comments | Response options |
| --- | --- | --- |
| 1.1 Was the allocation sequence random? | A computer-generated web response system was used to randomise participants in 1:1 baricitinib 4 mg or placebo. Investigators, study staff and participants were blinded. | <u>Y</u> |
| 1.2 Was the allocation sequence concealed until participants were enrolled and assigned to interventions? |  | <u>Y</u> |
| **1.3 Did baseline differences between intervention groups suggest a problem with the randomization process?** |  | <u>N</u> |
| **Risk-of-bias judgement** | Exact method of allocation of participants is not described. | Low |
| Optional: What is the predicted direction of bias arising from the randomization process? |  | NA / Favours experimental / Favours comparator / Towards null / Away from null / Unpredictable |

### Domain 2: Risk of bias due to deviations from the intended interventions (effect of assignment to intervention)

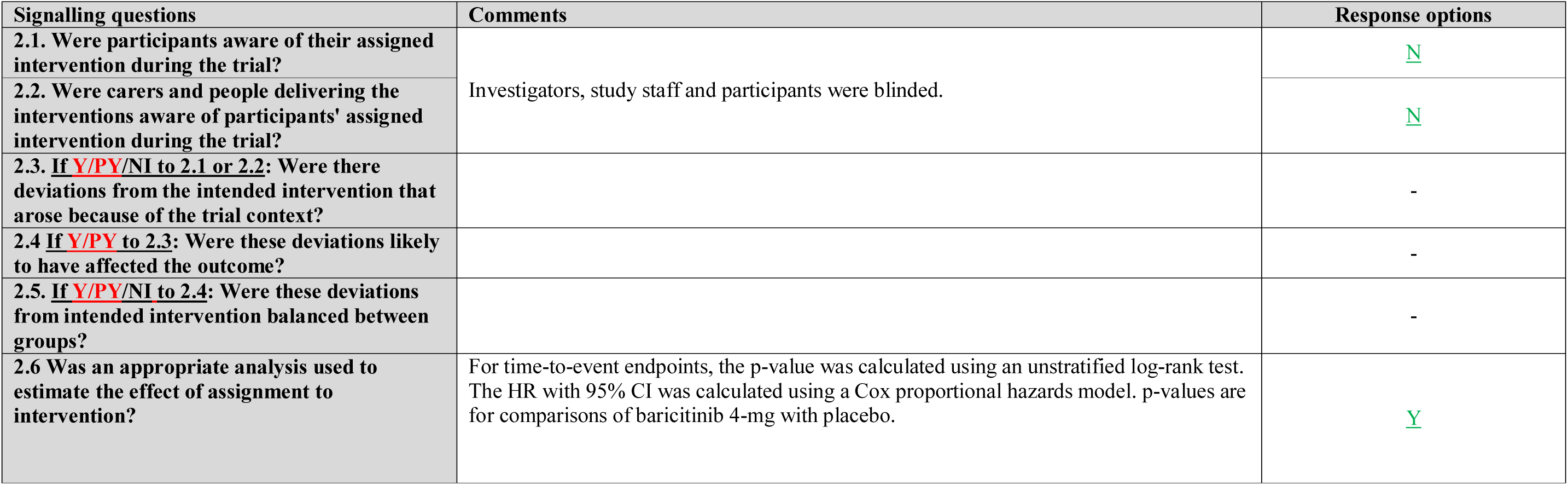
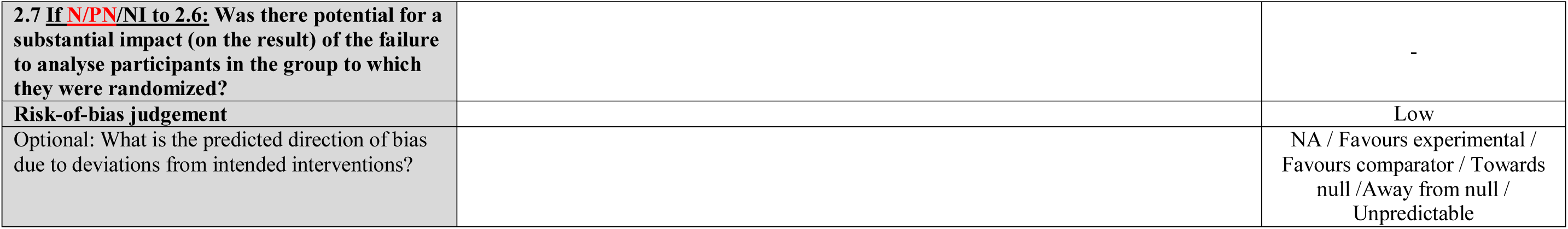

### Domain 2: Risk of bias due to deviations from the intended interventions (effect of adhering to intervention)

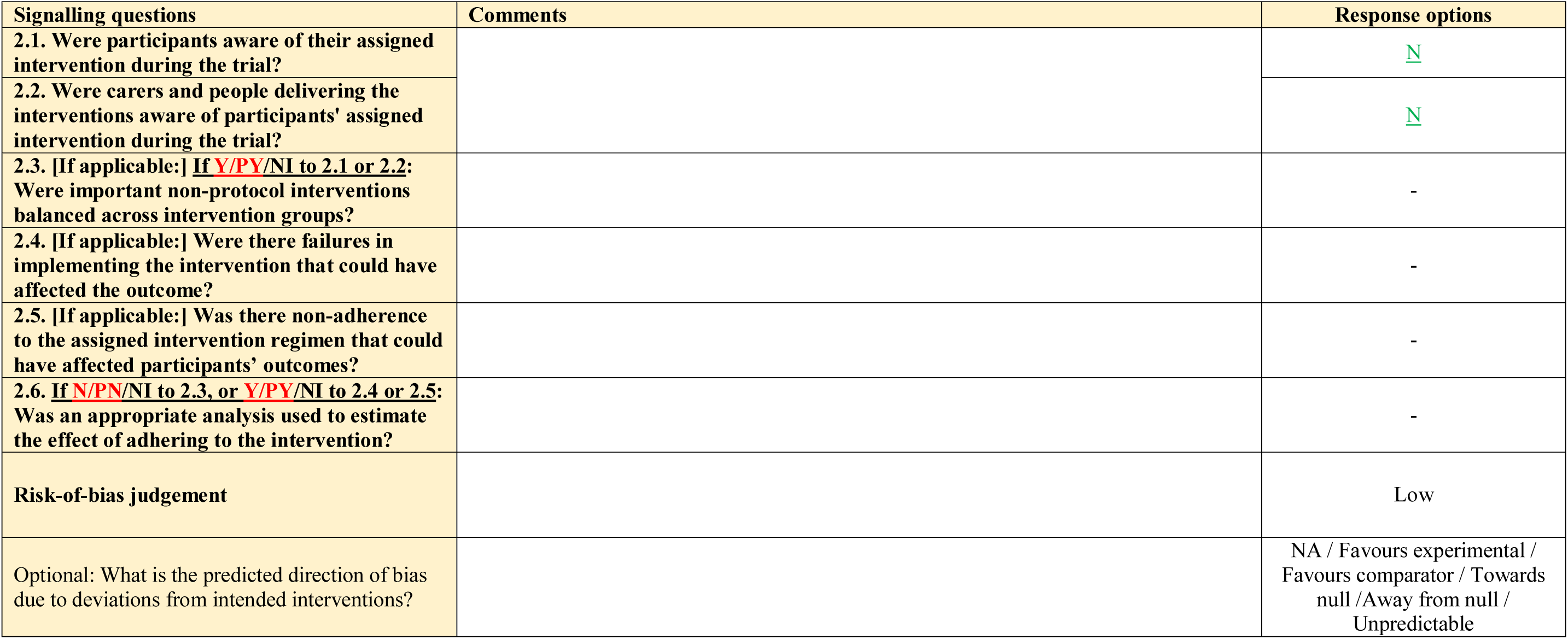

| Signalling questions | Comments | Response options |
| --- | --- | --- |
| **2.1.** Were participants aware of their assigned intervention during the trial? |  | <u>N</u> |
| **2.2.** Were carers and people delivering the interventions aware of participants' assigned intervention during the trial? |  | <u>N</u> |
| **2.3. [If applicable:] If <u>Y/PY/Ni</u> to 2.1 or 2.2:** Were important non-protocol interventions balanced across intervention groups? |  | - |
| **2.4. [If applicable:]** Were there failures in implementing the intervention that could have affected the outcome? |  | - |
| **2.5. [If applicable:]** Was there non-adherence to the assigned intervention regimen that could have affected participants' outcomes? |  | - |
| **2.6. If <u>N/PN/Ni</u> to 2.3, or <u>Y/PY/Ni</u> to 2.4 or 2.5:** Was an appropriate analysis used to estimate the effect of adhering to the intervention? |  | - |
| **Risk-of-bias judgement** |  | Low |
| Optional: What is the predicted direction of bias due to deviations from intended interventions? |  | NA / Favours experimental / Favours comparator / Towards null / Away from null / Unpredictable |

### Domain 3: Missing outcome data

| Signalling questions | Comments | Response options |
| --- | --- | --- |
| 3.1 Were data for this outcome available for all, or nearly all, participants randomized? |  | <u>Y</u> |
| 3.2 If **N/PN/N** to 3.1: Is there evidence that the result was not biased by missing outcome data? |  | NA |
| 3.3 If **N/PN** to 3.2: Could missingness in the outcome depend on its true value? |  | NA |
| 3.4 If **Y/PY/N** to 3.3: Is it likely that missingness in the outcome depended on its true value? |  | NA |
| **Risk-of-bias judgement** | Limitations of this study is the choice of disease progression used as the primary outcome. Disease progression was measured using NIAID-OS (National Institute of Allergy and Infectious Diseases ordinal scale) which reflected on treatment decisions (such as use of certain oxygen delivery devices) and may also be influenced by the heterogeneity of clinical practice across different locations. | Some concerns |
| Optional: What is the predicted direction of bias due to missing outcome data? |  | NA / Favours experimental / Favours comparator / Towards null / Away from null / Unpredictable |

### Domain 4: Risk of bias in measurement of the outcome

| Signalling questions | Comments | Response options |
| --- | --- | --- |
| 4.1 Was the method of measuring the outcome inappropriate? |  | <u>PN</u> |
| 4.2 Could measurement or ascertainment of the outcome have differed between intervention groups? | In the participants were further randomised for receiving SoC (dexamethasone/remdesivir/corticosteroid) which may lead to biased outcome. | <u>Y</u> |
| 4.3 If <u>N/PN/N</u>I to 4.1 and 4.2: Were outcome assessors aware of the intervention received by study participants? |  | - |
| 4.4 If <u>Y/PY/N</u>I to 4.3: Could assessment of the outcome have been influenced by knowledge of intervention received? | Participants receiving different SoC may have influenced the assessment. | NA |
| 4.5 If <u>Y/PY/N</u>I to 4.4: Is it likely that assessment of the outcome was influenced by knowledge of intervention received? |  | <u>PY</u> |
| Risk-of-bias judgement | The anti-inflammatory effects of baricitinib may have had an antagonistic impact in organ systems that are not evaluated by the NIAID-OS. | Some concerns |
| Optional: What is the predicted direction of bias in measurement of the outcome? |  | NA / Favours experimental / Favours comparator / Towards null / Away from null / Unpredictable |

### Domain 5: Risk of bias in selection of the reported result

| Signalling questions | Comments | Response options |
| --- | --- | --- |
| **5.1** Were the data that produced this result analysed in accordance with a pre-specified analysis plan that was finalized before unblinded outcome data were available for analysis? |  | <u>Y</u> |
| Is the numerical result being assessed likely to have been selected, on the basis of the results, from... |  |  |
| **5.2.** ... multiple eligible outcome measurements (e.g. scales, definitions, time points) within the outcome domain? |  | <u>N</u> |
| **5.3** ... multiple eligible analyses of the data? |  | NI |
| **Risk-of-bias judgement** |  | Low |
| Optional: What is the predicted direction of bias due to selection of the reported result? |  | NA / Favours experimental / Favours comparator / Towards null / Away from null / Unpredictable |

### Overall risk of bias

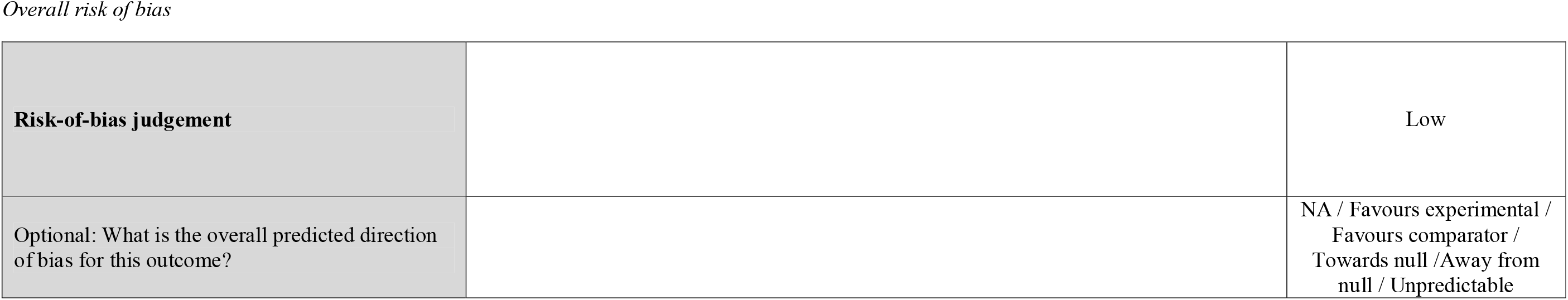
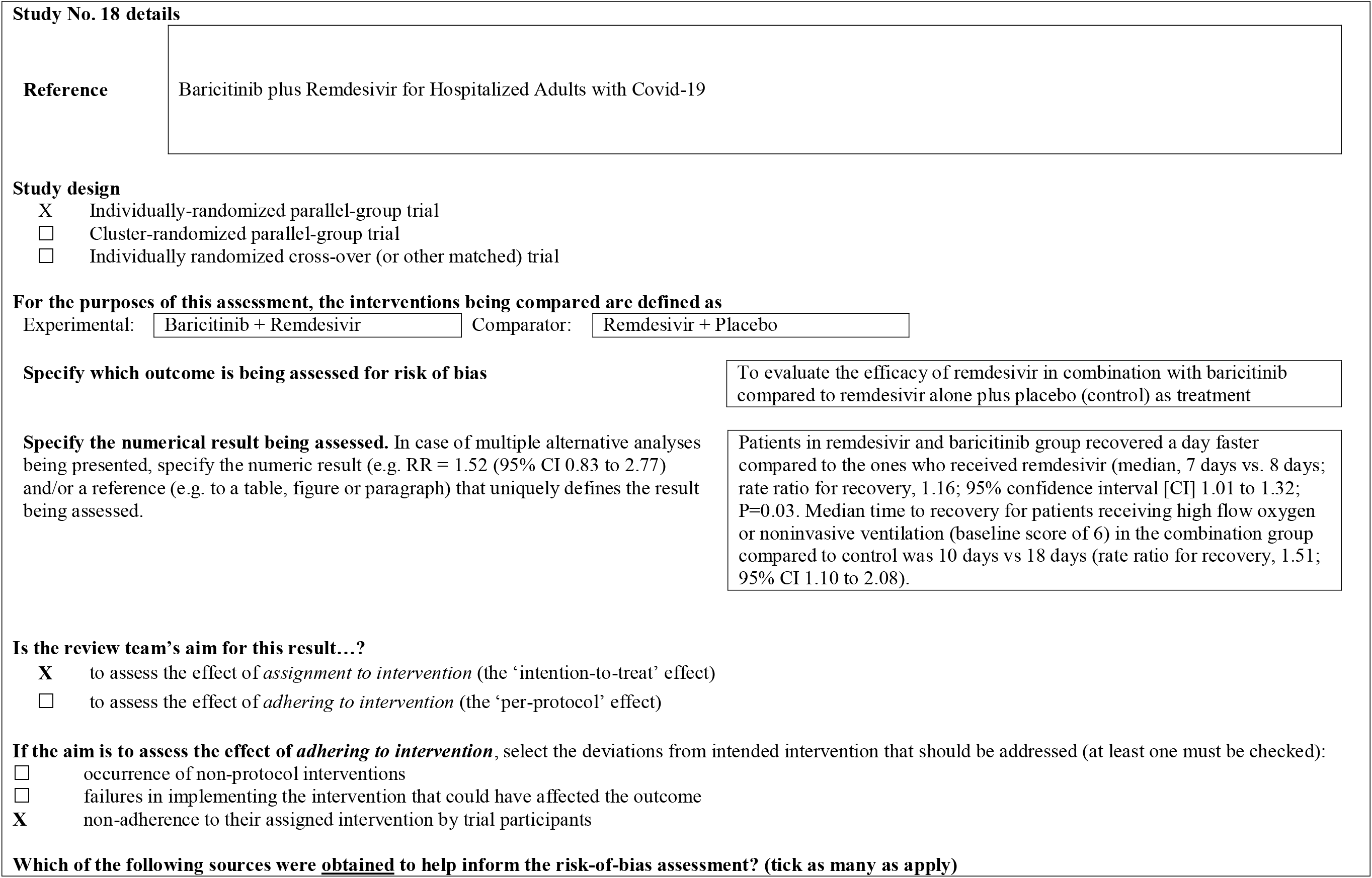
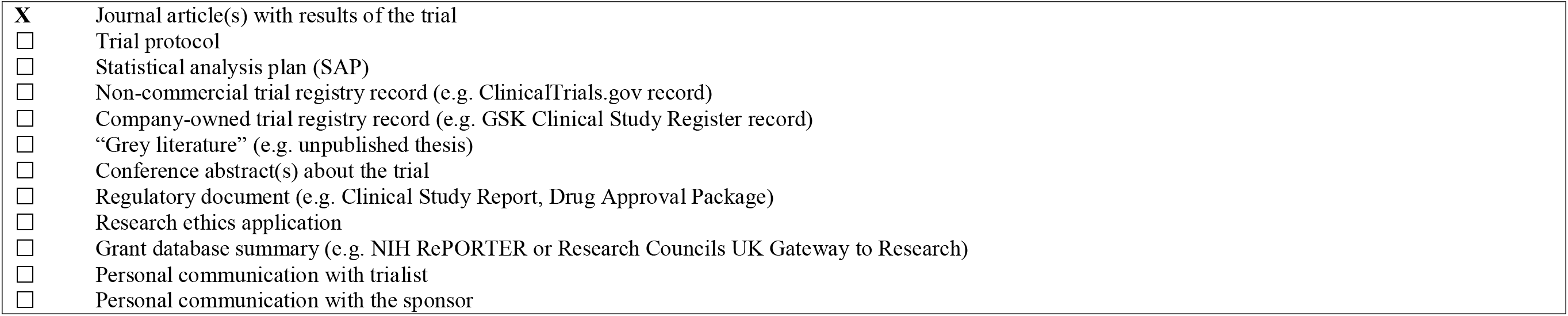

## Risk of bias assessment

### Domain 1: Risk of bias arising from the randomization process

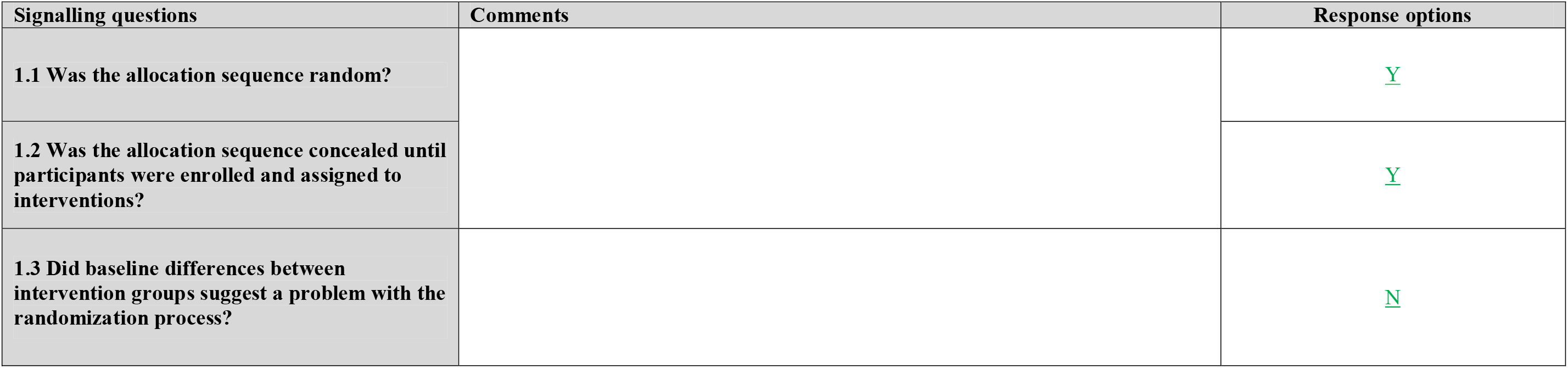
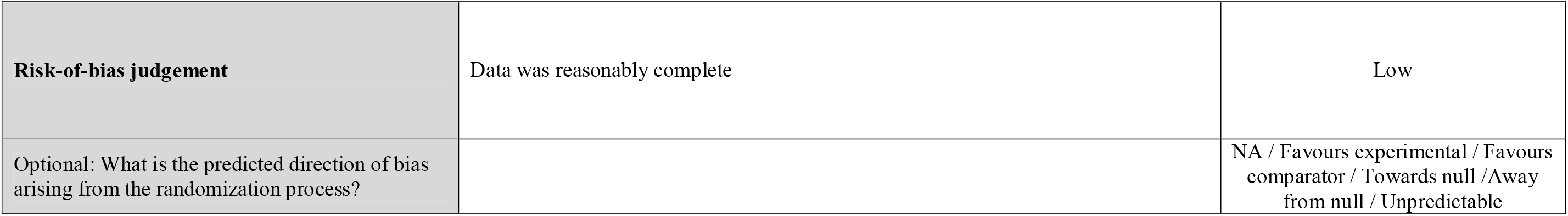

### Domain 2: Risk of bias due to deviations from the intended interventions (effect of assignment to intervention)

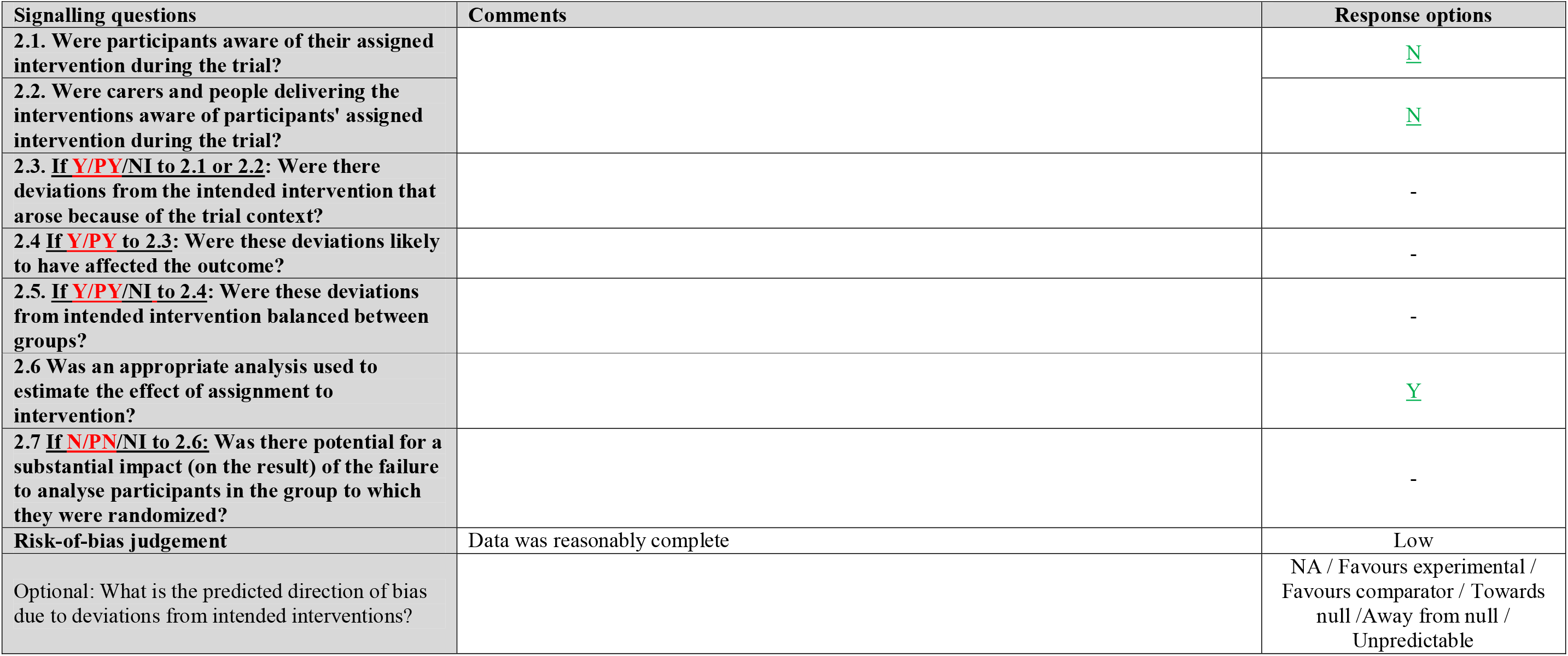

### Domain 2: Risk of bias due to deviations from the intended interventions (effect of adhering to intervention)

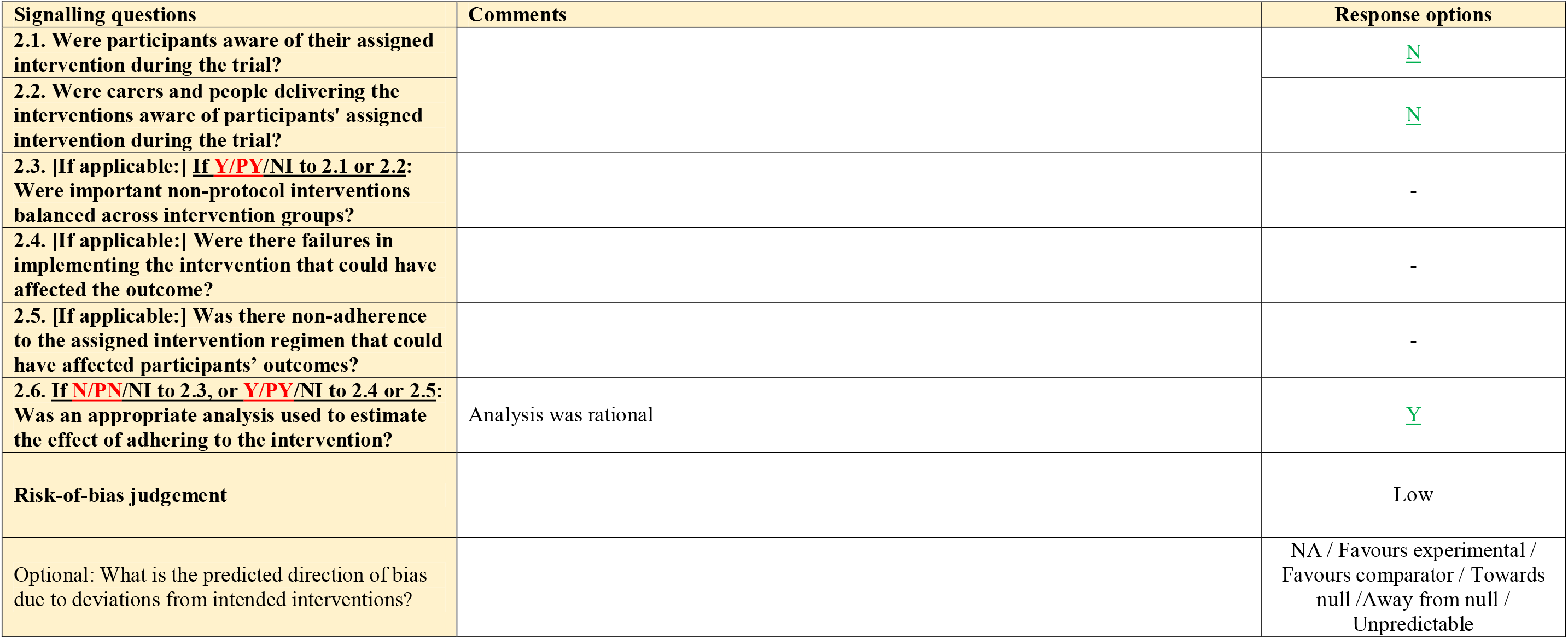

### Domain 3: Missing outcome data

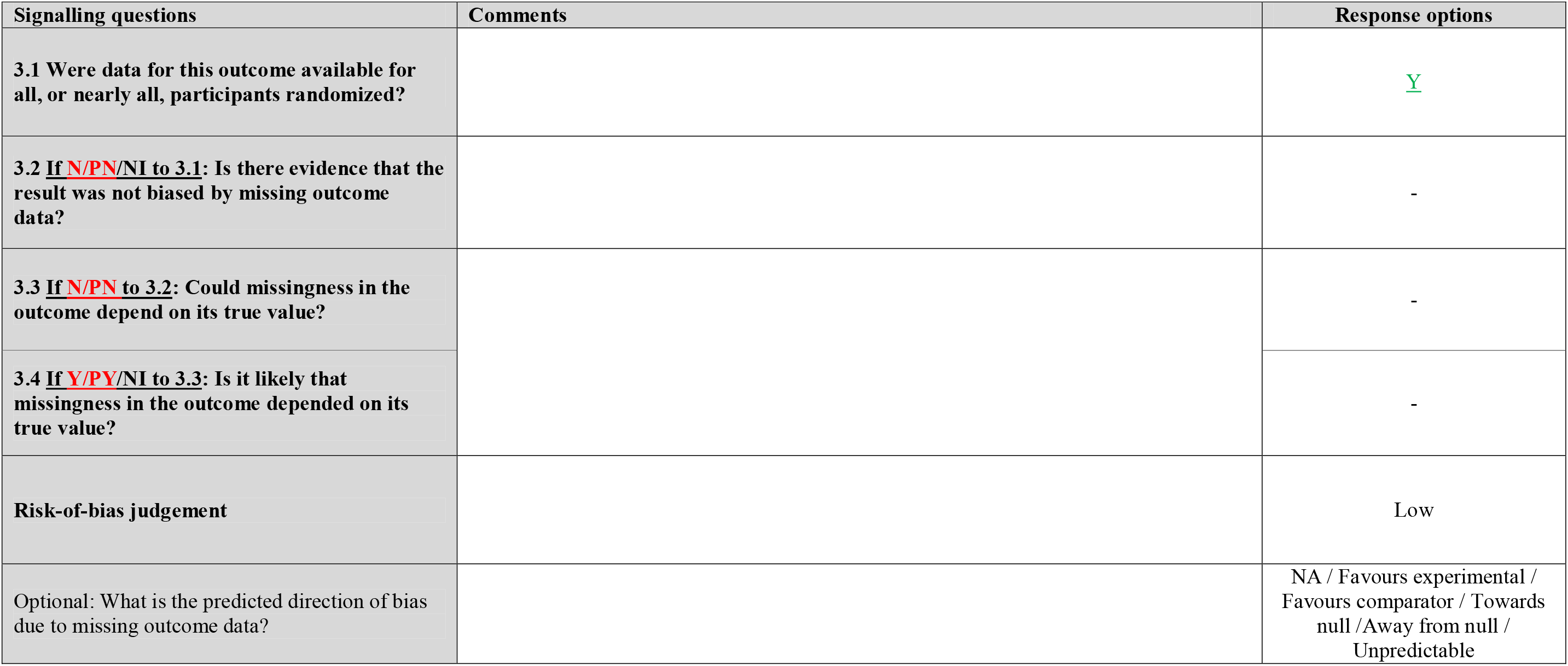

### Domain 4: Risk of bias in measurement of the outcome

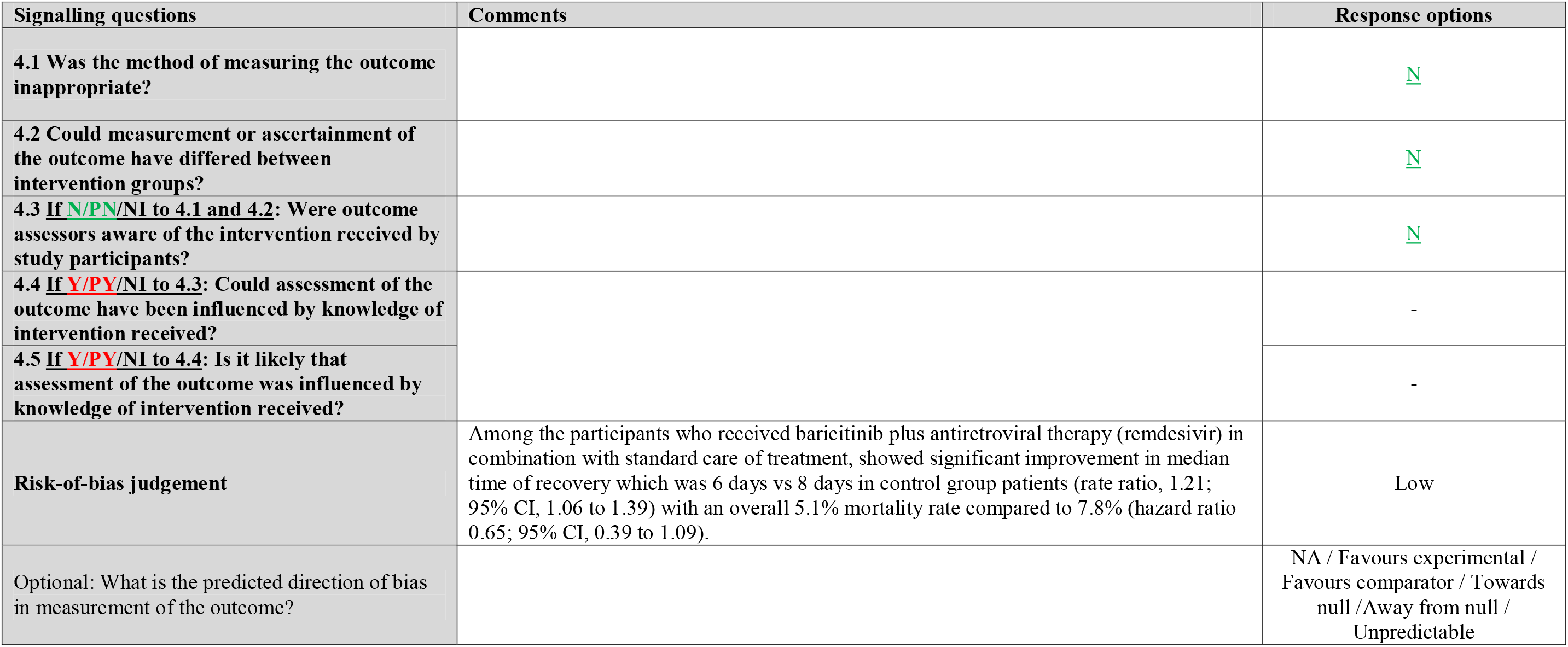

### Domain 5: Risk of bias in selection of the reported result

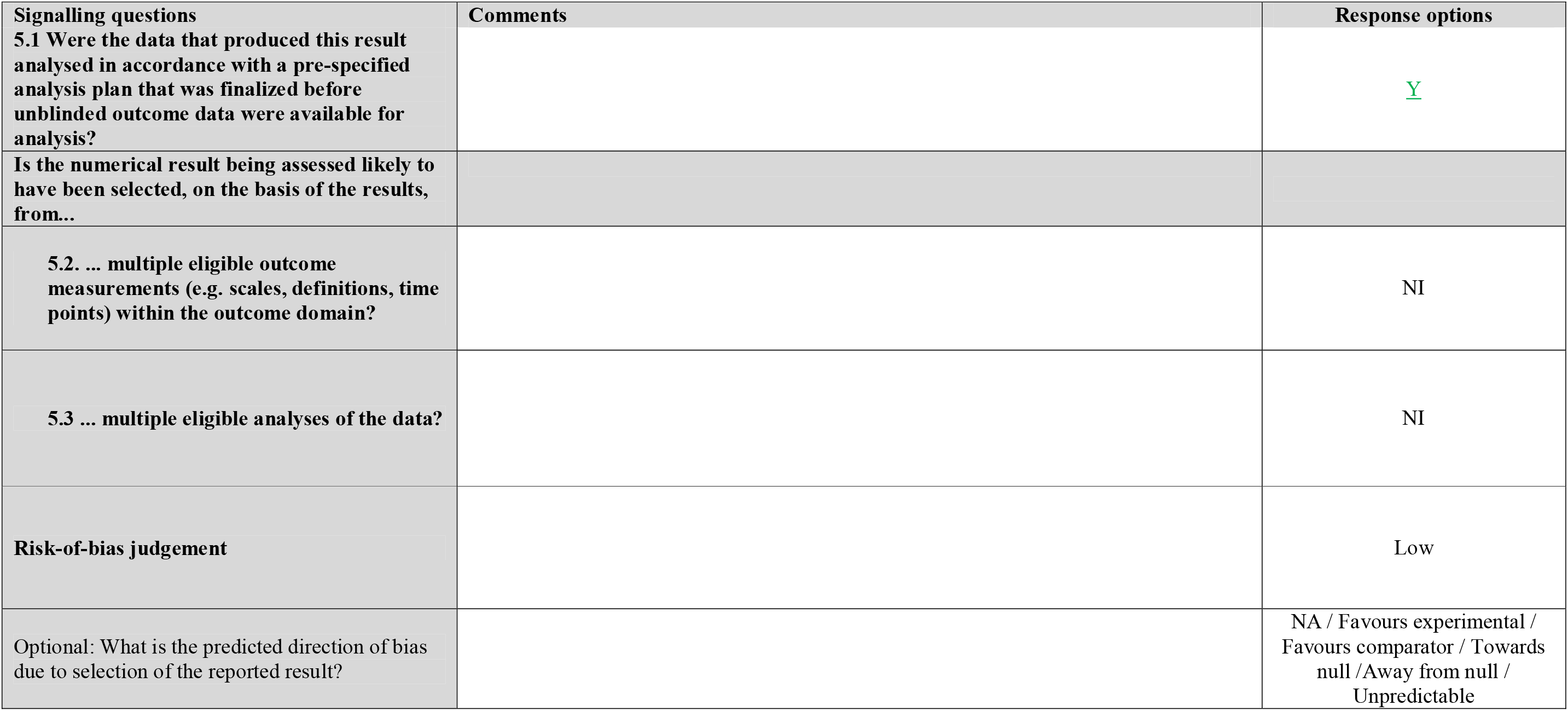

### Overall risk of bias

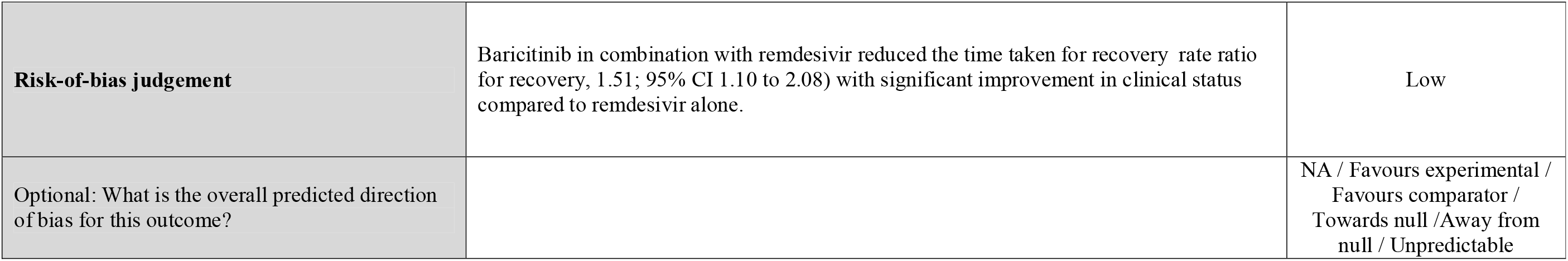
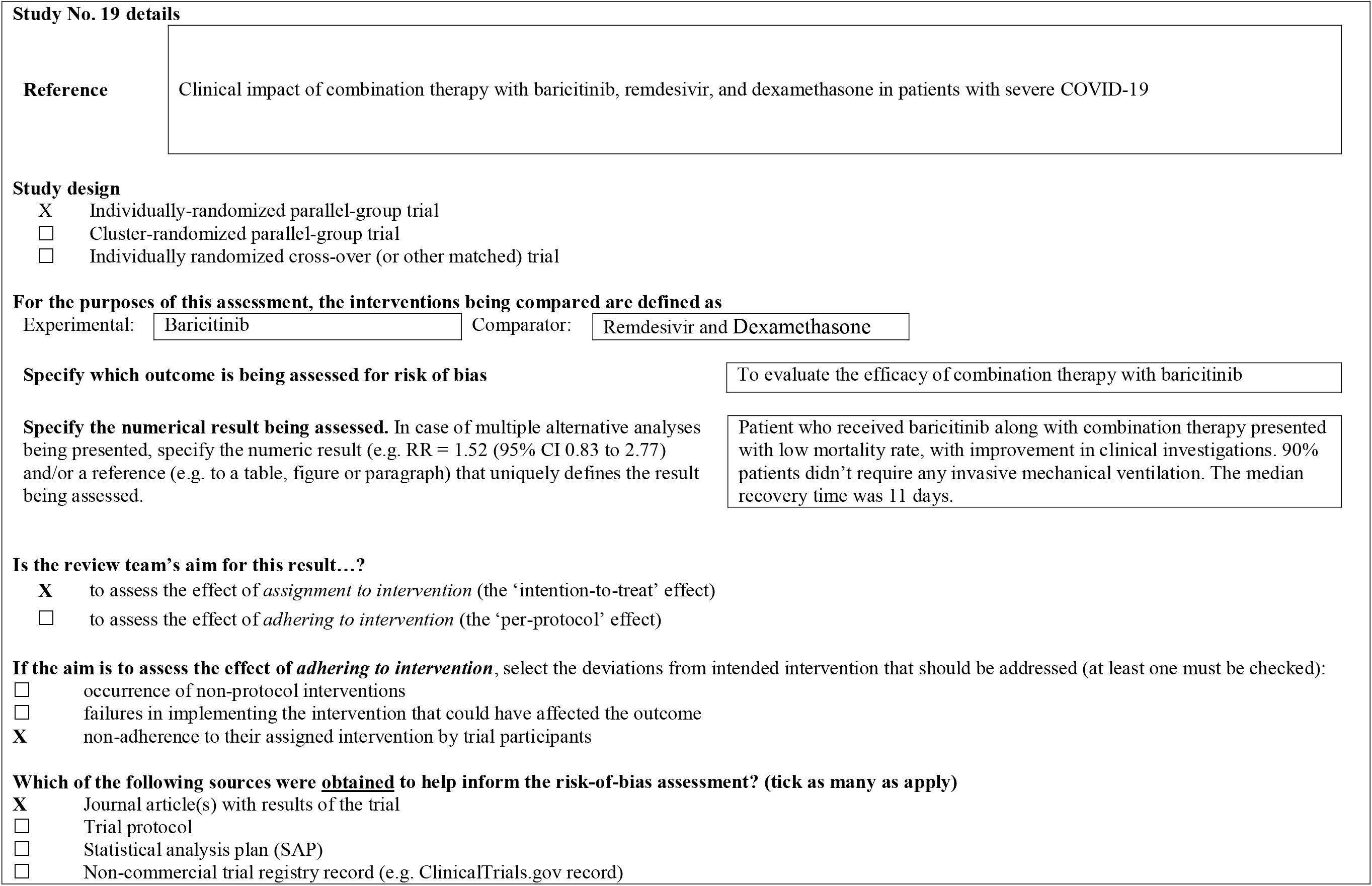
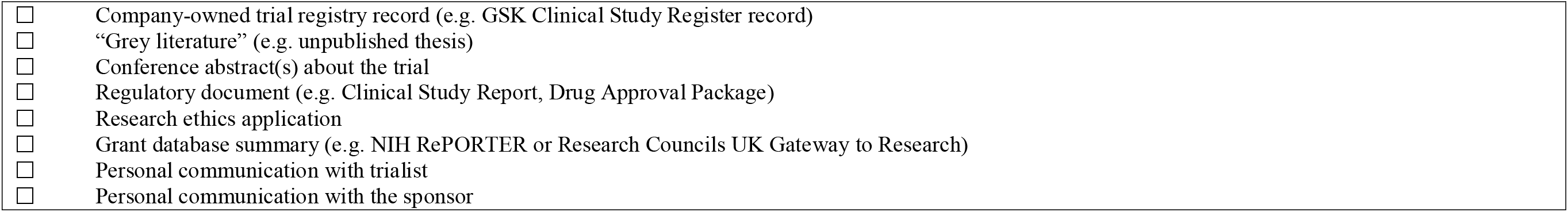

## Risk of bias assessment

### Domain 1: Risk of bias arising from the randomization process

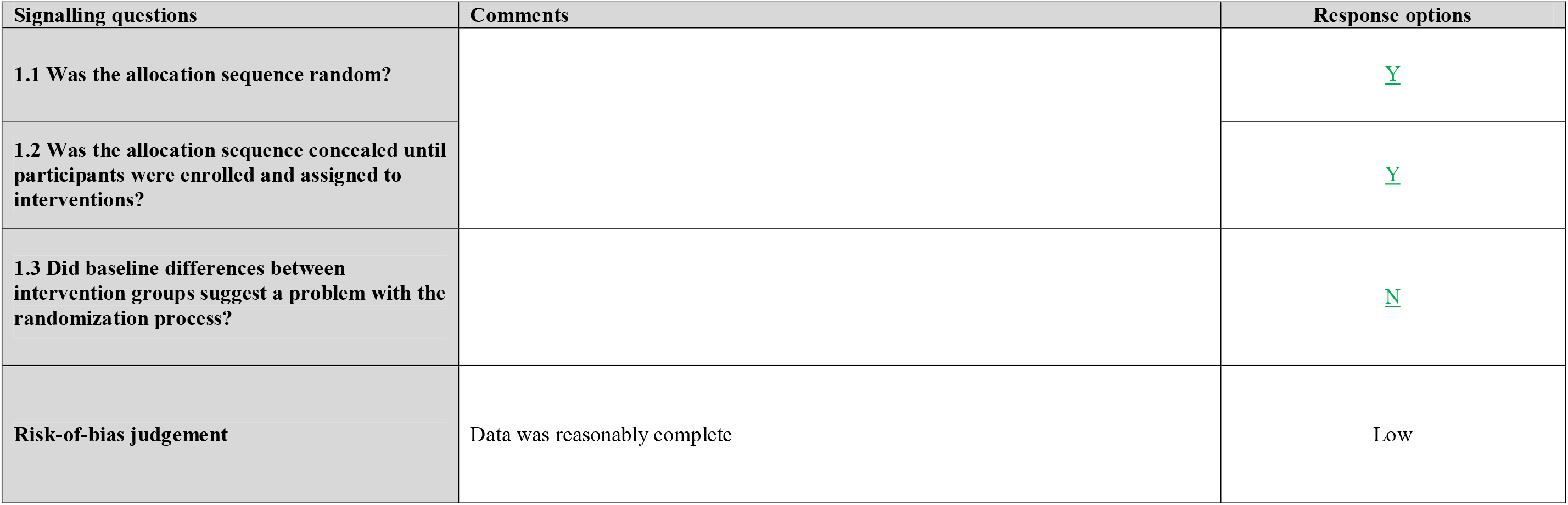
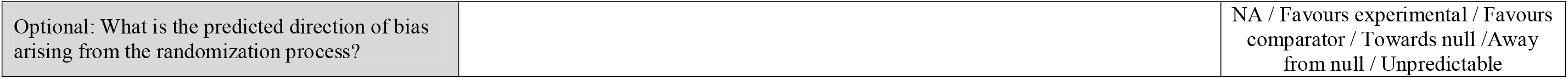

### Domain 2: Risk of bias due to deviations from the intended interventions (effect of assignment to intervention)

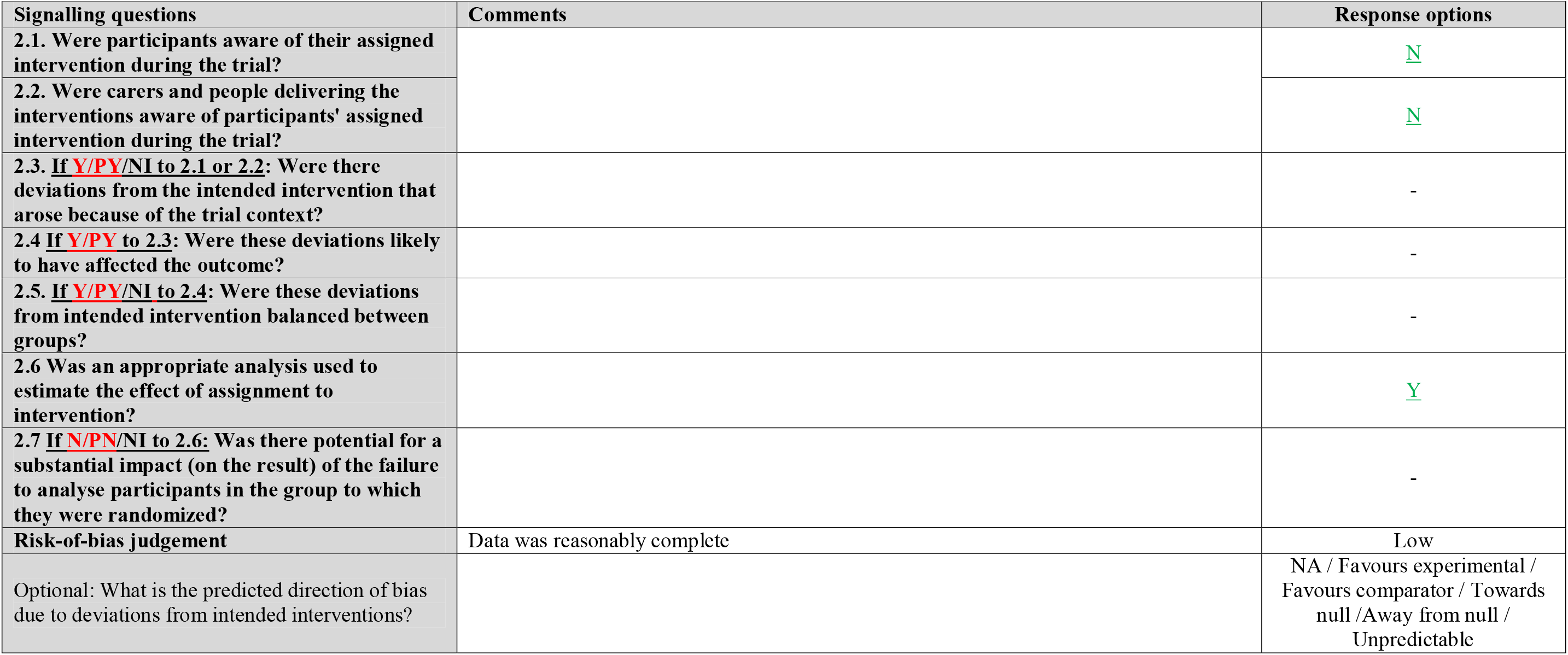

### Domain 2: Risk of bias due to deviations from the intended interventions (effect of adhering to intervention)

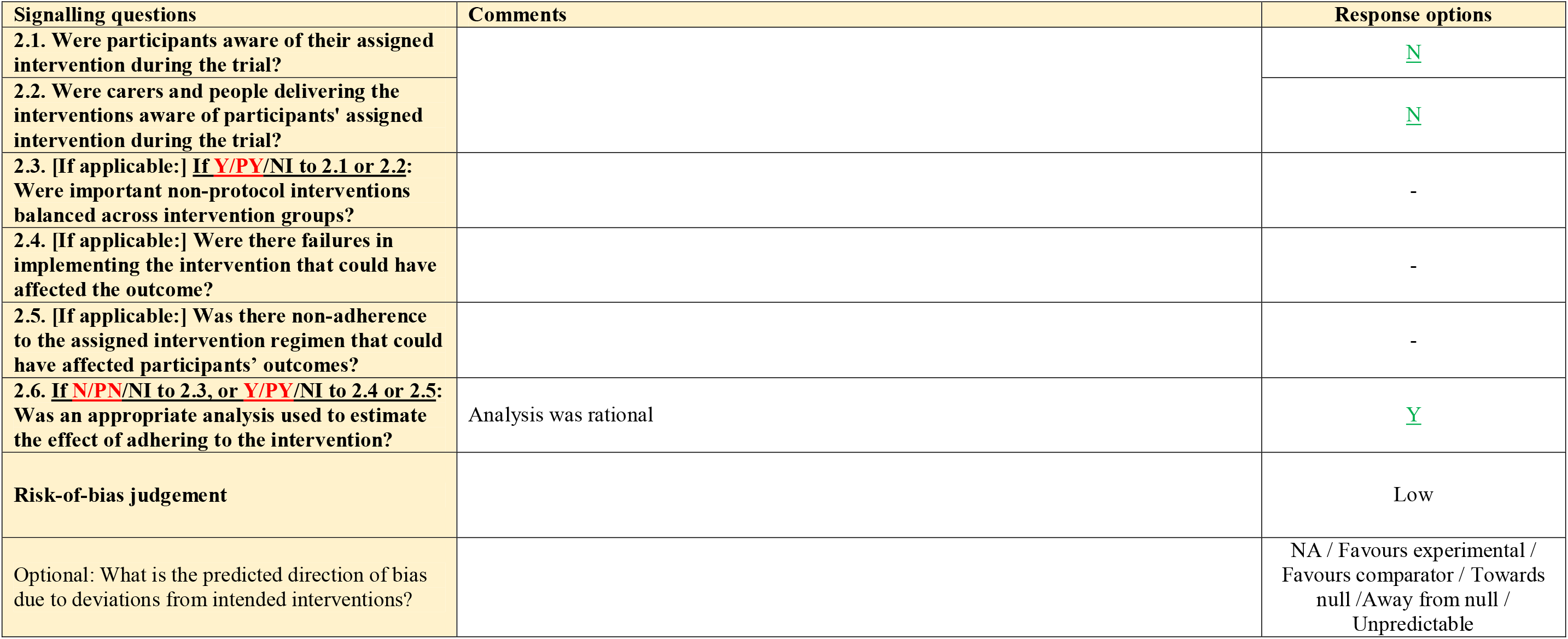

### Domain 3: Missing outcome data

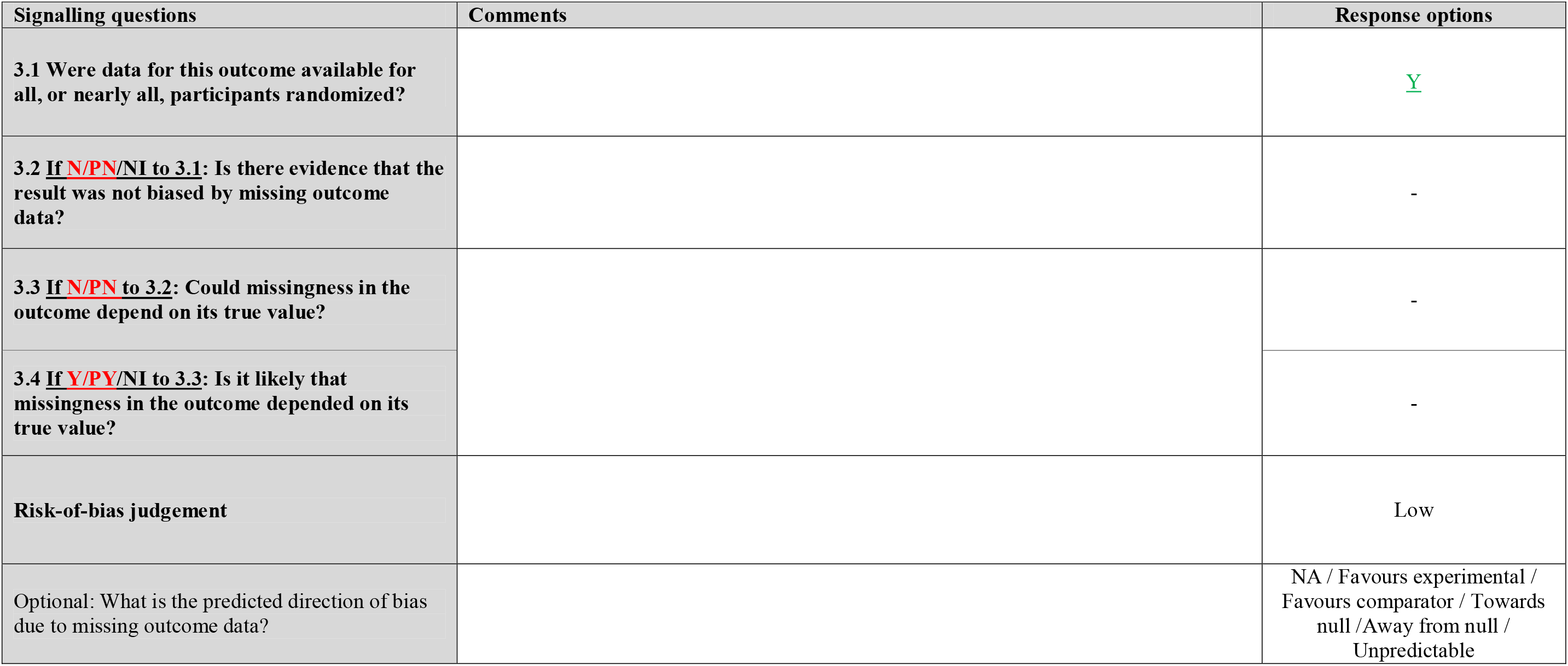

### Domain 4: Risk of bias in measurement of the outcome

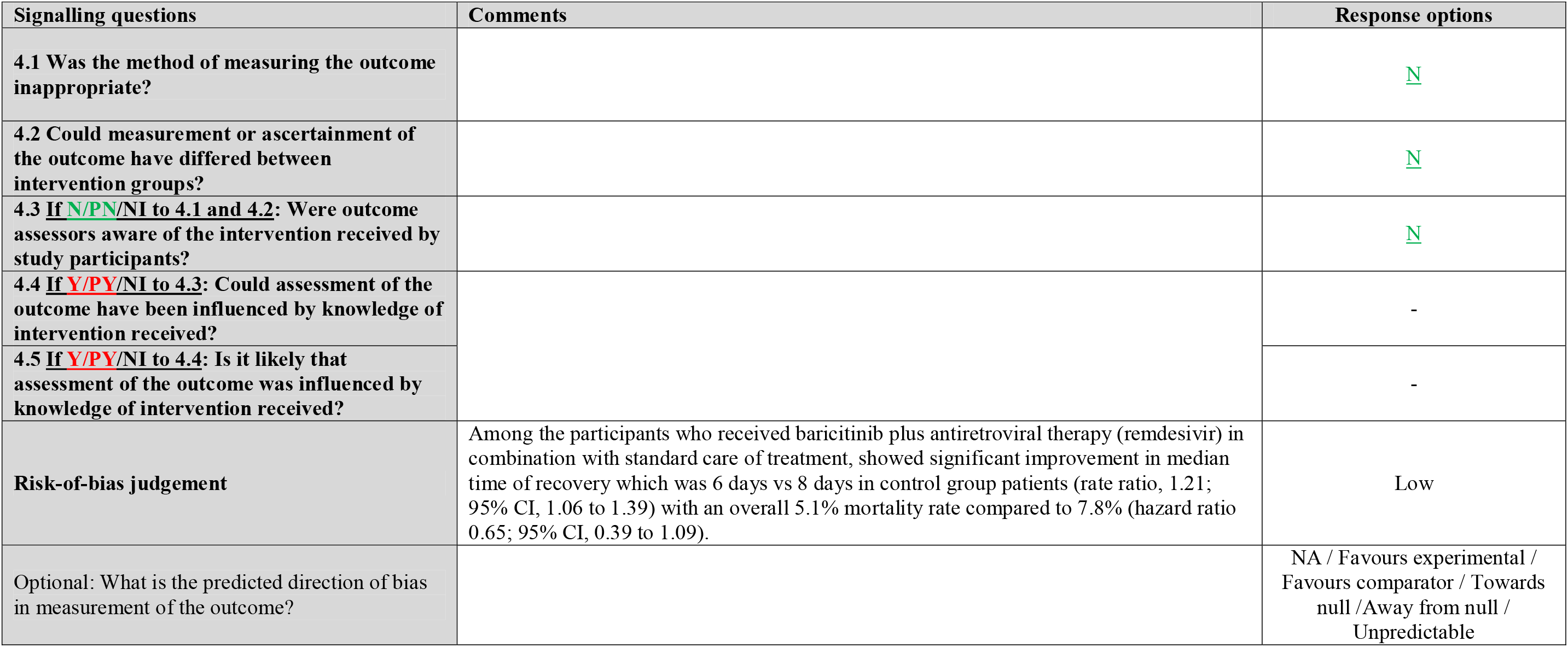

### Domain 5: Risk of bias in selection of the reported result

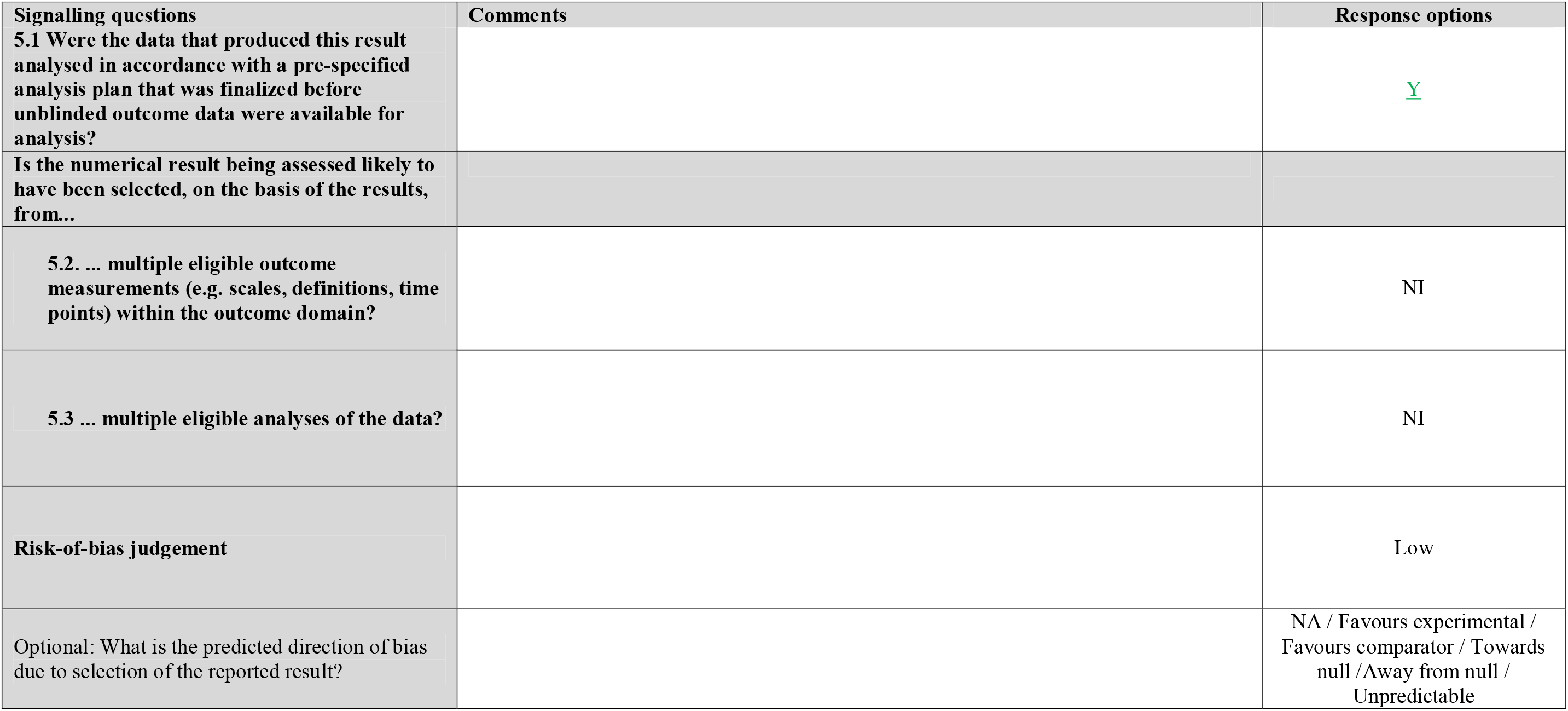

### Overall risk of bias

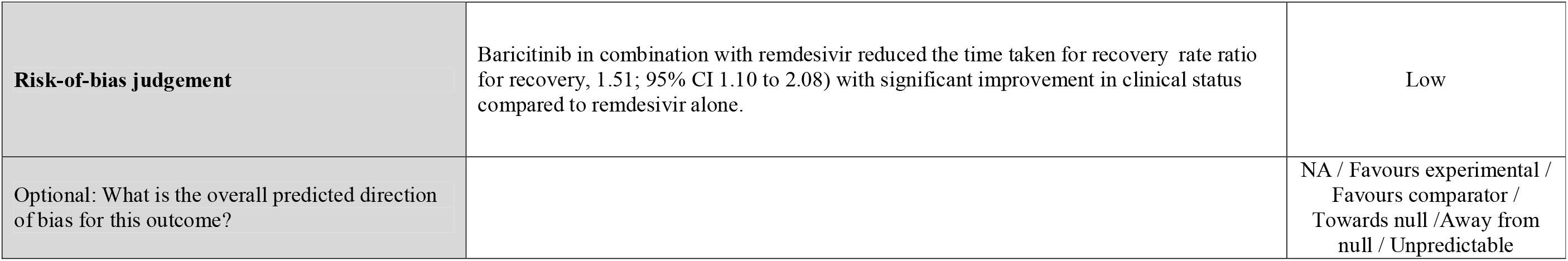

## Reference

1. Coronavirus [Internet]. [cited 2021 Dec 8]. Available from: https://www.who.int/westernpacific/health-topics/coronavirus

2. Huang C, Wang Y, Li X, Ren L, Zhao J, Hu Y, et al. Clinical features of patients infected with 2019 novel coronavirus in Wuhan, China. Lancet Lond Engl. 2020;395(10223):497–506.

3. WHO Coronavirus (COVID-19) Dashboard [Internet]. [cited 2021 Dec 8]. Available from: https://covid19.who.int

4. Guidelines Introduction [Internet]. COVID-19 Treatment Guidelines. [cited 2021 May 29]. Available from: https://www.covid19treatmentguidelines.nih.gov/introduction/

5. Commissioner O of the. Coronavirus (COVID-19) Update: FDA Authorizes Drug Combination for Treatment of COVID-19 [Internet]. FDA. FDA; 2020 [cited 2021 Jun 8]. Available from: https://www.fda.gov/news-events/press-announcements/coronavirus-covid-19-update-fda-authorizes-drug-combination-treatment-covid-19

6. PINHO AC. EMA starts evaluating use of Olumiant in hospitalised COVID-19 patients requiring supplemental oxygen [Internet]. European Medicines Agency. 2021 [cited 2021 Jun 8]. Available from: https://www.ema.europa.eu/en/news/ema-starts-evaluating-use-olumiant-hospitalised-covid-19-patients-requiring-supplemental-oxygen

7. Lilly accelerating baricitinib’s availability in India following receipt of permission for restricted emergency use as a COVID-19 therapy via donations and licensing agreements | Eli Lilly and Company [Internet]. [cited 2021 May 29]. Available from: https://investor.lilly.com/news-releases/news-release-details/lilly-accelerating-baricitinibs-availability-india-following

8. Markham A. Baricitinib: First Global Approval. Drugs. 2017 Apr;77(6):697–704.

9. Gao Q, Liang X, Shaikh AS, Zang J, Xu W, Zhang Y. JAK/STAT Signal Transduction: Promising Attractive Targets for Immune, Inflammatory and Hematopoietic Diseases. Curr Drug Targets. 2018 Mar 19;19(5):487–500.

10. Damsky W, Peterson D, Ramseier J, Al-Bawardy B, Chun H, Proctor D, et al. The emerging role of Janus kinase inhibitors in the treatment of autoimmune and inflammatory diseases. J Allergy Clin Immunol. 2021 Mar;147(3):814–26.

11. Mehta P, McAuley DF, Brown M, Sanchez E, Tattersall RS, Manson JJ. COVID-19: consider cytokine storm syndromes and immunosuppression. The Lancet. 2020 Mar;395(10229):1033–4.

12. Cantini F, Niccoli L, Matarrese D, Nicastri E, Stobbione P, Goletti D. Baricitinib therapy in COVID-19: A pilot study on safety and clinical impact. J Infect. 2020 Aug 1;81(2):318–56.

13. Cantini F, Niccoli L, Nannini C, Matarrese D, Natale MED, Lotti P, et al. Beneficial impact of Baricitinib in COVID-19 moderate pneumonia; multicentre study. J Infect. 2020 Oct;81(4):647–79.

14. Seif F, Aazami H, Khoshmirsafa M, Kamali M, Mohsenzadegan M, Pornour M, et al. JAK Inhibition as a New Treatment Strategy for Patients with COVID-19. Int Arch Allergy Immunol. 2020;181(6):467–75.

15. Spinelli FR, Conti F, Gadina M. HiJAKing SARS-CoV-2? The potential role of JAK inhibitors in the management of COVID-19. Sci Immunol. 2020 May 8;5(47).

16. WHO-COVID-19: Global literature on coronavirus disease [Internet]. [cited 2021 Jun 8]. Available from: https://search.bvsalud.org/global-literature-on-novel-coronavirus-2019-ncov/?u_filter%5B%5D=fulltext&u_filter%5B%5D=db&u_filter%5B%5D=collection&u_filter%5B%5D=mj_cluster&u_filter%5B%5D=type_of_study&u_filter%5B%5D=clinical_aspect&u_filter%5B%5D=la&u_filter%5B%5D=year_cluster&u_filter%5B%5D=ta_cluster&fb=ta_cluster%3A70&output=site&lang=en&from=1&sort=DATENTRY_DESC&format=summary&count=20&page=1&skfp=&index=tw&q=Baricitinib&where=&range_year_start=&range_year_end=&range_year_start=&range_year_end=#ta_cluster

17. Clinical Spectrum [Internet]. COVID-19 Treatment Guidelines. [cited 2021 Dec 8]. Available from: https://www.covid19treatmentguidelines.nih.gov/overview/clinical-spectrum/

18. Cochrane T. Assessing risk of bias in a randomized trial [Internet]. [cited 2021 Jun 8]. Available from: /handbook/current/chapter-08

19. Study Quality Assessment Tools | NHLBI, NIH [Internet]. Available from: https://www.nhlbi.nih.gov/health-topics/study-quality-assessment-tools

20. Hasan MdJ, Rabbani R, Anam AM, Huq SMR, Polash MMI, Nessa SST, et al. Impact of high dose of baricitinib in severe COVID-19 pneumonia: a prospective cohort study in Bangladesh. BMC Infect Dis. 2021 May 7;21:427.

21. Hasan MJ, Rabbani R, Anam AM, Huq SMR. Additional baricitinib loading dose improves clinical outcome in COVID-19. Open Med. 2021 Jan 1;16(1):041–6.

22. Titanji BK, Farley MM, Mehta A, Connor-Schuler R, Moanna A, Cribbs SK, et al. Use of Baricitinib in Patients With Moderate to Severe Coronavirus Disease 2019. Clin Infect Dis. 2021 Apr 1;72(7):1247–50.

23. Bronte V, Ugel S, Tinazzi E, Vella A, Sanctis FD, Canè S, et al. Baricitinib restrains the immune dysregulation in patients with severe COVID-19. J Clin Invest. 2020 Dec 1;130(12):6409–16.

24. Falcone M, Tiseo G, Barbieri G, Galfo V, Russo A, Virdis A, et al. Role of Low-Molecular-Weight Heparin in Hospitalized Patients With Severe Acute Respiratory Syndrome Coronavirus 2 Pneumonia: A Prospective Observational Study. Open Forum Infect Dis [Internet]. 2020 Dec 1 [cited 2021 Jul 27];7(12). Available from: https://doi.org/10.1093/ofid/ofaa563

25. Rosas J, Liaño FP, Cantó ML, Barea JMC, Beser AR, Rabasa JTA, et al. Experience With the Use of Baricitinib and Tocilizumab Monotherapy or Combined, in Patients With Interstitial Pneumonia Secondary to Coronavirus COVID19: A Real-World Study. Reumatol Clínica [Internet]. 2020 Nov 28 [cited 2021 Jul 19]; Available from: https://www.sciencedirect.com/science/article/pii/S1699258X20302710

26. Milligan PS, Amsden J, Norris SA, Baker RL, Myers J, Preston F, et al. Clinical Outcomes in a Cohort of Non-Ventilated COVID-19 Patients with Progressive Hypoxemia and Hyper-Inflammatory Response Treated with Baricitinib [Internet]. Rochester, NY: Social Science Research Network; 2020 May [cited 2021 Jul 19]. Report No.: ID 3594565. Available from: https://papers.ssrn.com/abstract=3594565

27. Santos CS, Férnandez XC, Moriano Morales C, Álvarez ED, Álvarez Castro C, López Robles A, et al. Biological agents for rheumatic diseases in the outbreak of COVID-19: friend or foe? RMD Open. 2021 Jan;7(1):e001439.

28. Pérez-Alba E, Nuzzolo-Shihadeh L, Aguirre-García GM, Espinosa-Mora J, Lecona-Garcia JD, Flores-Pérez RO, et al. Baricitinib plus dexamethasone compared to dexamethasone for the treatment of severe COVID-19 pneumonia: A retrospective analysis. J Microbiol Immunol Infect Wei Mian Yu Gan Ran Za Zhi. 2021 Oct;54(5):787–93.

29. Amarnath A, Das A, Mutya V, Ibrahim I. The Effect Of Baricitinib Usage On The Clinical And Biochemical Profiles Of Covid-19 Patients- A Retrospective Observational Study. Cold Spring Harb Lab. 2021;

30. García-García JA, Pérez-Quintana M, Ramos-Giráldez C, Cebrián-González I, Martín-Ponce ML, Del Valle-Villagrán J, et al. Anakinra versus Baricitinib: Different Strategies for Patients Hospitalized with COVID-19. J Clin Med. 2021 Sep 6;10(17):4019.

31. Rodriguez-Garcia JL, Sanchez-Nievas G, Arevalo-Serrano J, Garcia-Gomez C, Jimenez-Vizuete JM, Martinez-Alfaro E. Baricitinib improves respiratory function in patients treated with corticosteroids for SARS-CoV-2 pneumonia: an observational cohort study. Rheumatol Oxf Engl. 2021 Jan 5;60(1):399–407.

32. Iglesias Gómez R, Méndez R, Palanques-Pastor T, Ballesta-López O, Borrás Almenar C, Megías Vericat JE, et al. Baricitinib against severe COVID-19: effectiveness and safety in hospitalised pretreated patients. Eur J Hosp Pharm Sci Pract. 2021 Jul 28;ejhpharm-2021-002741.

33. Abizanda P, Calbo Mayo JM, Mas Romero M, Cortés Zamora EB, Tabernero Sahuquillo MT, Romero Rizos L, et al. Baricitinib reduces 30-day mortality in older adults with moderate-to-severe COVID-19 pneumonia. J Am Geriatr Soc. 2021 Oct;69(10):2752–8.

34. Marconi VC, Ramanan AV, Bono S de, Kartman CE, Krishnan V, Liao R, et al. Efficacy and safety of baricitinib in patients with COVID-19 infection: Results from the randomised, double-blind, placebo-controlled, parallel-group COV-BARRIER phase 3 trial. medRxiv. 2021 May 30;2021.04.30.21255934.

35. Frost MT, Jimenez-Solem E, Ankarfeldt MZ, Nyeland ME, Andreasen AH, Petersen TS. The Adaptive COVID-19 Treatment Trial-1 (ACTT-1) in a real-world population: a comparative observational study. Crit Care Lond Engl. 2020 Dec 7;24(1):677.

36. Kalil AC, Patterson TF, Mehta AK, Tomashek KM, Wolfe CR, Ghazaryan V, et al. Baricitinib plus Remdesivir for Hospitalized Adults with Covid-19. N Engl J Med. 2021 Mar 4;384(9):795–807.

37. Izumo T, Kuse N, Awano N, Tone M, Sakamoto K, Takada K, et al. Clinical impact of combination therapy with baricitinib, remdesivir, and dexamethasone in patients with severe COVID-19. Respir Investig. 2021 Nov;59(6):799–803.

38. Risk of bias tools - RoB 2 tool [Internet]. [cited 2021 Jul 31]. Available from: https://www.riskofbias.info/welcome/rob-2-0-tool

39. Hall F. mulTi-Arm Therapeutic Study in Pre-ICu Patients Admitted With Covid-19 - Repurposed Drugs (TACTIC-R) [Internet]. clinicaltrials.gov; 2020 May [cited 2021 Jun 30]. Report No.: NCT04390464. Available from: https://clinicaltrials.gov/ct2/show/NCT04390464

40. National Institute of Allergy and Infectious Diseases (NIAID). A Multicenter, Adaptive, Randomized Blinded Controlled Trial of the Safety and Efficacy of Investigational Therapeutics for the Treatment of COVID-19 in Hospitalized Adults (ACTT-4) [Internet]. clinicaltrials.gov; 2021 May [cited 2021 Jun 10]. Report No.: NCT04640168. Available from: https://clinicaltrials.gov/ct2/show/NCT04640168

41. Tombetti E. Factorial, Multicentric, Randomized Clinical Trial of Remdesivir and Immunotherapy in Combination With Dexamethasone for Moderate COVID-19 (the AMMURAVID Trial) [Internet]. clinicaltrials.gov; 2021 Apr [cited 2021 Jun 15]. Report No.: NCT04832880. Available from: https://clinicaltrials.gov/ct2/show/NCT04832880

42. Cumulative adaptive, multiarm, multistage and multicentre randomized clinical trial with immunotherapy for Moderate COVID-19 [Internet]. [cited 2021 Jul 3]. Available from: https://www.clinicaltrialsregister.eu/ctr-search/trial/2020-001854-23/IT

43. Cantini F. Baricitinib Combined With Antiviral Therapy in Symptomatic Patients Infected by COVID-19: an Open-label, Pilot Study [Internet]. clinicaltrials.gov; 2020 Apr [cited 2021 Jun 13]. Report No.: NCT04320277. Available from: https://clinicaltrials.gov/ct2/show/NCT04320277

44. University of Colorado, Denver. Safety and Efficacy of Baricitinib for COVID-19 [Internet]. clinicaltrials.gov; 2021 Mar [cited 2021 Jun 13]. Report No.: NCT04340232. Available from: https://clinicaltrials.gov/ct2/show/NCT04340232

45. Baricitinib for coRona Virus pnEumonia (COVID-19): a THerapeutic Trial - Full Text View - ClinicalTrials.gov [Internet]. [cited 2021 Jul 21]. Available from: https://clinicaltrials.gov/ct2/show/NCT04399798

46. Menichetti F. BARICIVID-19 STUDY: MultiCentre, Randomised, Phase IIa Clinical Trial Evaluating Efficacy and Tolerability of Baricitinib as add-on Treatment of In-patients With COVID-19 Compared to Standard Therapy [Internet]. clinicaltrials.gov; 2020 May [cited 2021 Jun 10]. Report No.: NCT04393051. Available from: https://clinicaltrials.gov/ct2/show/NCT04393051

47. University of Southern California. A Phase II Randomized Double-Blind Trial of Baricitinib or Placebo Combined With Antiviral Therapy in Patients With Moderate and Severe COVID-19 [Internet]. clinicaltrials.gov; 2021 May [cited 2021 Jun 13]. Report No.: NCT04373044. Available from: https://clinicaltrials.gov/ct2/show/NCT04373044

48. Hospital Universitario de Fuenlabrada. Prospective, Phase II, Randomized, Open-label, Parallel Group Study to Evaluate the Efficacy of Baricitinib, Imatinib or Supportive Treatment in Patients With SARS Cov2 Pneumonia [Internet]. clinicaltrials.gov; 2021 Feb [cited 2021 Jun 13]. Report No.: NCT04346147. Available from: https://clinicaltrials.gov/ct2/show/NCT04346147

49. Barrett L. Treatment of Moderate to Severe Coronavirus Disease (COVID-19) in Hospitalized Patients [Internet]. clinicaltrials.gov; 2020 Jul [cited 2021 Jun 13]. Report No.: NCT04321993. Available from: https://clinicaltrials.gov/ct2/show/NCT04321993

50. Moreno-González G, Mussetti A, Albasanz-Puig A, Salvador I, Sureda A, Gudiol C, et al. A Phase I/II Clinical Trial to evaluate the efficacy of baricitinib to prevent respiratory insufficiency progression in onco- hematological patients affected with COVID19: A structured summary of a study protocol for a randomised controlled trial. Trials. 2021 Feb 5;22(1):116.

51. Kulkarni S, Fisk M, Kostapanos M, Banham-Hall E, Bond S, Hernan-Sancho E, et al. Repurposed immunomodulatory drugs for Covid-19 in pre-ICu patients - mulTi-Arm Therapeutic study in pre-ICu patients admitted with Covid-19 – Repurposed Drugs (TACTIC-R): A structured summary of a study protocol for a randomised controlled trial. Trials. 2020 Jul 8;21(1):626.

52. NIH closes enrollment in trial comparing COVID-19 treatment regimens [Internet]. National Institutes of Health (NIH). 2021 [cited 2021 Jul 28]. Available from: https://www.nih.gov/news-events/news-releases/nih-closes-enrollment-trial-comparing-covid-19-treatment-regimens

53. Overview | Baricitinib for treating moderate to severe atopic dermatitis | Guidance | NICE [Internet]. NICE; [cited 2021 Dec 9]. Available from: https://www.nice.org.uk/guidance/ta681

54. Update on Omicron [Internet]. [cited 2021 Dec 9]. Available from: https://www.who.int/news/item/28-11-2021-update-on-omicron

55. Hospitalized Adults: Therapeutic Management | COVID-19 Treatment Guidelines [Internet]. [cited 2021 Dec 9]. Available from: https://www.covid19treatmentguidelines.nih.gov/management/clinical-management/hospitalized-adults--therapeutic-management/

56. Therapeutics and COVID-19: living guideline [Internet]. [cited 2021 Dec 9]. Available from: https://app.magicapp.org/#/guideline/5666

57. COVID_Management_Algorithm_23092021.pdf [Internet]. [cited 2021 Dec 9]. Available from: https://www.icmr.gov.in/pdf/covid/techdoc/COVID_Management_Algorithm_23092021.pdf

